# Prophylaxis for covid-19: living systematic review and network meta-analysis

**DOI:** 10.1101/2021.02.24.21250469

**Authors:** Jessica J Bartoszko, Reed AC Siemieniuk, Elena Kum, Anila Qasim, Dena Zeraatkar, Long Ge, Mi Ah Han, Behnam Sadeghirad, Arnav Agarwal, Thomas Agoritsas, Derek K Chu, Rachel Couban, Andrea J Darzi, Tahira Devji, Maryam Ghadimi, Kimia Honarmand, Ariel Izcovich, Assem Khamis, Francois Lamontagne, Mark Loeb, Maura Marcucci, Shelley L McLeod, Shahrzad Motaghi, Srinivas Murthy, Reem A Mustafa, John D Neary, Hector Pardo-Hernandez, Gabriel Rada, Bram Rochwerg, Charlotte Switzer, Britta Tendal, Lehana Thabane, Per O Vandvik, Robin WM Vernooij, Andrés Viteri-García, Ying Wang, Liang Yao, Zhikang Ye, Gordon H Guyatt, Romina Brignardello-Petersen

**Author notes:** Corresponding author: Reed AC Siemieniuk. Joint first authors (Jessica J Bartoszko, Reed AC Siemieniuk, Elena Kum, Anila Qasim, Dena Zeraatkar, Long Ge). Critical care physician (Kimia Honarmand, Francois Lamontagne, Bram Rochwerg). Data analyst (Assem Khamis).

## Abstract

**Objective:** To determine and compare the effects of drug prophylaxis on severe acute respiratory syndrome coronavirus virus 2 (SARS-CoV-2) infection and coronavirus disease 2019 (covid-19).

**Design:** Living systematic review and network meta-analysis.

**Data sources:** WHO covid-19 database, a comprehensive multilingual source of global covid-19 literature to 19 January 2021, and six additional Chinese databases to 20 January 2021.

**Study selection:** Randomized trials in which people at risk of covid-19 were randomized to drug prophylaxis or no prophylaxis (standard care or placebo). Pairs of reviewers independently screened potentially eligible articles.

**Methods:** After duplicate data abstraction, we conducted random-effects bayesian network meta-analysis. We assessed risk of bias of the included studies using a modification of the Cochrane risk of bias 2.0 tool and assessed the certainty of the evidence using the grading of recommendations assessment, development and evaluation (GRADE) approach.

**Results:** The first iteration of this living network meta-analysis includes nine randomized trials – six addressing hydroxychloroquine (6,059 participants), one addressing ivermectin combined with iota-carrageenan (234 participants) and two addressing ivermectin alone (540 participants), all compared to standard care or placebo. Hydroxychloroquine has no important effect on admission to hospital (risk difference (RD) 1 fewer per 1,000, 95% credible interval (CrI) 3 fewer to 4 more, high certainty) or mortality (RD 1 fewer per 1,000, 95% CrI 2 fewer to 3 more, high certainty). Hydroxychloroquine probably has no important effect on laboratory-confirmed infection (RD 2 more per 1,000, 95% CrI 18 fewer to 28 more, moderate certainty), probably increases adverse effects leading to drug discontinuation (RD 19 more per 1,000, 95% CrI 1 fewer to 70 more, moderate certainty) and may have no important effect on suspected, probable or laboratory-confirmed infection (RD 15 fewer per 1,000, 95% CrI 64 fewer to 41 more, low certainty). Due to serious risk of bias and very serious imprecision – and thus very low certainty evidence, the effects of ivermectin combined with iota-carrageenan on laboratory-confirmed infection (RD 52 fewer per 1,000, 95% CrI 58 fewer to 37 fewer), and ivermectin alone on laboratory-confirmed infection (RD 50 fewer per 1,000, 95% CrI 59 fewer to 16 fewer) and suspected, probable or laboratory-confirmed infection (RD 159 fewer per 1,000, 95% CrI 165 fewer to 144 fewer) remain uncertain.

**Conclusion:** Hydroxychloroquine prophylaxis does not have an important effect on hospital admission and mortality, probably increases adverse effects, and probably does not have an important effect on laboratory-confirmed SARS-CoV-2 infection. Because of serious risk of bias and very serious imprecision, we are highly uncertain whether ivermectin combined with iota-carrageenan and ivermectin alone reduce the risk of SARS-CoV-2 infection.

**Systematic review registration:** This review was not registered. The protocol established *a priori* is included as a supplement.

**Funding:** This study was supported by the Canadian Institutes of Health Research (grant CIHR-IRSC:0579001321).

**Readers’ note:** This article is a living systematic review that will be updated to reflect emerging evidence. Updates may occur for up to two years from the date of original publication.

## Introduction

As of 18 February 2021, more than 110 million people have been infected with severe acute respiratory syndrome coronavirus virus 2 (SARS-CoV-2), the virus responsible for coronavirus disease 2019 (covid-19); of these, more than 2.4 million have died.^1^ Cases and deaths continue to rise, with resurgence of the virus occurring in many countries that previously gained control. Drugs that are effective as prophylaxis, used either shortly after exposure (post-exposure prophylaxis, PEP) or prior to exposure in a high-risk population (pre-exposure prophylaxis, PrEP), to prevent or attenuate infection could have a monumental impact worldwide. Therefore, researchers around the world are enrolling participants in randomized trials of drugs for prophylaxis against covid-19.

A common understanding of the evidence from these randomized trials may be challenging for healthcare workers and people interested in using a drug for prophylaxis. Timely evidence summaries and associated guidelines could ameliorate the problem.^2^ Clinicians, patients, guideline bodies, and government agencies also face challenges of interpreting the results from trials that contribute to a rapidly evolving evidence-base. This environment necessitates well developed summaries that distinguish between trustworthy and untrustworthy evidence.

Living systematic reviews and network meta-analyses resolve an important limitation of traditional systematic reviews—that of providing an overview of the relevant evidence only at a specific point in time.^3^ The ability of a living network meta-analysis to present a complete, broad, and up-to-date view of the evidence makes it ideal to inform the development of practice recommendations, ideally in the form of living clinical practice guidelines.^3-5^ Network meta-analysis, rather than pairwise meta-analysis, provides useful information about the comparative effectiveness of treatments that have not been tested head-to-head. The lack of such direct comparisons is certain to limit inferences in the covid-19 setting; therefore, network meta-analysis is critical to inform the selection of the best drug among all alternative options. Moreover, the incorporation of indirect evidence can strengthen evidence in comparisons that were tested head to head.^6^

In this living systematic review and network meta-analysis, we compare the effects of drug prophylaxis for covid-19. This living network meta-analysis will – similarly to our established living network meta-analysis on covid-19 treatment^7^ – directly inform living WHO guidelines on drugs for covid-19^4,5^, a collaborative effort between WHO and the MAGIC Evidence Ecosystem Foundation (www.magicproject.org), inspired by the *BMJ* Rapid Recommendations.^8,9^ This review will inform trustworthy, actionable, and living guidance to clinicians caring for and patients with covid-19.

## Methods

The protocol provides detailed methods of this systematic review (see supplementary file). We report this living systematic review following the guidelines of the preferred reporting items for systematic reviews and network meta-analyses (PRISMA) checklist.^10^ A living systematic review is a cumulative synthesis updated regularly as new evidence becomes available.^11^ The linked *BMJ* Rapid Recommendations methods team approved all decisions relevant to data synthesis.

### Eligibility criteria

We included randomized trials of people at risk of covid-19 that compared drugs for prophylaxis against one another or against no intervention, placebo, or standard care, with no restriction on language. We included studies addressing patients with pre- or post-exposure status and risk groups (i.e. unexposed community member, member of the same household with one or more positive cases, contact of index case, healthcare worker or long-term care resident). We also included trials of traditional Chinese medicines if the drug comprised one or more specific molecules with a defined molecular weight dosing.

We excluded randomized trials published only as press releases and that evaluated vaccination, drug treatments, antibody and cellular therapies, nutrition, and non-drug supportive care interventions. We identified and separately categorized trials that evaluated these interventions. We synthesize randomized trials that evaluate drug treatments, and antibody and cellular therapies for covid-19 in separate living network meta-analyses.^7^

### Information sources

We performed daily searches from Monday to Friday in the World Health Organization (WHO) covid-19 database for eligible studies – a comprehensive multilingual source of global literature on covid-19.^12^ Prior to its merge with the WHO covid-19 database on 9 October 2020, we performed daily searches from Monday to Friday in the US Centers for Disease Control and Prevention (CDC) covid-19 Research Articles Downloadable Database for eligible studies.^13^ The database includes, but is not limited to the following 25 bibliographic and grey literature sources: Medline (Ovid and PubMed), PubMed Central, Embase, CAB Abstracts, Global Health, PsycInfo, Cochrane Library, Scopus, Academic Search Complete, Africa Wide Information, CINAHL, ProQuest Central, SciFinder, the Virtual Health Library, LitCovid, WHO covid-19 website, CDC covid-19 website, Eurosurveillance, China CDC Weekly, Homeland Security Digital Library, ClinicalTrials.gov, bioRxiv (preprints), medRxiv (preprints), chemRxiv (preprints), and SSRN (preprints). The supplementary file includes the CDC literature search strategy.

We designed the daily searches to match the update schedule of the database and capture eligible studies the day of or the day after publication. To identify randomized trials, we filtered the results through a validated and highly sensitive machine learning model.^14^ We tracked preprints of randomized trials until publication and, when discrepant, updated data to match that in the peer-reviewed publication. When needed, we reconciled multiple versions of preprints, post-hoc analyses, corrections and retractions.

In addition, monthly searches, utilizing adapted search terms for covid-19 developed by the CDC for the Chinese language, included six Chinese databases: Wanfang, Chinese Biomedical Literature, China National Knowledge Infrastructure, VIP, Chinese Medical Journal Net (preprints), and ChinaXiv (preprints). The Chinese literature search also included search terms for randomized trials (available upon request to corresponding author).

The search included monitoring, on an ongoing basis, living evidence retrieval services including the Living Overview of the Evidence (L-OVE) covid-19 Repository by the Epistemonikos Foundation and in collaboration with the Cochrane Canada Centre at McMaster University, the Systematic and Living Map on covid-19 Evidence by the Norwegian Institute of Public Health,.^15,16^

The search included all English information sources from 1 December 2019 to 19 January 2021, and the Chinese literature from conception of the databases to 20 January 2021.

### Study selection

Using a systematic review software, Covidence,^17^ following training and calibration exercises, pairs of reviewers independently screened all titles and abstracts, followed by full texts of trials that were identified as potentially eligible. A third reviewer adjudicated conflicts.

### Data collection

For each eligible trial, following training and calibration exercises, pairs of reviewers extracted data independently using a standardised, pilot tested data extraction form. Reviewers collected information on trial characteristics (trial registration, publication status, study status, design), participant characteristics (country, age, sex, comorbidities), exposure characteristics (exposure status, exposure duration, high risk group) and outcomes of interest (means or medians and measures of variability for continuous outcomes and the number of participants analysed and the number of participants who experienced an event for dichotomous outcomes). Reviewers resolved discrepancies by discussion and, when necessary, with adjudication by a third party.

The review team selected outcomes of interest based on importance to patients and these were informed by clinical expertise in the systematic review team and the linked guideline panel responsible for the WHO living guidelines for covid-19.^4,5^ The panel includes unconflicted clinical experts and patient partners, and was recruited to ensure global representation. We rated outcomes from 1 to 9 based on importance to individual patients (9 being most important), and we included any outcome rated 7 or higher by any panel member. This process resulted in choice of the following outcomes: laboratory-confirmed SARS-CoV-2 infection; a composite of suspected, probable or laboratory-confirmed SARS-CoV-2 infection; admission to hospital (within 28 days); mortality (closest to 90 days); adverse effects leading to discontinuation (within 28 days); and time to symptom resolution or clinical improvement in the subset of participants that became infected with SARS-CoV-2. In anticipation for inclusion in the linked WHO recommendation for prophylactic drugs, data for drug-specific adverse effects included trials reporting on hydroxychloroquine versus standard care or placebo. For only the first iteration of this living systematic review, the supplementary file includes the results of pairwise meta-analyses and related subgroup analyses for cardiac toxicity and non-serious gastrointestinal adverse effects.

Because of inconsistent reporting across trials, when possible we preferentially extracted participant characteristics and outcome data for participants PCR-negative for SARS-CoV-2 infection at baseline. If authors did not report data separately for those who were PCR-negative for SARS-CoV-2 infection at baseline, we extracted data from all participants, regardless of their PCR status at baseline.

### Risk of bias within individual studies

For each eligible trial, following training and calibration exercises, reviewers used a revision of the Cochrane tool for assessing risk of bias in randomized trials (RoB 2.0)^18^ to rate trials at the outcome level as either at i) low risk of bias, ii) some concerns—probably low risk of bias, iii) some concerns—probably high risk of bias, or iv) high risk of bias, across the following domains: bias arising from the randomization process; bias due to departures from the intended intervention; bias due to missing outcome data; bias in measurement of the outcome; bias in selection of the reported results, including deviations from the registered protocol; bias arising from early termination for benefit; and bias arising from competing risks. We rated trials at high risk of bias overall if one or more domains were rated as some concerns—probably high risk of bias or as high risk of bias and as low risk of bias overall if all domains were rated as some concerns—probably low risk of bias or low risk of bias. Reviewers resolved discrepancies by discussion and, when not possible, with adjudication by a third party.

### Data synthesis

#### Summary measures

We summarised the effect of interventions on dichotomous outcomes using odds ratios (ORs) and corresponding 95% credible intervals (CrIs). To mitigate results with highly implausible and extremely imprecise effect estimates, the analyses included only prophylactic drugs with at least 100 participants or 20 events, regardless of the number of studies in which the drug was assessed or the number of participants who received the drug in each study.^7^ The analysis plan included, data permitting, adjustment for cluster randomization.

#### Treatment nodes

We created nodes for each prophylactic drug (or combination of drugs), independent of dose or duration. Standard care and placebo arms across included trials were combined into a single node for analyses. The *networkplot* command of Stata version 15.1 (StataCorp, College Station, TX) provided software for network plots in which the inverse variance of the direct comparison determined the thickness of lines between nodes and the size of nodes.^19^

#### Statistical analysis

For outcomes with sufficient data (i.e. laboratory-confirmed SARS-CoV-2 infection, and suspected, probable or laboratory-confirmed SARS-CoV-2 infection), we performed random effects network-meta analysis using the R package *gemtc*^20^ and used three Markov chains with 100 000 iterations after an initial burn-in of 10 000 and a thinning of 10. Node splitting models provided methods to obtain indirect estimates and to assess local heterogeneity.^21^ For all other outcomes, we performed random-effects bayesian meta-analysis using *bayesmeta* package in RStudio version 3.5.3 (R Studio, Boston, MA, USA).^22^ An empirical study provided the basis for choosing a plausible prior for the variance parameter and a uniform prior for the effect parameter.^23^

### Certainty of the evidence

The grading of recommendations assessment, development and evaluation (GRADE) approach for network meta-analysis provided the framework for assessing the certainty of evidence.^6,24^ Two methodologists with experience in using GRADE rated each domain for each comparison separately and resolved discrepancies by discussion. Criteria for rating the certainty for each comparison and outcome as high, moderate, low, or very low, included considerations of risk of bias, inconsistency, indirectness, publication bias, intransitivity, incoherence (difference between direct and indirect effects) and imprecision.^24^ Judgments of imprecision for this systematic review were made using a minimally contextualised approach.^25^ The minimally contextualised approach considers whether credible intervals include the null effect or when the point estimate is close to the null effect, whether the credible interval lies within the boundaries of small but important benefit and harm.

We rated the certainty of no important effect for the outcomes laboratory-confirmed infection; suspected, probable or laboratory-confirmed infection; admission to hospital; and mortality. Pending data from quantitative studies of patient values, we chose thresholds of small, but important effects of 0.5% for mortality, 3% for infection (whether laboratory-confirmed or not), and 1% for admission to hospital.^25^ We rated the certainty that there is an increase or decrease in adverse effects leading to discontinuation using the null effect as a threshold. GRADE evidence summaries (Summary of Findings tables) in the MAGIC Authoring and Publication Platform (www.magicapp.org) provided user friendly formats for clinicians and patients, and allowed re-use in the context of clinical practice guidelines for covid-19, such as the WHO living guidelines.^4,5^ Interim updates and additional study data will appear on our website (www.covid19lnma.com).

### Interpretation of results

To facilitate interpretation of the results, we calculated absolute effects for outcomes in which the summary measure was an OR. For mortality, we used the event rate among all participants randomized to standard care or placebo to calculate the baseline risk. For all other outcomes, we use the median event rate in the standard care or placebo arms to calculate the baseline risk.

### Subgroup and sensitivity analysis

The analysis plan includes performing subgroup analyses of pre-exposure versus post-exposure studies, preprints versus peer-reviewed studies and high versus low risk of bias studies when there are at least two studies in each subgroup. We plan to perform network meta-regression to explore if duration of prophylactic drug use may modify the relative effect of the drug on adverse effects leading to discontinuation hypothesizing that, if the drug is active at the time of exposure, it will have a greater relative effect. The linked independent WHO guideline panels may direct, in the future, additional subgroup analyses; in this first report, the panel provided direction to perform subgroup analyses by drug prophylaxis duration and dose. The Credibility of Effect Modification Analyses in randomized controlled trials and meta-analyses (ICEMAN) tool provides the methodology for, whenever statistical evidence of a subgroup effect exists, assessing subgroup hypothesis credibility.^26^

### Patient and public involvement

As part of the WHO living guidelines and *BMJ* Rapid Recommendations initiative, patients participated in defining clinical questions and rating the importance of outcomes for this systematic review and are also involved in the interpretation of results and the generation of parallel recommendations.

## Results

As of 19 January 2021, after screening 25,763 titles and abstracts and 479 full texts, ten unique randomized trials that evaluated prophylactic drugs proved eligible (fig 1) – six addressing hydroxychloroquine; one ivermectin combined with iota-carrageenan; two ivermectin alone; and one ramipril.^27-36^ Searches of living evidence retrieval services identified two of these eligible randomized trials.^31,33^ The supplementary files includes a table of excluded full texts.

**Figure 1.**
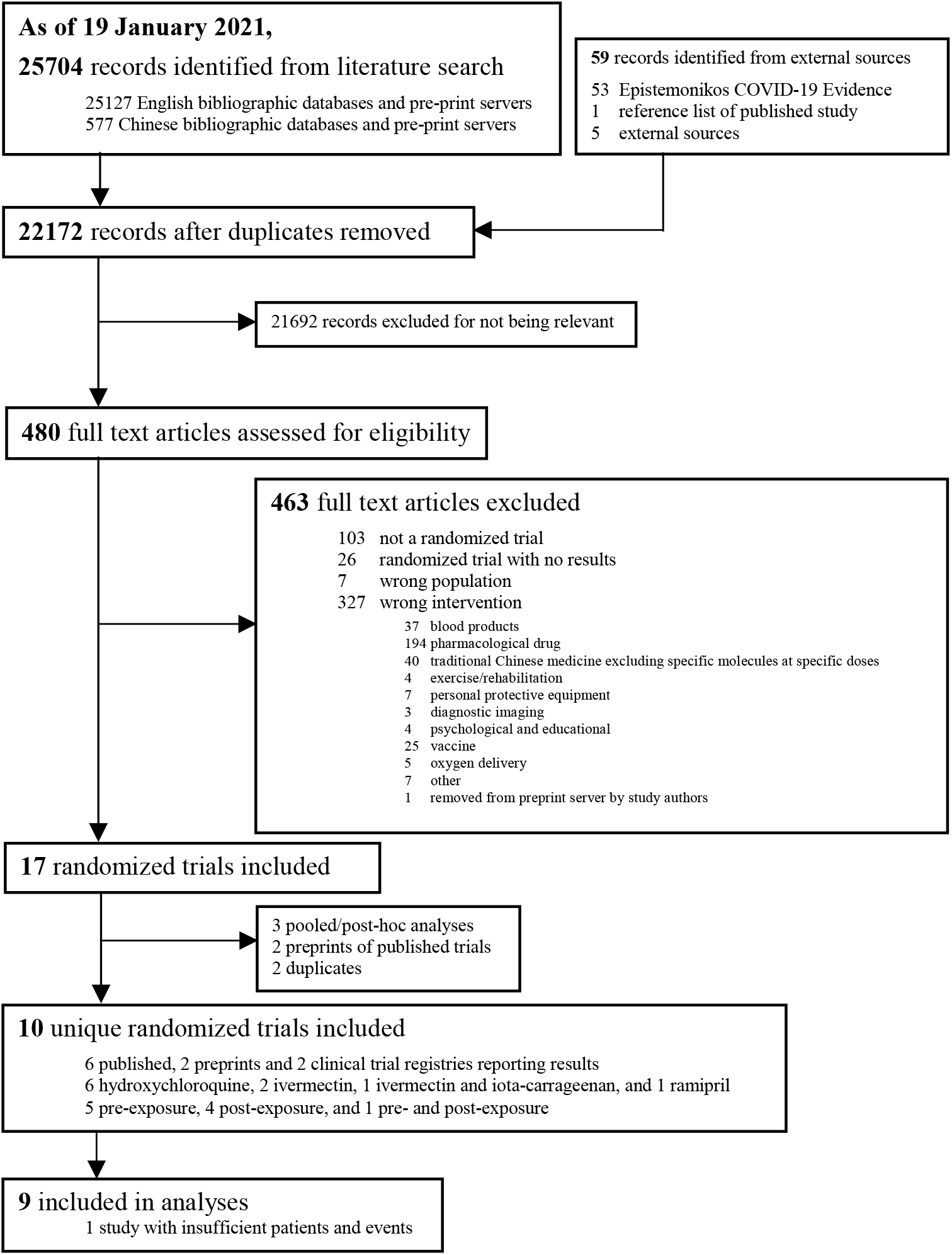
Study selection.

Of the ten eligible randomized trials, six were published in peer-reviewed journals^27-30,32,34^, two only as preprint^31,33^, and two were from clinical trial registries that reported results.^35,36^ All trials were registered, published in English and evaluated prophylactic drugs against standard care or placebo.^27-36^ Five evaluated prophylactic drugs in participants without documented exposure to covid-19^27,30-32,36^ and four evaluated prophylactic drugs in participants with documented exposure to covid-19.^28,29,34,35^ One randomized trial evaluated prophylactic ivermectin in both participants pre-exposure and post-exposure to covid-19.^33^ Table 1 presents the characteristics of the ten included studies, including prophylactic drug dose and duration, and extent and setting of participant exposure to covid-19.

**Table 1.**
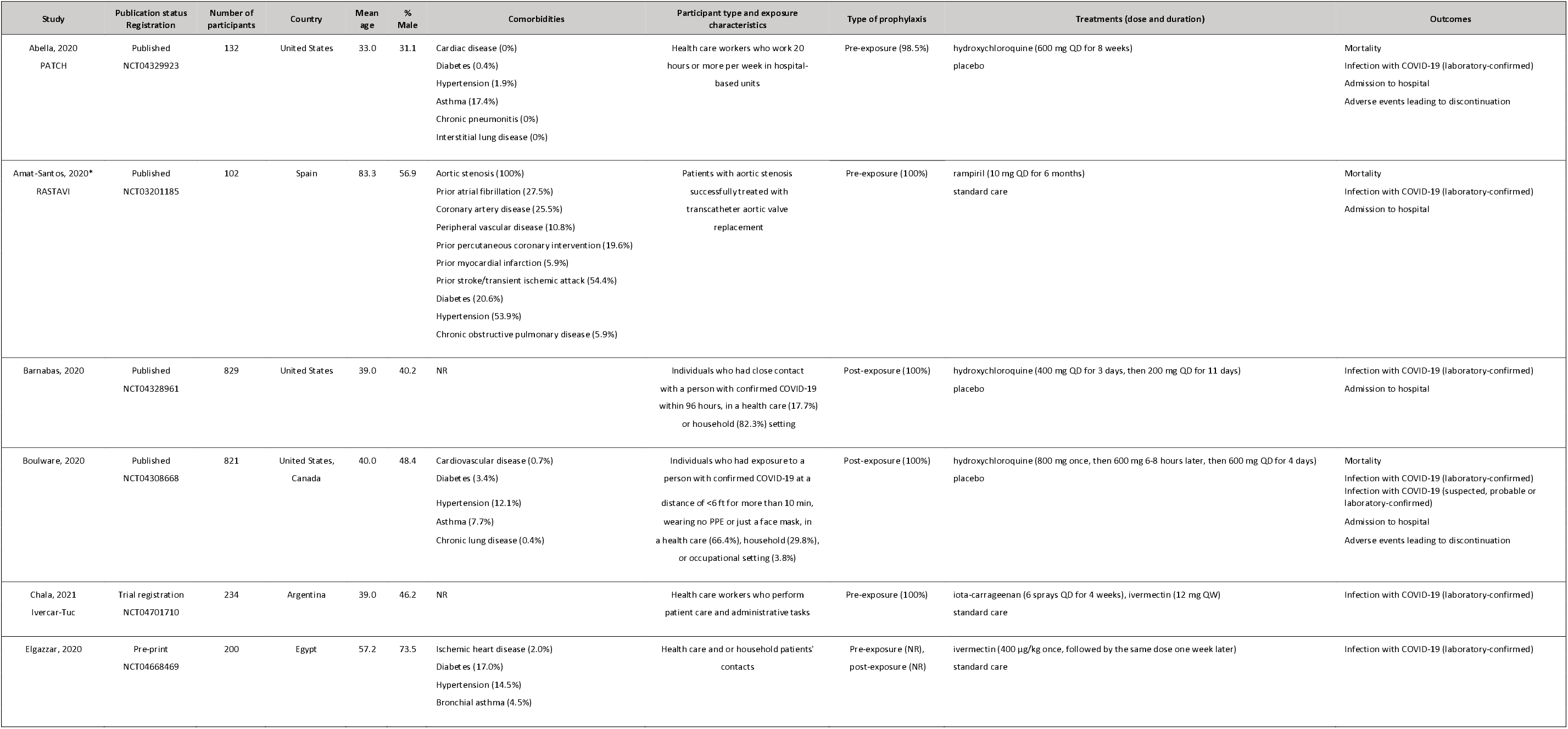
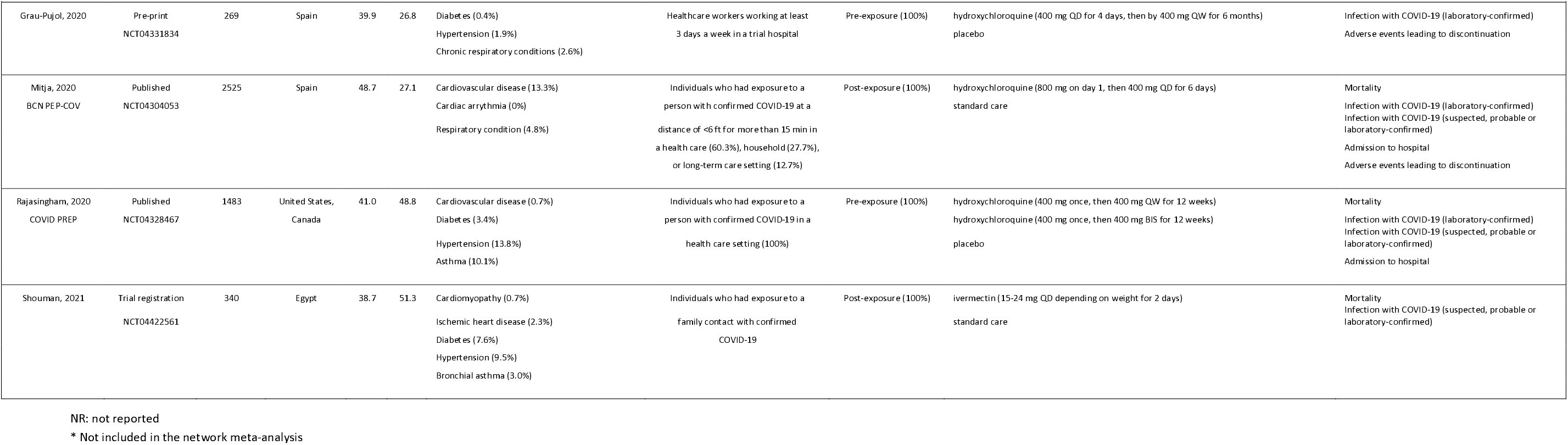
Study characteristics.

Six trials – one addressing ramipril, three hydroxychloroquine, and two ivermectin – reported results for one or more outcomes of interest that were not prespecified in protocols or registrations.^27,30,31,33-35^ The protocols and trial reports included no other discrepancies in reporting of our outcomes of interest. Two trials were initially posted as preprints and subsequently published in full after peer review.^29,32^ One trial published multiple iterations of their preprint.^33^ The supplementary file presents additional study characteristics, outcome data, and reporting differences between versions of study preprints and/or peer-reviewed publications.

We performed the analyses on 18 January 2021 and included nine randomized trials that evaluated hydroxychloroquine and ivermectin (with and without iota-carrageenan) prophylaxis against no prophylaxis (standard care or placebo).^28-36^ One randomized trial that evaluated ramipril against standard care, not included in the network meta-analysis because it enrolled less than 100 patients and observed less than 20 events in the ramipril arm, reported that ramipril had no effect on the incidence of SARS-CoV-2 infection or severity of covid-19.^27^ Since molecule, not dose or duration, dictates choice of nodes, we combined the two active arms in one included three-arm trial – hydroxychloroquine once weekly and hydroxychloroquine twice weekly.^32^ In this report, because the authors did not report the intracluster correlation coefficient, we could not adjust for cluster randomization in one analysed trial addressing hydroxychloroquine.^29^ A published post-hoc analysis with one of the analysed trials addressing hydroxychloroquine did not include information beyond what was already reported in the original peer-reviewed publication of the trial.^28,37^

### Risk of bias in included studies

The supplementary material presents the assessment of risk of bias of the ten included studies for each outcome. Five studies addressing hydroxychloroquine proved at low risk of bias across all outcomes.^28,30-32,34^ Five studies proved at high risk of bias overall.^27,29,33,35,36^

### Effects of the interventions

The supplementary material presents the network and forest plots depicting the interventions included in the network meta-analysis of each outcome. The supplementary file also presents detailed relative and absolute effect estimates and certainty of the evidence for all comparisons and outcomes. We did not detect statistical incoherence in any of the comparisons or outcomes. Five trials evaluated hydroxychloroquine against placebo^28,30-32,34^ and four trials – one addressing hydroxychloroquine,^29^ one ivermectin combined with iota-carrageenan^36^ and two ivermectin alone^33,35^ – were evaluated against standard care, defined as no specific therapy^29,35^, standard biosecurity care^36^ and personal protective measures.^33^ Figure 2 presents a summary of the effects of hydroxychloroquine, ivermectin combined with iota-carrageenan and ivermectin alone on the outcomes, and we describe these results herein.

**Figure 2.**
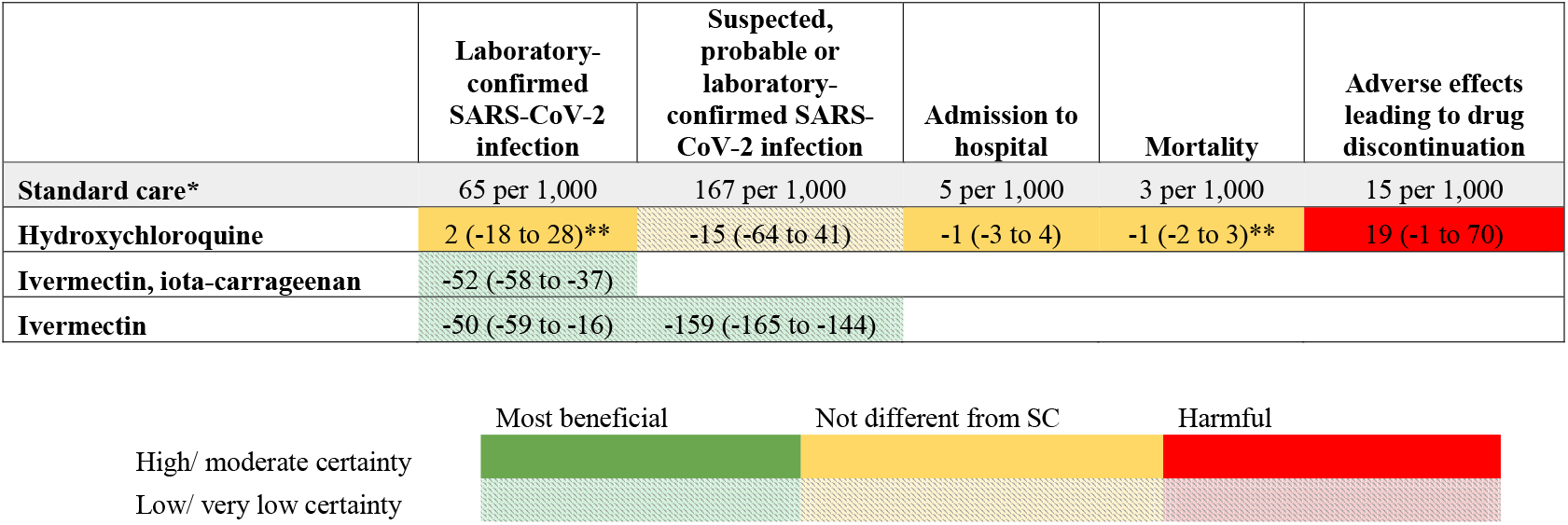
Summary of effects compared with standard care (SC) * The expected risk of each outcome with standard care is reported in the grey row. ** The best estimate of effect was obtained from direct evidence Empty cells: there was no evidence for the specific intervention Numbers in the coloured cells are the estimated risk differences (95% CI) per 1000 patients or mean difference (95% CI) in days when compared to standard care.

#### Laboratory-confirmed SARS-CoV-2 infection

Eight trials reported on laboratory-confirmed infection in 5,728 participants who were PCR-negative at baseline and were included in random-effects bayesian network meta-analysis.^28-34,36^ The network nodes included were hydroxychloroquine, ivermectin combined with iota-carrageenan, ivermectin alone and standard care or placebo. Hydroxychloroquine, compared to standard care or placebo, probably has no effect on laboratory-confirmed infection (OR 1.03, 95% CrI 0.71 to 1.47, RD 2 more per 1,000, 95% CrI 18 fewer to 28 more, moderate certainty). We are uncertain whether ivermectin combined with iota-carrageenan, when compared to standard care, reduces the risk of laboratory-confirmed infection (OR 0.12, 95% CrI 0.03 to 0.38, RD 52 fewer per 1,000, 95% CrI 58 fewer to 37 fewer, very low certainty) (Fig 2). We are also uncertain whether ivermectin alone, when compared to standard care, reduces risk of laboratory-confirmed infection (OR 0.16, 95% CrI 0.02 to 0.73, RD 50 fewer per 1,000, 95% CrI 59 fewer to 16 fewer, very low certainty).

#### Suspected, probable or laboratory-confirmed SARS-CoV-2 infection

Four trials reported on the composite of suspected, probable or laboratory-confirmed infection in 4,531 participants and were included in random-effects bayesian network meta-analysis consisting of hydroxychloroquine, ivermectin and standard care or placebo nodes.^28,29,32,35^ For the outcome suspected, probable and laboratory-confirmed infection, low certainty evidence suggests that hydroxychloroquine, compared to standard care or placebo, may have no effect (OR 0.90, 95% CrI 0.58 to 1.31; RD 15 fewer per 1,000 participants, 95% CrI 64 fewer to 41 more). Due to very low certainty evidence, the effect of ivermectin, compared to standard care, in reducing the risk of suspected, probable or laboratory-confirmed infection remains uncertain (OR 0.06, 95% CrI 0.02 to 0.13; RD 159 fewer per 1,000 participants, 95% CrI 165 fewer to 144 fewer).

#### Hospital admission

Five trials reported hospital admission in 5,659 participants randomized to hydroxychloroquine, standard care or placebo.^28-30,32,34^ Trials on other prophylactic drugs eligible for analysis did not report on the outcome hospital admission, precluding network meta-analysis. Hydroxychloroquine has no important effect on hospital admission when compared to standard care or placebo (OR 0.87, 95% CrI 0.42 to 1.77, RD 1 fewer per 1,000 participants, 95% CrI 3 fewer to 4 more, high certainty) (Fig 2).

#### Mortality

Five trials reported mortality in 5,153 participants randomized to hydroxychloroquine, ivermectin, standard care, or placebo.^28-30,32,35^ Although we had sufficient data to perform network meta-analysis, the network did not converge; therefore, we present results from pairwise meta-analyses. Hydroxychloroquine has no important effect on mortality when compared to standard care or placebo (OR 0.70, 95% CrI 0.24 to 1.99; RD 1 fewer per 1,000 participants, 95% CrI 2 fewer to 3 more, high certainty) (Fig 2). Because there were no deaths in the one ivermectin trial reporting mortality, we are very uncertain about its effect on this patient-important outcome.

#### Adverse effects leading to drug discontinuation

Four trials reported adverse effects leading to drug discontinuation in 3,616 participants randomized to hydroxychloroquine, standard care, or placebo.^28-31^ Trials on other prophylactic drugs eligible for analysis did not report any adverse effects leading to drug discontinuation, precluding network meta-analysis. Hydroxychloroquine probably increases adverse effects when compared to standard care or placebo (OR 2.34, 95% CrI 0.93 to 6.08, RD 19 more per 1,000 participants, 95% CrI 1 fewer to 70 more, moderate certainty) (Fig 2).

#### Time to symptom resolution or clinical improvement

No randomized trials reported on time to symptom resolution or clinical improvement in the subset of participants that developed SARS-CoV-2 infection.

### Subgroup analysis

Insufficient data precluded subgroup analysis for trials randomizing patients to ivermectin alone or ivermectin with iota-carrageenan versus standard care. Thus, we limited subgroup analysis to hydroxychloroquine trials only. We did not find any statistical evidence of differences in laboratory-confirmed SARS-CoV-2 infection, suspected, probable or laboratory-confirmed SARS-CoV-2 infection, hospital admission, or adverse effects leading to discontinuation between pre-exposure^30-32^ and post-exposure^28,29,34^ studies or based on hydroxychloroquine dosing regimens (see supplementary file). Extremely low event rates precluded investigation of subgroup effects for mortality.

## Discussion

This living systematic review and network meta-analysis provides a comprehensive overview of the evidence for prophylaxis against covid-19 up to 19 January 2021 and directly informs WHO living guidelines on prophylaxis.^5^ Hydroxychloroquine versus no prophylaxis, ivermectin with iota-carrageenan versus no prophylaxis and ivermectin versus no prophylaxis were the comparisons for which there was informative evidence. Hydroxychloroquine probably increases adverse effects leading to drug discontinuation (moderate certainty evidence). For other outcomes (laboratory-confirmed SARS-CoV-2 infection, hospital admission and mortality), study results provide moderate and high certainty evidence – none of which support any benefit to hydroxychloroquine. Due to serious risk of bias and very serious imprecision, we are highly uncertain whether ivermectin combined with iota-carrageenan and ivermectin alone reduce the risk of patient-important outcomes.

These findings are consistent with those reported in a meta-analysis of hydroxychloroquine prophylaxis against no prophylaxis, which did not find any statistical evidence of a benefit with hydroxychloroquine prophylaxis for the patient-important outcomes of SARS-CoV-2 infection, hospital admission and death.^38^ The study also concluded that hydroxychloroquine is likely to increase the risk of adverse effects.^38^ The risk of death is much lower in people at risk of covid-19 compared to those diagnosed with covid-19.^7^ Similarly, the risk of SARS-CoV-2 infection varies depending on pre- or post-exposure status and setting. Therefore, prophylactic research necessitates very large trials and/or a focus on the highest risk populations to detect a possible benefit on outcomes of most importance to patients. Further, rare but important harms may not be detected with randomized trials unless they enrol an extremely large sample size, diligently follow-up to ascertain these outcomes, or include patients at greater risk. Guideline panels, which independently rate the certainty of the evidence, therefore have to consider the trade-offs between concluding probably no benefit and meaningful adverse effects, and waiting for more precise data. For example, the WHO living guidelines issued a strong recommendation against hydroxychloroquine for prophylaxis in covid-19.^5^

### Strengths and limitations of this review

This, the first network meta-analysis published on prophylactic drugs for covid-19, incorporates the most up-to-date evidence on hydroxychloroquine, ivermectin with iota-carrageenan and ivermectin alone. It adds to our living systematic review on drugs for covid-19 and directly informs the WHO living guidelines, together constituting major innovations in the evidence ecosystem.^5^

The search strategy was comprehensive with explicit eligibility criteria, and no restrictions on the language of publication. To ensure expertise in all areas, our team includes clinical and methods experts who have undergone training and calibration exercises for all stages of the review process. In order to avoid spurious findings, we prespecified that we would only analyse drugs to which at least 100 people had been randomized or 20 events have been observed. The single trial that evaluated ramipril against standard care was therefore omitted from the network meta-analysis, necessitated by the priority to avoid issues that arise from network meta-analysis of sparse data (uninformative and implausible results).^7^

The GRADE approach provided the structure for rating certainty of evidence and interpreting the results considering absolute effects. To rate the GRADE domain of imprecision, we prespecified thresholds of effect that most would consider small but important. In the absence of empiric data, these thresholds represent our collective experience but are, to a large extent, arbitrary. People placing a larger or smaller value on certain outcomes may reasonably make different inferences about the certainty of evidence for no important effect. For example, people who consider that the smallest important effect in mortality is increasing or reducing 2 per 1000 or more deaths, would rate down the certainty of the evidence due to imprecision and conclude that hydroxychloroquine probably does not have an effect on this outcome (i.e. moderate certainty).

With regard to limitations of the review, some conclusions are based on very low certainty evidence and we therefore anticipate future studies evaluating ivermectin for prophylaxis may substantially change the results, particularly for outcomes of infection and mortality.^39^ One cluster randomized trial did not report the design effect or the intracluster correlation coefficient itself necessary to calculate the design effect, precluding adjustment in analyses – potentially leading to falsely narrow credible intervals.^29^ Cluster sizes were, however, small, making substantial bias unlikely.

The living nature of our systematic review and network meta-analysis could amplify publication bias, because studies with promising results are more likely to be published sooner than studies with negative results. Given the failure of hydroxychloroquine trials to show benefit, this is not a concern for hydroxychloroquine. This is, however, a concern for the evidence to date on ivermectin, for which none of the data has been peer-reviewed. With the inclusion of this data in network meta-analysis from one preprint^33^ and two clinical trial registries reporting results,^35,36^ we found evidence of large positive effects; however, bias from simple errors and reporting limitations might have been introduced. We include these data, regardless of publication status and risk of bias, because of the urgent need for information and because so many of the studies on covid-19 are published first as preprints.

Another limitation of the evidence to date is lack of blinding, which might introduce bias through differences in co-interventions between randomization groups, especially when measuring the outcomes clinically suspected and probable infection, and adverse effects leading to discontinuation of the drug. We chose to consider the treatment arms that did not receive an active experimental drug (i.e., placebo or standard care) within the same node: it is possible that unblinded standard care groups^29,33,35,36^ may have received systematically different co-interventions or changed their personal protective behaviours when compared to groups randomized to receive a placebo.^28,30-32,34^ Laboratory confirmation mitigates risk of bias from lack of blinding in outcome measurement; however, the availability of diagnostic testing differs across health systems, warranting the additional use of a symptomatic case definition for infection. This was the case for the majority of participants, including healthcare workers, enrolled in one study in the United States, which risked overestimating incidence of infection by using a symptomatic definition for infection.^28,40^

We will periodically update this living systematic review and network meta-analysis. The changes from each version will be highlighted for readers and the most updated version will be the one available in the publication platform. Previous versions will be archived in the supplementary material. This living systematic review and network meta-analysis will also be accompanied by an interactive infographic and a website for users to access the most updated results in a user friendly format (http://app.magicapp.org/public/guideline/L6RxYL, www.covid19lnma.com)

## Conclusions

This living systematic review and network meta-analysis on prophylactic drugs for covid-19 provides evidence that hydroxychloroquine does not have an important effect on mortality and hospital admission, probably increases adverse effects, and probably does not have an important effect on laboratory-confirmed SARS-CoV-2 infection. We are very uncertain if ivermectin with or without iota-carrageenan reduces the risk of SARS-CoV-2 infection and mortality because of serious risk of bias, very serious imprecision and the effect estimates are likely to change substantially with additional evidence from ongoing trials. No other drug has been studied in large enough trials to make any inferences regarding effects of prophylaxis for covid-19.

## Supporting information

PRISMA Reporting Checklist

## Data Availability

No additional data available.

## Acknowledgements

DKC is a CAAIF-CSACI-AllerGen Emerging Clinician-Scientist Research Fellow, supported by the Canadian Allergy, Asthma and Immunology Foundation (CAAIF), the Canadian Society of Allergy and Clinical Immunology (CSACI), and AllerGen NCE (the Allergy, Genes and Environment Network, funded through the Canadian Institutes of Health Research)

## Contributors

RACS, JJB, LG, AQ, EK, and DZ contributed equally to the systematic review and are joint first authors. RACS, JJB, DZ, LG, EK, AQ and RB-P were the core team leading the systematic review. JJB, RC, RWMV, SM, YW, ZY, CS, LY, MG and AV-G identified and selected the studies. DZ, EK, RWMV, AA, YW, KH, HP-H, MAH, SLM, AQ and LY collected the data. AQ, LG, BS, GHG, and LT analysed the data. RB-P, HPH, AI, RAM, TD, and DC assessed the certainty of the evidence. SLM, FL, BR, TA, POV, GHG, MM, JDN, ML, BT and GR provided advice at different stages. JJB, RACS, RB-P, and GHG drafted the manuscript. All authors approved the final version of the manuscript. RACS is the guarantor. The corresponding author attests that all listed authors meet authorship criteria and that no others meeting the criteria have been omitted.

## Ethical approval

Not applicable. All the work was developed using published data.

## Data sharing

No additional data available.

RACS affirms that this manuscript is an honest, accurate, and transparent account of the study being reported; that no important aspects of the study have been omitted; and that any discrepancies from the study as planned have been explained.

Dissemination to participants and related patient and public communities: The infographic and MAGICapp decision aids (available at www.magicapp.org/) were created to facilitate conversations between healthcare providers and patients or their surrogates. The MAGICapp decision aids were co-created with people who have lived experience of covid-19.

