## Supplementary File for "Prophylaxis for covid-19: living systematic review and network meta-analysis"

**Supplementary Material**

**Table of contents	Page**

Protocol	1

English search strategy	14

Description of studies excluded at full text stage	20

Additional study characteristics and outcome data	39

Table of reporting differences between versions of study preprints	44

and/or peer-reviewed publications

Risk of bias assessments	45

Network plots by outcome	46

Forest plots for pairwise meta-analyses	47

Complete network meta-analysis results and GRADE	52

Subgroup analyses	53

**Therapies for treatment and prophylaxis of COVID-19: introduction and methods for a living systematic review and network meta-analyses**

An international collaborative project

**Correspondence to**

Reed AC Siemieniuk

**Abstract**

*Objectives*: To compare the effects of therapies for prophylaxis and treatment of COVID-19

*Design:* Living systematic review and network meta-analysis (NMA).

*Data sources:* U.S. Centers for Disease Control and Prevention (CDC) COVID-19 Research Articles Downloadable Database, which includes 25 electronic databases.

*Study selection:* We will include randomized clinical trials (RCT) in which persons exposed to COVID-19 or with suspected, probable or confirmed COVID-19 were treated with pharmaceuticals or blood products aimed at prophylaxis or treatment. Pairs of independent reviewers will screen in duplicate title and abstract and full text of potentially eligible articles.

*Methods:* After duplicate data abstraction, we will conduct a Bayesian-random effects network meta-analysis for each of the outcomes of interest. We will assess the risk of bias of the included studies using a modification of the Cochrane Risk of Bias 2.0 tool, and the certainty of the evidence using the GRADE approach for NMA. We will classify the interventions in groups from the most to the least effective/ harmful following GRADE guidance using a minimally contextualized approach.

*Publication and Updating of Results:* We will publish and update the results in *The BMJ* and magicapp.org. We will update the living NMA when the question is no longer of clinical importance, or new evidence that might impact on the conclusions is unlikely to be forthcoming.

**Background**

COVID-19 is a rapidly evolving global health emergency. As of 27 May 2020, over 5.6 million people have been infected and of these, 335,000 have died,^1^ resulting in an enormous perceived need to implement a possibly effective intervention. Public figures,^2^ guideline bodies,^3^ and government agencies ^4^ have suggested using interventions without established benefit. Clinicians have responded to these suggestions by administering such interventions to large numbers of patients.^5^

With many teams conducting randomized clinical trials (RCTs) of drug interventions—over 1,000 intervention trials registered as of 10 May 2020 ^6^—evidence on the comparative effectiveness of drug interventions will emerge rapidly. The new environment will amplify the need for evidence based medicine: distinguishing trustworthy from untrustworthy evidence, interpreting the results, and judging net benefits of interventions against standard treatment and one another.^7^

Reliable guidance for COVID-19 will require adherence to standards of trustworthy clinical practice guidelines,^8^ including methodically rigorous and rapidly updated evidence summaries. Such summaries will identify interventions with sufficient evidence of net benefit to warrant use. Establishing absence of net benefit in previously highly touted interventions may be equally important. Further, if studies suggest that more than one intervention provides net benefit, clinicians and patients will face the challenge of deciding which intervention to use.

Our living systematic review and network meta-analysis (NMA), updated in real time, will answer this urgent demand for trustworthy evidence on the available therapeutic options. This systematic review is part of the *BMJ Rapid Recommendations* project, a collaborative effort from the MAGIC Evidence Ecosystem Foundation ([www.magicproject.org](http://www.magicproject.org)) and *The BMJ*. Our living NMA will thus directly inform *BMJ Rapid Recommendations*,^9^ providing trustworthy, actionable, and living guidance to clinicians and patients soon after new and potentially practice-changing evidence is made available. The living NMA will be freely available in user-friendly formats, through *The BMJ* and MAGICapp ([www.magicapp.orsg](http://www.magicapp.orsg)), and ready for re-use and adaptation at national and local levels.

**Methods**

**Structure and organization**

The team working in the development of this living systematic review and NMA is composed of the following groups:

a. Oversight group composed of experts in the clinical area and systematic review methodology (RAC, TA, GHG, BR, FL, SM, PV). The role of this team is to ensure that the process follows the highest methodological standards, and that the decisions made are clinically sensible and keep consistency with the needs of the *BMJ Rapid Recommendations*.
b. Core systematic review team leaders. This group is composed of methodologists leading teams in charge of study identification (JB), data abstraction (DZ), data analysis (LG), and assessment of certainty of the evidence and presentation (RBP).
c. Reviewers. This team is composed of clinicians, graduate students, methodologists, and biostatisticians conducting or providing advice for screening, data abstraction, and assessments of certainty of the evidence.

In addition, decisions in this protocol have been made considering the requirements from the *Rapid Recommendations* guideline panel, which includes patients.

**Eligibility criteria**

We will include RCTs in which persons exposed to COVID-19 or with suspected, probable, or confirmed COVID-19 are treated with pharmacologic or blood products aimed at prophylaxis or treatment. We will include trials in which researchers compare any intervention against another or against no intervention, placebo, or standard of care, and report any outcome. We will include trials that report results regardless of publication status (peer-reviewed, in press, or pre-print, but not news reports alone) or language. There will be no restrictions on acuity of disease, nor setting.

We will include trials of pharmaceuticals, blood products, vitamins, minerals and, if the drug is one specific molecule, Chinese medicines. We will exclude quasi-randomized studies and randomized trials evaluating external organ support, plasma exchange, oxygen delivery, ventilation strategies, vaccination, nutrition, traditional Chinese herbal medicines (that typically include more than one molecule or a molecule without specific molecular weighted dosing), exercise/rehabilitation, psychological and educational interventions, personal protective equipment, or any other non-drug supportive care interventions.

**Data sources and searches**

We will search the U.S. Centers for Disease Control and Prevention (CDC) COVID-19 Research Articles Downloadable Database for eligible studies—the most comprehensive database of COVID-19 research articles from December 2019 and which is maintained by the Stephen B. Thacker CDC library ^10^. We will update our search daily Monday to Friday to match the update schedule of the database. The database includes 25 bibliographic and grey literature sources: Medline (Ovid and PubMed), PubMed Central, Embase, CAB Abstracts, Global Health, PsycInfo, Cochrane Library, Scopus, Academic Search Complete, Africa Wide Information, CINAHL, ProQuest Central, SciFinder, the Virtual Health Library, LitCovid, WHO COVID-19 website, CDC COVID-19 website, Eurosurveillance, China CDC Weekly, Homeland Security Digital Library, ClinicalTrials.gov, bioRxiv (preprints), medRxiv (preprints), chemRxiv (preprints), and SSRN (preprints).

We will filter the results from the CDC’s database through a validated and highly sensitive machine learning model to identify RCTs.^11^ We will track preprints of RCTs until publication and data updated to match that in the peer-reviewed publication when discrepant. In addition, we will search six Chinese databases on a biweekly basis: Wanfang, CBM, CNKI, VIP, Chinese Medical Journal Net (preprints), and ChinaXiv (preprints). The search terms for COVID-19 developed by the CDC have been adapted to the Chinese language. For the search of the Chinese literature, we will also include search terms for randomized trials. The search strategy is available in the Supplementary Material.

We will also monitor living evidence retrieval services on an ongoing basis. Two such services are the Systematic and Living Map on COVID-19 Evidence by the Norwegian Institute of Public Health, in collaboration with the Cochrane Canada Centre at McMaster University,^12^ and the Living Overview of Evidence (L·OVE) by Epistemonikos Foundation.^13^ Our publication will be accompanied with a public call for information sharing, with hopes that investigators will initiate contact and share information when available. RCTs may also be identified organically through informal networks.

**Study selection**

Pairs of reviewers will independently and in duplicate screen titles and abstracts followed by full texts. A third reviewer will adjudicate conflicts.

**Data extraction**

For each eligible trial, two reviewers, who have undergone training and have completed calibration exercises, will extract data independently and in duplicate using a standardised, pilot-tested data extraction form. Discrepancies will be resolved by discussion, and when necessary adjudicated by a third person. The study characteristics and baseline participant information that will be extracted is presented in Box 1.

**Box 1.** Study characteristics and baseline participant information that will be extracted

- Geographic location
- Physical location (outpatient, inpatient, intensive care)
- Patient and public involvement in the study design or interpretation
- Funder (public, private)
- Number randomized
- Number randomized to each intervention
- Dose, frequency, route of administration, and duration for each intervention
- Number who received each intervention
- Mean age
- Percent male
- Severity of illness (non-severe, severe, critically ill)
- Mean oxygen saturation on room air, or mean baseline amount of supplemental oxygen
- Percent receiving mechanical ventilation at baseline
- Percent current and former smokers
- Percent with hypertension
- Percent with underlying chronic respiratory condition including chronic obstructive pulmonary disease (COPD), asthma, and others
- Percent with diabetes
- Percent taking angiotension-converting-enzyme (ACE) inhibitors or angiotensin receptor blockers (ARBs)
- Mean alanine aminotransferase (ALT)
- Mean C-reactive protein (CRP)
- Mean d-dimer
- Mean lactate dehydrogenase (LDH)
- Mean lymphocyte count, and percent with lymphopaenia

**Outcomes**

We will begin by focusing on the patient-important outcomes listed below, based on the WHO’s candidate core outcome set.^14^ The list of outcomes may be modified at the discretion of the BMJ *Rapid Recommendations* standing panel of experts, which includes frontline healthcare workers and patient-partners. As harms are typically specific to individual pharmacologic therapies, these outcomes will be selected at the time they are needed to support specific linked Rapid Recommendations. We will extract the outcome data closest to the prespecified outcome time point.

The initial outcomes examined for the studies of treatment will include:

- Mortality (time frame: closest to 90 days)
- Mechanical ventilation in patients not initially mechanically ventilated (time frame: closest to 90 days)
- Duration of hospitalization
- Admission to hospital (time frame: closest to 28 days)
- Adverse effects leading to discontinuation of the intervention (time frame: closest to 28 days)
- Time to symptom resolution
- Time until the amount of SARS-CoV2 viral particles is below the threshold set by the RCT authors for being likely no longer infectious (as a surrogate for transmissibility)
- Undetectable nCoV-19 by PCR (time frame: closest to 7 days and not less than 4 days or more than 10 days)

The initial outcomes examined for the studies of prophylaxis will include:

- Symptomatic SARS-CoV2 infection (time frame: closest to 28 days)
- Mortality (time frame: closest to 90 days)
- Admission to hospital (time frame: closest to 28 days)
- Adverse effects leading to discontinuation of the intervention (time frame: closest to 28 days)
- Time to symptom resolution (those not infected will be considered 0)

Other outcomes that will be extracted and reported from the trials, but not initially reviewed will include:

- Ventilator-free days (time frame: 28 days)
- Venous thromboembolism (time frame: closest to 90 days)
- Clinically important bleeding (time frame: closest to 90 days)

**Risk of bias assessments**

Following calibration exercises, two reviewers, working independently and in duplicate, will use the Clinical Advances Through Research and Information Translation (CLARITY) revisions of the Cochrane tool for assessing risk of bias in randomized trials (RoB 2.0) to rate trials as either ‘low risk of bias’, ‘some concerns – probably low risk of bias’, ‘some concerns – probably high risk of bias’ and ‘high risk of bias’, across the following domains: bias arising from the randomization process, bias due to departures from the intended intervention, bias due to missing outcome data, bias in measurement of the outcome, bias in selection of the reported results.^15 16^ The response choice ‘some concerns’ was modified to include a judgement about whether the concerns probably do or do not result in high risk of bias.^16^ We will not consider judgements about applicability when considering risk of bias because applicability is judged separately within indirectness domain of the GRADE framework.^17^ In addition, the modified tool includes an additional domain to assess risk of bias related to RCTs stopping early for benefit.^18^ Reviewers will resolve discrepancies by discussion, and when not possible, adjudication by a third-party research methodologist. A detailed guide for our risk of bias assessments is available in the Supplementary Material. If the body of evidence is rated as high risk of bias for missing data only (i.e., no serious concerns with any other of the risk of bias domains), we may perform sensitivity analyses with worst plausible assumptions to see if results remain robust to missing data.

**Treatment nodes**

We will perform three separate network meta-analyses: i) pharmacologic treatments for patients with suspected or confirmed COVID-19, ii) blood products for treatment of patients with suspected or confirmed COVID-19, and iii) pharmacologic prophylactic therapy for people exposed to COVID-19.

Treatments will be grouped into common nodes based on molecule but not dose or duration: we will include all doses and durations of the same medication in a single treatment node. When an intervention includes more than one medication, it will be included as a separate node. We will include drugs from the same class within the same node. Chloroquine and hydroxychloroquine will be included in the same node for COVID-19 specific effects and separated for disease-independent adverse effects. Prespecified nodes include angiotensin-converting enzyme inhibitors and angiotensin receptor blockers, anti-interleukin-6 agents (i.e., tocilizumab, sarilumab), glucocorticoids, interferons, JAK inhibitors, statins, antiplatelet agents, and anticoagulants. Antibiotics will be grouped into nodes by class (e.g., macrolides, beta-lactams). The NMA oversight group and the linked Rapid Recommendation panels will make final decisions on how to group treatment nodes.

**Data synthesis**

We will perform random-effects pairwise meta-analysis for each comparison for each outcome using the Bayesian framework. We will use a plausible prior for variance parameter,^19^ and uniform prior for the effect parameter. We will calculate Ratio of Means (RoM) and corresponding 95% credible intervals (CrIs) for continuous outcomes in which we expect variationa cross populations (time to symptoms resolution and time to viral clearance) and mean differences (MDs) and corresponding 95% credible intervals (CrIs) for other continuous outcomes. For dichotomous outcomes, we will calculate odds ratios (ORs) with corresponding CrIs. Absolute effects will be calculated based on the ORs and baseline risk of standard of care.^20^ We will create a funnel plot to assess the publication bias when 10 or more studies are available for a specific direct comparison.^21^

We will conduct a random-effects network meta-analysis using the Bayesian framework with same priors for the variance and effect parameters.^19^ We will use three Markov-chains with 100,000 iterations after an initial burn-in of 10,000 and a thinning of 10. We will assess the convergence based on trace plots and the Brooks-Gelman-Rubin statistic, with an acceptable threshold of <1.05 for all nodes. If convergence is not achieved, then we will use 500,000 iterations and a burn-in of 50,000 iterations and a thinning of 10. We will use automated generation of node-splitting models to assess local incoherence and to obtain indirect estimates.^22^ We will estimate ranking probabilities and calculate surface under the cumulative ranking curves (SUCRA). For all dichotomous outcomes, we will calculate the absolute treatment effects of the network estimates based-on the odds ratios and the baseline risk using the transitive risks model.^23^ To obtain the baseline risk, we will use the highest quality prognostic evidence available at the time of publication, typically based on a systematic review of prognostic studies.^24^

When networks are sparse, between-study heterogeneity variances are often imprecisely estimated. That may generate implausibly wide credible intervals from network estimates, even when the direct and indirect estimates are coherent.^25^ When this occurs, we will conduct sensitivity analyses by using empirically informative priors,^26^ and fixed effects models.^25^ Another situation in which implausibly wide confidence intervals occur is with random effects when the number of studies is small and the heterogeneity is considerable, particularly when study size varies markedly. This is a second situation in which we may opt for fixed effect models.

Pairwise meta-analysis will be conducted using the bayesmeta package of R version 4.0.0 (RStudio, Boston, MA).^27^ All network meta-analyses will be performed using the *gemtc* package of R version 4.0.0 (RStudio, Boston, MA),^28^ absolute effects in networks will be calculated using *R2jags* package of R version 4.0.0 (RStudio, Boston, MA)^29^. *networkplot* command of Stata version 15.1 (StataCorp, College Station, Texas, USA) will be used to draw the network plots with thickness of edges of the nodes based on inverse variance.^30^ The foundational R code that we will use is available in the Supplementary Material.

**Subgroups and sensitivity analyses**

We will conduct sensitivity analyses and network meta-regression to explore factors that may modify the comparative effect estimates. Specific analyses will be guided by the linked *Rapid Recommendation* guideline panels with the directive to explore a limited number of pre-defined subgroup effects with a specified rationale and anticipated direction of the effect. When sufficient heterogeneity in risk of bias exists, we will perform subgroup analyses comparing studies at low versus high risk of bias. We will classify all studies with at least 1 domain judged at high risk of bias or probably high risk of bias as a high risk of bias study. Ultimately, we will base decisions regarding the credibility of subgroup analyses on the ICEMAN instrument,^31^ and follow GRADE guidance on how to deal with low, intermediate, and high credibility subgroup effects.^32^

For all outcomes, we will first include both peer-reviewed and non-peer reviewed data, and then perform a sensitivity analysis by restricting to peer-reviewed publications. When the guideline panel makes a recommendation about a drug class with more than one drug, we will consider performing a sensitivity analysis with individual molecules rather than drug class as the nodes.

**Certainty assessment**

We will evaluate the certainty of evidence using the GRADE approach for network meta-analysis.^33-35^ We will rate the certainty for each comparison and outcome as high, moderate, low, or very low, based on considerations of risk of bias, inconsistency, indirectness, publication bias, intransitivity, incoherence, and imprecision. If and when sufficient effective treatments are available we will, for each outcome, use the GRADE framework to make conclusions by classifying the interventions in groups from the most to the least effective using a minimally contextualized approach.^36^

**Updating**

Full results will be updated at minimum every two weeks on webpages hosted by BMJ.com and magicapp.org. A summary of the interim results, risk of bias assessments, individual study data summaries, and the date of the search will be posted in a table within the online publication. Results will be updated more frequently if the project steering committee or the linked BMJ Rapid Recommendations guidelines panel judges that there is sufficient new information to possibly change practice and requests an earlier update. Updates will be submitted to international indexes through the mechanism for corrections but labelled as updates. Old versions and details of any changes will be dated and documented in the Supplementary Material. It may be necessary to adjust some of the processes, outcomes, and analytic methods based on the data. The systematic review team will decide together whether changes to the pre-specified protocol are needed and these changes will be documented in the methods section of the updated review.

**External review**

A standing peer-review committee editorial staff at The BMJ, clinical experts, patients, and statistician(s) will be tasked with providing feedback on the protocol and initial publication, and a subcommittee will peer review updates. The systematic review team will respond to these comments on an urgent basis. Comments from peer reviewers and responses will be made public. The study webpage will include functionality for comments to be made by any member of the public. The systematic review team will endeavor to respond to all public comments in a timely manner.

**Publication**

The papers will be published in a traditional format for systematic reviews and network meta-analyses. In addition, results will be published in interactive evidence summary and decision aid formats for multiple comparisons and pairwise comparisons on MAGICapp ([www.magicapp.org](http://www.magicapp.org)). The publication will include infographics that highlight the key messages from the current state of the evidence.

**Determining the end of project**

The project will terminate at the discretion of the project oversight group, which includes representation from the MAGIC Evidence Ecosystem Foundation, The BMJ, the WHO, and international collaborators with clinical and research expertise. We anticipate that the project will end when the question is no longer of clinical importance, or new evidence that might impact on the conclusions is unlikely to be forthcoming. If the influx of new RCT results slows, then the project oversight committee may choose to decrease the frequency of updates.

**Data access**

All extracted data will be made available publicly at the time of the updates to be shared and used freely.

**Changes to the protocol**

All changes to the protocol are reported in Table 1 and will be updated as necessary.

**Discussion**

Our living network meta-analysis will provide up to date information to interested users, including healthcare workers, patients, healthcare agencies, guideline bodies, and governments. It will directly inform joint clinical practice guidelines from BMJ Rapid Recommendations.

We anticipate several challenges. First, living systematic reviews require substantial dedication and human resources.^37^ To address this, we will organize our large team into smaller groups with specific responsibilities, including i) a study identification team, ii) a data extraction and management team, iii), a data analysis team, iv) a grading and publications team, and v) an oversight team. In addition, *The BMJ* will recruit a standing external review committee that will become familiar with the research methods and respond quickly to updates.

Second, the need for recurrent updates conflicts with traditional publication formats. Frequent updates resulting in new publications can have a devastating effect on a journal’s impact factor. It may also be confusing for users who want to find the most recent update. To solve this problem, we plan to i) have a reference website with the links to the latest versions, ii) use headers on previous versions stating that they are outdated with a link to the current version, iii) use PubMed’s corrections mechanism so that the updated publication keeps the same digital object identifier (DOI) and PubMed identification number.

Evidence summaries will also be published online with MAGICapp ([www.magicapp.org](http://www.magicapp.org)) developed for the purpose of dynamic updating and with electronic publication formats not subject to the limitations of traditional publishing formats thus allowing presentation of evidence summaries in multilayered user-friendly formats.^38^

Third, the optimal methodological and statistical methods may change as the number of RCTs and participants increases. In particular, early NMAs are likely to be sparse, and we may have to use specific analytic methods to accommodate these situations.^25^ Our oversight committee and semi-independent guideline panels will provide direction and guidance on whether and how the methods should be adopted to the current situation.

Another major concern with living systematic reviews is limitations in the evidence included, in particular bias. We will assess risk of bias of each study using a standardized tool,^15 16^ and assess risk of bias for each comparison using the GRADE framework.^33 34^ Publication bias is of particular concern with living reviews: studies with positive results are more likely to be published at all, and when published are likely to be published earlier.^39^ We will use the GRADE approach, which considers publication bias, to rate certainty for the estimated effect for each comparison.^21^ Clinical practice guideline panels of whom the majority will be independent of this review, will consider this issue when making recommendations for practice, and their scrutiny may bear on certainty of evidence judgements.^9^

There are at least two other groups planning a similar living systematic review and NMA.^40 41^ More than one group performing similar analyses will allow controlled replication and the scientific community to assess reproducibility of the findings. Our review has advantages over the alternatives, including the use of and experience with GRADE for NMA, an established platform in the *BMJ Rapid Recommendations* project; an established collaboration with and early involvement of the publisher (*The* *BMJ*); and interpretation from a semi-independent *BMJ Rapid Recommendations* guideline panels; inclusion of a dedicated search of the Chinese literature; and co-publication in MAGICapp through its interactive multilayered evidence presentation format.

**Conclusions**

The COVID-19 pandemic necessitates rapid interpretation of new evidence addressing therapeutic and prophylactic options. Our living systematic review and NMA will provide a reference point for those interested in the most trustworthy evidence regarding these therapies. Our project will facilitate the movement of evidence into practice much sooner than traditional publication methods.

**Table 1**. Changes to the protocol

| Date | Change | Rationale |
| --- | --- | --- |
| 15 Jul 2020 | Network analyses will only include treatment nodes with at least 100 patients or at least 20 events. | Early analyses with sparse data resulted in implausible and uninformative effect estimates. |
| 28 Aug 2020 | Chinese databases searches will be completed monthly. | Balancing feasibility with search result yield. |

4. U.S. Food and Drug Administration. Request for Emergency Use Authorization For Use of Chloroquine Phosphate or

Hydroxychloroquine Sulfate Supplied From the Strategic National Stockpile for Treatment

of 2019 Coronavirus Disease. In: (HHS) USDoHaHS, ed., 2020:1-8.

5. Kim AHJ, Sparks JA, Liew JW, et al. A Rush to Judgment? Rapid Reporting and Dissemination of Results and Its Consequences Regarding the Use of Hydroxychloroquine for COVID-19. *Ann Intern Med* 2020 doi: 10.7326/m20-1223 [published Online First: 2020/04/01]

6. Cytel. Global Coronavirus COVID-19 Clinical Trial Tracker 2020 [Available from: <https://www.covid19-trials.org/> accessed 6 MAy 2020.

7. World Health Organization. SUBJECT IN FOCUS: providing timely and accurate information to dispel the ‘infodemic’. Coronavirus disease 2019 (COVID-19) Situation Report. Geneva, Switzerland, 2020.

8. Institute of Medicine. Institute of Medicine (US) Committee on Standards for Developing Trustworthy Clinical Practice Guidelines. Washington, DC: National Academy Press (US) 2011.

19. Röver C. Bayesian random-effects meta-analysis using the bayesmeta R package. *arXiv preprint arXiv:171108683* 2017

**CDC Search Strategy for English Databases**

| **Database** | **Strategy** |
| --- | --- |
| Medline (Ovid) 1946- | (coronavir* OR corona virus* OR betacoronavir* OR covid19 OR covid 19 OR nCoV OR novel CoV OR CoV 2 OR CoV2 OR sarscov2 OR 2019nCoV OR wuhan virus*).mp. OR ((wuhan OR hubei OR huanan) AND (severe acute respiratory OR pneumonia*) AND outbreak*).mp. OR Coronavirus Infections/ OR Coronavirus/ OR betacoronavirus/  Limits: 2020- OR  (novel coronavir* OR novel corona virus* OR covid19 OR covid 19 OR nCoV OR novel CoV OR CoV 2 OR CoV2 OR sarscov2 OR 2019nCoV OR wuhan virus*).mp. OR ((wuhan OR hubei OR huanan) AND (severe acute respiratory OR pneumonia*) AND outbreak*).mp. OR ((wuhan OR hubei OR huanan) AND (coronavir* OR betacoronavir*)).mp.  Limits: 2019- |
| Embase (Ovid) 1947- | (coronavir* OR corona virus* OR betacoronavir* OR covid19 OR covid 19 OR nCoV OR novel CoV OR CoV 2 OR CoV2 OR sarscov2 OR 2019nCoV OR wuhan virus*).mp. OR ((wuhan OR hubei OR huanan) AND (severe acute respiratory OR pneumonia*) AND outbreak*).mp. OR Coronavirus infection/ OR coronavirinae/ OR exp betacoronavirus/  Limits: 2020- OR  (novel coronavir* OR novel corona virus* OR covid19 OR covid 19 OR nCoV OR novel CoV OR CoV 2 OR CoV2 OR sarscov2 OR 2019nCoV OR wuhan virus*).mp. OR ((wuhan OR hubei OR huanan) AND (severe acute respiratory OR pneumonia*) AND outbreak*).mp. OR ((wuhan OR hubei OR huanan) AND (coronavir* OR betacoronavir*)).mp.  Limits: 2019- |
| CAB  Abstracts (Ovid) 1910- | (coronavir* OR corona virus* OR betacoronavir* OR covid19 OR covid 19 OR nCoV OR novel CoV OR CoV 2 OR CoV2 OR sarscov2 OR 2019nCoV OR wuhan virus*).mp. OR ((wuhan OR |

|  | hubei OR huanan) AND (severe acute respiratory OR pneumonia*) AND outbreak*).mp. OR exp Betacoronavirus/  Limits: 2020- OR  (novel coronavir* OR novel corona virus* OR covid19 OR covid 19 OR nCoV OR novel CoV OR CoV 2 OR CoV2 OR sarscov2 OR 2019nCoV OR wuhan virus*).mp. OR ((wuhan OR hubei OR huanan) AND (severe acute respiratory OR pneumonia*) AND outbreak*).mp. OR ((wuhan OR hubei OR huanan) AND (coronavir* OR betacoronavir*)).mp.  Limits: 2019- |
| --- | --- |
| Global Health (Ovid) 1910- | (coronavir* OR corona virus* OR betacoronavir* OR covid19 OR covid 19 OR nCoV OR novel CoV OR CoV 2 OR CoV2 OR sarscov2 OR 2019nCoV OR wuhan virus*).mp. OR ((wuhan OR hubei OR huanan) AND (severe acute respiratory OR pneumonia*) AND outbreak*).mp. OR exp Betacoronavirus/  Limits: 2020- OR  (novel coronavir* OR novel corona virus* OR covid19 OR covid 19 OR nCoV OR novel CoV OR CoV 2 OR CoV2 OR sarscov2 OR 2019nCoV OR wuhan virus*).mp. OR ((wuhan OR hubei OR huanan) AND (severe acute respiratory OR pneumonia*) AND outbreak*).mp. OR ((wuhan OR hubei OR huanan) AND (coronavir* OR betacoronavir*)).mp.  Limits: 2019- |
| PsycInfo (Ovid) 1806- | (coronavir* OR corona virus* OR betacoronavir* OR covid19 OR covid 19 OR nCoV OR novel CoV OR CoV 2 OR CoV2 OR sarscov2 OR 2019nCoV OR wuhan virus*).mp. OR ((wuhan OR hubei OR huanan) AND (severe acute respiratory OR pneumonia*) AND outbreak*).mp.  Limits: 2020- OR  (novel coronavir* OR novel corona virus* OR covid19 OR covid 19 OR nCoV OR novel CoV OR CoV 2 OR CoV2 OR sarscov2 OR 2019nCoV OR wuhan virus*).mp. OR ((wuhan OR hubei OR huanan) AND (severe acute respiratory OR pneumonia*) AND outbreak*).mp. OR ((wuhan OR hubei OR huanan) AND (coronavir* OR betacoronavir*)).mp.  Limits: 2019- |

| Cochrane Library | #1(coronavir* OR "corona virus" OR betacoronavir* OR covid19 OR "covid 19" OR nCoV OR "CoV 2" OR CoV2 OR sarscov2 OR 2019nCoV OR "novel CoV" OR "wuhan virus"):ti,ab,kw OR ((wuhan OR hubei OR huanan) AND ("severe acute respiratory"  OR pneumonia*) AND outbreak*):ti,ab,kw #2MeSH descriptor: [Coronavirus] this term only  #3MeSH descriptor: [Coronavirus Infections] this term only #4MeSH descriptor: [Betacoronavirus] this term only  #5 #1 OR #2 OR #3 OR#4  Limits: 2020- OR  #1 ( "novel coronavirus" OR "novel corona virus" OR covid19 OR "covid 19" OR nCoV OR "novel CoV" OR "CoV 2" OR CoV2 OR sarscov2 OR 2019nCoV OR "wuhan virus"):ti,ab,kw OR ((wuhan OR hubei OR huanan) AND ("severe acute respiratory" OR pneumonia*) AND outbreak*):ti,ab,kw OR ((wuhan OR hubei OR huanan) AND (coronavir* OR betacoronavir*)):ti,ab,kw  Limits: 2019- |
| --- | --- |
| Scopus 1960- | TITLE-ABS-KEY ( coronavir* OR "corona virus" OR betacoronavir* OR covid19 OR "covid 19" OR ncov OR "CoV 2" OR cov2 OR sarscov2 OR 2019ncov OR "novel CoV" OR "wuhan virus" ) OR ( TITLE-ABS-KEY ( wuhan OR hubei OR huanan ) AND TITLE-ABS-KEY ( "severe acute respiratory" OR pneumonia* ) AND TITLE-ABS-KEY ( outbreak* ) ) AND ( LIMIT-TO ( PUBYEAR , 2020 ) )  OR  TITLE-ABS-KEY ( "novel coronavirus" OR "novel corona virus" OR covid19 OR "covid 19" OR ncov OR "CoV 2" OR cov2 OR sarscov2 OR 2019ncov OR "novel CoV" OR "wuhan virus" ) OR TITLE-ABS-KEY ( ( wuhan OR hubei OR huanan ) AND ( "severe acute respiratory" OR pneumonia* ) AND outbreak* ) OR TITLE-ABS-KEY ( ( wuhan OR hubei OR huanan ) AND ( coronavir* OR betacoronavir* ) ) AND ( LIMIT-TO ( PUBYEAR , 2020 ) OR LIMIT-TO ( PUBYEAR , 2019 ) ) |
| Academic Search Complete (Ebsco) | TI,AB,SU( (coronavir* OR "corona virus" OR betacoronavir* OR covid19 OR "covid 19" OR nCoV OR "CoV 2" OR CoV2 OR sarscov2 OR 2019nCoV OR "novel CoV" OR "wuhan virus") OR ((wuhan OR hubei OR huanan) AND ("severe acute respiratory" OR pneumonia*) AND (outbreak*)) )  Limits: Dec. 2019-, peer-reviewed |
| Africa Wide Information (Ebsco) | TI,AB,SU( (coronavir* OR "corona virus" OR betacoronavir* OR covid19 OR "covid 19" OR nCoV OR "CoV 2" OR CoV2 OR sarscov2 OR 2019nCoV OR "novel CoV" OR "wuhan virus") OR |

|  | ((wuhan OR hubei OR huanan) AND ("severe acute respiratory" OR pneumonia*) AND (outbreak*)) )  Limits: 2020-, peer-reviewed OR  TI,AB,SU( ("novel coronavirus" OR "novel corona virus" OR covid19 OR "covid 19" OR nCoV OR "CoV 2" OR CoV2 OR sarscov2 OR 2019nCoV OR "novel CoV" OR "wuhan virus") OR ((wuhan OR hubei OR huanan) AND ("severe acute respiratory" OR pneumonia*) AND outbreak*) OR ((wuhan OR hubei OR huanan) AND (coronavir* OR betacoronavir*)) )  Limits: 2019-, peer-reviewed |
| --- | --- |
| CINAHL  (Ebsco) | TI,AB,SU( (coronavir* OR "corona virus" OR betacoronavir* OR covid19 OR "covid 19" OR nCoV OR "CoV 2" OR CoV2 OR sarscov2 OR 2019nCoV OR "novel CoV" OR "wuhan virus") OR ((wuhan OR hubei OR huanan) AND ("severe acute respiratory" OR pneumonia*) AND (outbreak*)) ) OR (MH "Coronavirus") OR (MH "Coronavirus Infections")  Limits: Dec. 2019-, peer-reviewed |
| ProQuest Central (Proquest) 1952- | TI,AB,SU( (coronavir* OR "corona virus" OR betacoronavir* OR covid19 OR "covid 19" OR nCoV OR "CoV 2" OR CoV2 OR sarscov2 OR 2019nCoV OR "novel CoV" OR "wuhan virus") OR ((wuhan OR hubei OR huanan) AND ("severe acute respiratory" OR pneumonia*) AND (outbreak*)) )  Limits: Dec. 2019-, peer-reviewed |
| PubMed Central | TITLE-ABSTRACT( (coronavirus OR "corona virus" OR coronavirinae OR coronaviridae OR betacoronavirus OR covid19 OR "covid 19" OR nCoV OR "CoV 2" OR CoV2 OR sarscov2 OR 2019nCoV OR "novel CoV" OR "wuhan virus" ) OR ((wuhan OR hubei OR huanan) AND ( "severe acute respiratory" OR pneumonia ) AND (outbreak)) ) OR "COVID-19" [Supplementary Concept] OR "severe acute respiratory syndrome coronavirus 2" [Supplementary Concept]  Limits: Dec. 2019- |
| Medline (PubMed) | TITLE-ABSTRACT( ( coronavirus OR "corona virus" OR coronavirinae OR coronaviridae OR betacoronavirus OR covid19 OR "covid 19" OR nCoV OR "CoV 2" OR CoV2 OR sarscov2 OR 2019nCoV OR "novel CoV" OR "wuhan virus" ) OR ((wuhan OR hubei OR huanan) AND ( "severe acute respiratory" OR pneumonia ) AND (outbreak)) ) OR "COVID-19" [Supplementary Concept] OR "severe acute respiratory syndrome coronavirus 2" [Supplementary Concept]  Limits: Dec. 2019- |

| LitCovid (NLM) | <https://www.ncbi.nlm.nih.gov/research/coronavirus/> |
| --- | --- |
| SciFinder (CAS) | References ( coronavir* OR "corona virus" OR betacoronavir* OR covid19 OR covid OR nCoV OR "CoV 2" OR CoV2 OR sarscov2 OR 2019nCoV OR "novel CoV" OR "wuhan virus" )  Limits: 2020- OR  References ( "novel coronavirus" OR "novel corona virus" OR covid19 OR covid OR nCoV OR "CoV 2" OR CoV2 OR sarscov2 OR 2019nCoV OR "novel CoV" OR "wuhan virus" )  Limits: 2019- |
| Virtual Health Library (WHO) | Filter VHL created: <https://bvsalud.org/vitrinas/post_vitrines/novo_coronavirus/> **Database changed on 6/16/2020 and integrated into the larger WHO database**  Limited: 2019- OR  TI,AB:( (coronavirus OR "corona virus" OR coronavirinae OR coronaviridae OR betacoronavirus OR covid19 OR "covid 19" OR nCoV OR "CoV 2" OR CoV2 OR sarscov2 OR 2019nCoV OR "novel CoV" OR "wuhan virus" ) OR ((wuhan OR hubei OR huanan) AND ("severe acute respiratory" OR pneumonia) AND (outbreak)) )  Limits: 2020- OR  TI,AB( "novel coronavirus" OR "novel corona virus" OR covid19 OR "covid 19" OR nCoV OR "CoV 2" OR CoV2 OR sarscov2 OR 2019nCoV OR "novel CoV" OR "wuhan virus") OR ((wuhan OR hubei OR huanan) AND ("severe acute respiratory" OR pneumonia) AND outbreak) OR ((wuhan OR hubei OR huanan) AND (coronavirus OR betacoronavirus))  Limits: 2019- |
| [WHO Novel](https://www.who.int/emergencies/diseases/novel-coronavirus-2019) [Coronavirus](https://www.who.int/emergencies/diseases/novel-coronavirus-2019) [page](https://www.who.int/emergencies/diseases/novel-coronavirus-2019) | Download of their global research on COVID 19 database: [https://www.who.int/emergencies/diseases/novel-coronavirus-](https://www.who.int/emergencies/diseases/novel-coronavirus-2019/global-research-on-novel-coronavirus-2019-ncov) [2019/global-research-on-novel-coronavirus-2019-ncov](https://www.who.int/emergencies/diseases/novel-coronavirus-2019/global-research-on-novel-coronavirus-2019-ncov) |

|  | OR  Hand search |
| --- | --- |
| [CDC Novel](https://www.cdc.gov/coronavirus/2019-ncov/index.html) [Coronavirus](https://www.cdc.gov/coronavirus/2019-ncov/index.html) [page](https://www.cdc.gov/coronavirus/2019-ncov/index.html) | <https://www.cdc.gov/coronavirus/2019-ncov/publications.html> OR  Hand search |
| EuroSurveill ance | [https://www.eurosurveillance.org/content/2019-](https://www.eurosurveillance.org/content/2019-ncov?pageSize=100&page=1) [ncov?pageSize=100&page=1](https://www.eurosurveillance.org/content/2019-ncov?pageSize=100&page=1) |
| [China CDC](http://weekly.chinacdc.cn/) [MMWR](http://weekly.chinacdc.cn/) | Hand search |
| Homeland Security Digital Library | Title or Summary: Coronavirus OR “Corona virus” OR Betacoronavirus OR Coronaviridae OR coronavirinae OR Covid OR Covid19 OR nCoV OR CoV OR CoV2 OR Wuhan  Limits: 2019- |
| ClinicalTrials | Condition, disease, other term: Coronavirus OR “Corona virus” OR Betacoronavirus OR Coronaviridae OR coronavirinae OR Covid OR Covid19 OR nCoV OR CoV OR CoV2 OR Wuhan  Limits: 2019- |
| [bioRxiv](https://www.biorxiv.org/) [medRxiv](https://www.medrxiv.org/) [chemRxiv](https://chemrxiv.org/) (preprints) | Condition, disease, other term: Coronavirus OR “Corona virus” OR Betacoronavirus OR Coronaviridae OR coronavirinae OR Covid OR Covid19 OR nCoV OR CoV OR CoV2 OR Wuhan  Limits: 2019- |
| [SSRN](https://www.ssrn.com/index.cfm/en/)  (preprints) | Condition, disease, other term: Coronavirus OR “Corona virus” OR Betacoronavirus OR Coronaviridae OR coronavirinae OR Covid OR Covid19 OR nCoV OR CoV OR CoV2 OR Wuhan  Limits: 2019- |

**Table of studies excluded at full text screening stage (n=463)**

References available upon request

| **Study** | **Title** | **Exclusion Reason** |
| --- | --- | --- |
| Abbaspour Kasgari 2020 | Evaluation of the efficacy of sofosbuvir plus daclatasvir in combination with ribavirin for hospitalized COVID-19 patients with moderate disease compared with standard care: a single-centre, randomized controlled trial | Drug treatment |
| Abd-Elsalam 2020 | Hydroxychloroquine in the Treatment of COVID-19: A Multicenter Randomized Controlled Study | Drug treatment |
| Abd-Elsalam 2020 | Do Zinc Supplements Enhance the Clinical Efficacy of Hydroxychloroquine?: a Randomized, Multicenter Trial | Drug treatment |
| Abdelalim 2021 | Corticosteroid nasal spray for recovery of smell sensation in COVID-19 patients: A randomized controlled trial | Drug treatment |
| Abdelmaksoud 2021 | Olfactory disturbances as presenting manifestation among Egyptian patients with COVID-19: Possible role of zinc | Drug treatment |
| Abolghasemi 2020 | Clinical efficacy of convalescent plasma for treatment of COVID-19 infections: Results of a multicenter clinical study | Not a randomized trial |
| Agarwal 2020 | Convalescent plasma in the management of moderate COVID-19 in India: An open-label parallel-arm phase II multicentre randomized controlled trial (PLACID Trial) | Blood product |
| Agarwal 2020 | Convalescent plasma in the management of moderate COVID-19 in India: An open-label parallel-arm phase II multicentre randomized controlled trial (PLACID Trial) | Blood product |
| Agarwal 2020 | Convalescent plasma in the management of moderate COVID-19 in India: An open-label parallel-arm phase II multicentre randomized controlled trial (PLACID Trial) | Blood product |
| Agarwal 2020 | Convalescent plasma in the management of moderate COVID-19 in India: An open-label parallel-arm phase II multicentre randomized controlled trial (PLACID Trial) | Blood product |
| Ahmed 2020 | A five day course of ivermectin for the treatment of COVID-19 may reduce the duration of illness | Drug treatment |
| Ai 2020 | Effect of Integrated Traditional Chinese and Western Medicine on T Lymphocyte Subsets of Patients with Normal Type of COVID-19 | Traditional Chinese medicine |
| Ai 2020 | Therapeutic effect of integrated traditional Chinese and western medicine on COVID-19 in Guangzhou | Traditional Chinese medicine |
| Al Qahtan 2020 | Randomized controlled trial of convalescent plasma therapy against standard therapy in 2 patients with severe COVID-19 disease | Blood product |
| Al-Sadawi 2020 | Hydroxychloroquine and Azithromycin Usage in African American Patients With Coronavirus Disease 2019 (COVID-19) and Their Effects on QT Interval | Not a randomized trial |
| Alabama 2020 | Trial of Hydroxychloroquine In Covid-19 Kinetics | Randomized trial with no results (e.g. trial registry) |
| Alamdari 2020 | Application of methylene blue -vitamin C - N-acetyl cysteine for treatment of critically ill COVID-19 patients, report of a phase-I clinical trial. (Special Issue: Therapeutic targets and pharmacological treatment of COVID-19.) | Not a randomized trial |
| Alsan 2020 | Comparison of Knowledge and Information-Seeking Behavior After General COVID-19 Public Health Messages and Messages Tailored for Black and Latinx Communities : A Randomized Controlled Trial. | Not exposed to or diagnosed with (confirmed or suspected) COVID-19 |
| AlShehry 2021 | Safety and Efficacy of Convalescent Plasma for Severe COVID-19: Interim Report of a Multicenter Phase II Study from Saudi Arabia | Not a randomized trial |
| Altay 2020 | Combined metabolic cofactor supplementation accelerates recovery in mild-to- moderate COVID-19 | Drug treatment |
| Anderson 2020 | Safety and Immunogenicity of SARS-CoV-2 mRNA-1273 Vaccine in Older Adults | Vaccine |
| Angus 2020 | Effect of Hydrocortisone on Mortality and Organ Support in Patients With Severe COVID-19 The REMAP-CAP COVID-19 Corticosteroid Domain Randomized Clinical Trial | Drug treatment |
| Ankrum 2020 | Can cell therapies halt cytokine storm in severe COVID-19 patients? | Not a randomized trial |
| Ansarin 2020 | Effect of bromhexine on clinical outcomes and mortality in COVID-19 patients: A randomized clinical trial | Drug treatment |
| Araimo 2020 | Ozone as Adjuvant Support in the Treatment of COVID-19: A Preliminary Report of Probiozovid Trial | Oxygen delivery |
| Avendaño-Solà 2020 | Convalescent Plasma for COVID-19: A multicenter, randomized clinical trial | Blood product |
| Axfors 2020 | Mortality outcomes with hydroxychloroquine and chloroquine 2 in COVID-19: an international collaborative meta-analysis of 3 randomized trials | Drug treatment |
| Ayerdi 2020 | Preventive efficacy of tenofovir/emtricitabine against SARS-CoV-2 among PREP users | Not a randomized trial |
| Azhar 2020 | Hydroxychloroquine, Oseltamivir and Azithromycin for the Treatment of COVID-19 Infection: An RCT | Randomized trial with no results (e.g. trial registry) |
| Babalola 2021 | Ivermectin shows clinical benefits in mild to moderate Covid19 disease: A randomised controlled double blind dose response study in Lagos. | Drug treatment |
| Baden 2020 | Efficacy and Safety of the mRNA-1273 SARS-CoV-2 Vaccine. | Vaccine |
| Bajpai 2020 | Efficacy of Convalescent Plasma Therapy compared to Fresh Frozen Plasma in Severely ill COVID-19 Patients: A Pilot Randomized Controlled Trial. | Blood product |
| Balcells 2020 | Early Anti-SARS-CoV-2 Convalescent Plasma in Patients Admitted for COVID- 19: A Randomized Phase II Clinical Trial | Blood product |
| Bandopadhyay 2020 | Nature and dimensions of the cytokine storm and its attenuation by convalescent plasma in severe COVID-19 | Blood product |
| Bao 2020 | Successful treatment of patients severely ill with COVID-19 | Not a randomized trial |
| Bautin 2020 | Inhalation surfactant therapy in the integrated treatment of severe COVID-19 pneumonia | Not a randomized trial |
| Beigel 2020 | Remdesivir for the Treatment of Covid-19 — Preliminary Report | Drug treatment |
| Beigel 2020 | Remdesivir for the Treatment of Covid-19 — Final Report | Drug treatment |
| BelenguerMuncharaz 2020 | Effectiveness of non-invasive ventilation in intensive care unit admitted patients due to SARS-CoV-2 pneumonia | Not a randomized trial |
| Bian 2020 | Meplazumab treats COVID-19 pneumonia: an open-labelled, concurrent controlled add-on clinical trial | Not a randomized trial |
| Biosciences 2020 | Efficacy and Safety Study of Allogeneic HB-adMSCs for the Treatment of COVID-19 | Randomized trial with no results (e.g. trial registry) |
| Bohua 2020 | Large-sample prospective clinical study of Huoxiangzhengqi Oral Liquid and Jinhaojiere Granules in the preventive intervention of COVID-19 in the community %J Chinese Journal of Chinese Materia Medica | Traditional Chinese medicine |
| Borba 2020 | Chloroquine diphosphate in two different dosages as adjunctive therapy of hospitalized patients with severe respiratory syndrome in the context of coronavirus (SARS-CoV-2) infection: Preliminary safety results of a randomized, double-blinded, phase IIb clinical trial (CloroCovid-19 Study) | Drug treatment |
| Borba 2020 | Effect of High vs Low Doses of Chloroquine Diphosphate as Adjunctive Therapy for Patients Hospitalized With Severe Acute Respiratory Syndrome Coronavirus 2 (SARS-CoV-2) Infection: A Randomized Clinical Trial | Drug treatment |
| Boyko 2020 | The first experience of using the drug Angiovit in the complex treatment of the acute stage of COVID-19 infection | Not a randomized trial |
| Branch-Elliman 2020 | Sarilumab for Patients With Moderate COVID-19 Disease: A Randomized Controlled Trial With a Play-The-Winner Design | Randomized trial with no results (e.g. trial registry) |
| Bretthauer 2020 | Randomized Re-Opening of Training Facilities during the COVID-19 pandemic | Others (e.g. mouthwash) |
| Brown 2020 | Hydroxychloroquine vs. Azithromycin for Hospitalized Patients with COVID-19 (HAHPS): Results of a Randomized, Active Comparator Trial | Drug treatment |
| Bundgaard 2020 | Effectiveness of Adding a Mask Recommendation to Other Public Health Measures to Prevent SARS-CoV-2 Infection in Danish Mask Wearers : A Randomized Controlled Trial. | Personal protective equipment |
| Cabanov 2020 | Treatment with tocilizumab does not inhibit induction of anti-COVID-19 antibodies in patients with severe SARS-CoV-2 infection | Not a randomized trial |
| Cadegiani 2020 | 5-Alpha-Reductase Inhibitors Reduce Remission Time of COVID-19: Results From a Randomized Double Blind Placebo Controlled Interventional Trial in 130 SARS- CoV-2 Positive Men | Drug treatment |
| Cadegiani 2021 | Dutasteride Reduces Viral Shedding, In ammatory Responses and Time-to-Remission in COVID-19: Biochemical Findings of a Randomized Double- Blind Placebo Controlled Interventional Trial (DUTA AndroCoV-Trial - Biochemical). | Drug treatment |
| Cadegiani 2021 | Dutasteride Reduces Viral Shedding, In ammatory Responses and Time-to-Remission in COVID-19: Biochemical Findings of a Randomized Double- Blind Placebo Controlled Interventional Trial (DUTA AndroCoV-Trial - Biochemical). | Drug treatment |
| Cai 2020 | Experimental Treatment with Favipiravir for COVID-19: An Open-Label Control Study | Not a randomized trial |
| CallejasRubio 2020 | Eficacia de los pulsos de corticoides en pacientes con s√≠ndrome de liberaci√≥n de citocinas inducido por infecci√≥n por SARS-CoV-2 | Not a randomized trial |
| Cantini 2020 | Baricitinib therapy in COVID-19: A pilot study on safety and clinical impact | Not a randomized trial |
| Cao 2020 | A Trial of Lopinavir-Ritonavir in Adults Hospitalized with Severe Covid-19 | Drug treatment |
| Cao 2020 | Ruxolitinib in treatment of severe coronavirus disease 2019 (COVID-19): A multicenter, single-blind, randomized controlled trial | Drug treatment |
| Carbillon 2021 | Hydroxychloroquine at usual doses as an option for coronavirus disease 2019 treatment. | Not a randomized trial |
| Carr 2020 | A new clinical trial to test high-dose vitamin C in patients with COVID-19 | Not a randomized trial |
| Casadevall 2020 | A Randomized Trial of Convalescent Plasma for COVID-19-Potentially Hopeful Signals | Not a randomized trial |
| Castillo 2020 | Effect of Calcifediol Treatment and best Available Therapy versus best Available Therapy on Intensive Care Unit Admission and Mortality Among Patients Hospitalized for COVID-19: A Pilot Randomized Clinical study¨90pt plus1fill | Drug treatment |
| Cavalcanti 2020 | Hydroxychloroquine with or without Azithromycin in Mild-to-Moderate Covid-19 | Drug treatment |
| Cavalcanti 2020 | Correction to: Hydroxychloroquine with or without Azithromycin in Mild-to-Moderate Covid-19 | Drug treatment |
| Cavalcanti 2020 | Correction to: Hydroxychloroquine with or without Azithromycin in Mild-to-Moderate Covid-19 | Drug treatment |
| Chaccour 2020 | The effect of early treatment with ivermectin on viral load, symptoms and humoral response in patients with mild COVID-19: a pilot, double-blind, placebo- controlled, randomized clinical trial | Drug treatment |
| Chachar 2021 | Effectiveness of Ivermectin in SARS-CoV-2/COVID-19 Patients | Drug treatment |
| Chang 2020 | Safety and Efficacy of Bronchoscopy in Critically Ill Patients with Coronavirus Disease 2019 | Not a randomized trial |
| Che 2020 | Randomized, double-blinded and placebo-controlled phase II trial of an inactivated SARS-CoV-2 vaccine in healthy adults | Vaccine |
| Chen 2020 | SARS-CoV-2 Neutralizing Antibody LY-CoV555 in Outpatients with Covid-19 | Blood product |
| Chen 2020 | A preliminary study of hydroxychloroquine sulfate in the treatment of patients with common 2019 coronavirus disease (COVID-19) | Drug treatment |
| Chen 2020 | [A pilot study of hydroxychloroquine in treatment of patients with moderate COVID-19] | Drug treatment |
| Chen 2020 | Efficacy of hydroxychloroquine in patients with COVID-19: results of a randomized clinical trial | Drug treatment |
| Chen 2020 | Favipiravir versus Arbidol for COVID-19: A Randomized Clinical Trial | Drug treatment |
| Chen 2020 | A pilot study of hydroxychloroquine in treatment of patients  with moderate COVID-19 | Drug treatment |
| Chen 2020 | Efficacy and safety of chloroquine or hydroxychloroquine in moderate type of COVID-19: a prospective open-label randomized controlled study | Drug treatment |
| Chen 2020 | A Multicenter, randomized, open-label, controlled trial to evaluate the efficacy and 4 tolerability of hydroxychloroquine and a retrospective study in adult patients with mild 5 to moderate Coronavirus disease 2019 (COVID-19) | Drug treatment |
| Chen 2020 | A multicenter, randomized, open-label, controlled trial to evaluate the efficacy and tolerability of hydroxychloroquine and a retrospective study in adult patients with mild to moderate coronavirus disease 2019 (COVID-19) | Drug treatment |
| Chen 2020 | Effect of Recombinant Human Granulocyte Colony–Stimulating Factor for Patients With Coronavirus Disease 2019 (COVID-19) and Lymphopenia A Randomized Clinical Trial | Drug treatment |
| Chen 2020 | Antiviral Activity and Safety of Darunavir/Cobicistat for Treatment of COVID-19 | Not a randomized trial |
| Chen 2020 | Clinical efficacy and safety of favipiravir in the treatment of COVID-19 patients | Not a randomized trial |
| Chen 2020 | The efficacy and safety of traditional Chinese medicines, modified Radix Fici Simplicissimae, combined with Western medicines amongst patients infected with the 2019 novel coronavirus (SARS-CoV-2) in tropical tourist area, China | Traditional Chinese medicine |
| ClinicalTrials.gov | Pilot Study on Cytokine Filtration in COVID-19 ARDS | Randomized trial with no results (e.g. trial registry) |
| ClinicalTrials.gov | Hyperbaric Oxygen Therapy Effect in COVID-19 RCT (HBOTCOVID19) | Randomized trial with no results (e.g. trial registry) |
| ClinicalTrials.gov | Pilot Randomized Controlled Trial: Fluoxetine to Reduce Hospitalization From COVID-19 Infection (FloR COVID-19) | Randomized trial with no results (e.g. trial registry) |
| ClinicalTrials.gov | Plasma Collection From Convalescent and/or Immunized Donors for the Treatment of COVID-19 | Randomized trial with no results (e.g. trial registry) |
| ClinicalTrials.gov | Ruxolitinib for Treatment of Covid-19 Induced Lung Injury ARDS | Randomized trial with no results (e.g. trial registry) |
| ClinicalTrials.gov | COVID-19 Patient Positioning Pragmatic Trial | Randomized trial with no results (e.g. trial registry) |
| Cohen 2021 | Continuation versus discontinuation of renin–angiotensin system inhibitors in patients admitted to hospital with COVID-19: a prospective, randomised, open-label trial | Drug treatment |
| Colombia 2020 | Effectiveness and Safety of Medical Treatment for SARS-CoV-2 (COVID-19) in Colombia | Randomized trial with no results (e.g. trial registry) |
| Cong 2020 | The clinical efficacy of high-dose or low-dose chloroquine diphosphate as an adjuvant treatment for inpatients with novel coronavirus pneumonia%J Chinese Medical Journal | Not a randomized trial |
| Corral-Gudino 2020 | GLUCOCOVID: A controlled trial of methylprednisolone in adults hospitalized with COVID-19 pneumonia | Drug treatment |
| Cremer 2021 | Canakinumab to Reduce Deterioration of Cardiac and Respiratory Function in Sars-cov2 Associated Myocardial Injury With Heightened Inflammation (canakinumab in Covid-19 Cardiac Injury): Results From the Randomized Three C Study | Drug treatment |
| Cruz 2020 | Treatment with an Anti-CK2 Synthetic Peptide Improves Clinical Response in 3 Covid-19 Patients with Pneumonia. A Randomized and Controlled Clinical Trial | Drug treatment |
| Cruz 2020 | Treatment with an Anti-CK2 Synthetic Peptide Improves Clinical Response in COVID-19 Patients with Pneumonia. A Randomized and Controlled Clinical Trial | Drug treatment |
| Dabbous 2020 | A Randomized Controlled Study Of Favipiravir Vs Hydroxychloroquine In COVID-19 Management: What Have We Learned So Far? | Drug treatment |
| Davoodi 2020 | Febuxostat therapy in outpatients with suspected COVID-19: A clinical trial | Drug treatment |
| Davoodi 2020 | Febuxostat as an alternative therapy in COVID-19: A double-blind randomized clinical trial | Drug treatment |
| Davoudi-Monfared 2020 | Efficacy and safety of interferon β-1a in treatment of severe COVID-19: A randomized clinical trial | Drug treatment |
| Davoudi-Monfared 2020 | Efficacy and safety of interferon β-1a in treatment 1 of severe COVID-19: A randomized 2 clinical trial | Drug treatment |
| Davoudi-Monfared 2020 | A Randomized Clinical Trial of the Efficacy and Safety of Interferon -1a in Treatment of Severe COVID-19 | Drug treatment |
| de Alencar 2020 | Double-blind, randomized, placebo-controlled trial with N-acetylcysteine for treatment of severe acute respiratory syndrome caused by COVID-19 | Drug treatment |
| De Marchi 2020 | Effects of photobiomodulation therapy combined with static magnetic field (PBMT-sMF) in patients with severe COVID-19 requiring intubation: a pragmatic randomized placebo-controlled trial | Exercise/rehabilitation |
| Deftereos 2020 | Effect of Colchicine vs Standard Care on Cardiac and Inflammatory Biomarkers and Clinical Outcomes in Patients Hospitalized With Coronavirus Disease 2019 The GRECCO-19 Randomized Clinical Trial | Drug treatment |
| Delgado-Enciso 2020 | Patient-Reported Health Outcomes After Treatment of COVID-19 with Nebulized and/or Intravenous Neutral Electrolyzed Saline Combined with Usual Medical Care Versus Usual Medical care alone: A Randomized, Open-Label, Controlled Trial. | Drug treatment |
| Dequin 2020 | Effect of Hydrocortisone on 21-Day Mortality or Respiratory Support Among Critically Ill Patients With COVID-19 A Randomized Clinical Trial | Drug treatment |
| Devos 2020 | Correction to: A randomized, multicentre, open-label phase II proof-of-concept trial investigating the clinical efficacy and safety of the addition of convalescent plasma to the standard of care in patients hospitalized with COVID-19: the Donated Antibodi | Randomized trial with no results (e.g. trial registry) |
| Dhibar 2020 | Post-exposure prophylaxis with hydroxychloroquine for the prevention of COVID-19, a myth or a reality? The PEP-CQ Study. | Not a randomized trial |
| Dillon 2020 | A real-world data study of coronavirus-2019 disease severity in patients with multiple sclerosis treated with ocrelizumab | Not a randomized trial |
| Ding 2020 | Study on the Clinical Efficacy and Mechanism of Qingfei Toxie Fuzheng Decoction in the Treatment of Novel Coronavirus Pneumonia | Traditional Chinese medicine |
| Ding 2020 | Clinical efficacy and mechanism of Qingfeitouxiefuzheng Fang in the treatment of new coronavirus pneumonia | Traditional Chinese medicine |
| Diurno 2020 | Eculizumab treatment in patients with COVID-19: preliminary results from real life ASL Napoli 2 Nord experience | Not a randomized trial |
| Doi 2020 | A prospective, randomized, open-label trial of early versus late favipiravir in 2 hospitalized patients with COVID-19 | Drug treatment |
| Doi 2020 | A prospective, randomized, open-label trial of early versus late favipiravir in 2 hospitalized patients with COVID-19 | Drug treatment |
| Duan 2020 | Clinical Observation of Jinhua Qinggan Granule in Treating Pneumonia Infected by Novel Coronavirus | Traditional Chinese medicine |
| Duan 2020 | Clinical Observation of Jinhua Qinggan Granules in Treating Pneumonia Infected by Novel Coronavirus | Traditional Chinese medicine |
| Duarte 2020 | Telmisartan for treatment of Covid-19 patients: an open randomized clinical trial - A preliminary report. | Drug treatment |
| Dubee 2020 | A placebo-controlled double blind trial of hydroxychloroquine in mild-to-moderate COVID-19 | Drug treatment |
| Duong-Quy 2020 | The use of exhaled nitric oxide and peak expiratory flow to demonstrate improved breathability and antimicrobial properties of novel face mask made with sustainable filter paper and Folium Plectranthii amboinicii oil: additional option for mask shortage d | Not exposed to or diagnosed with (confirmed or suspected) COVID-19 |
| Edalatifard 2020 | Intravenous methylprednisolone pulse as a treatment for hospitalised severe COVID-19 patients: results from a randomised controlled clinical trial | Drug treatment |
| Erdem 2020 | Treatment of SARS-cov-2 pneumonia with favipiravir: Early results from the Ege University cohort, Turkey. | Not a randomized trial |
| Eslami 2020 | The impact of sofosbuvir/daclatasvir or ribavirin in patients with severe COVID-19 | Not a randomized trial |
| Farahani 2020 | Evaluation of the Efficacy of Methylprednisolone Pulse Therapy in Treatment of Covid-19 Adult Patients with Severe Respiratory Failure: Randomized, Clinical Trial | Drug treatment |
| Feeney 2020 | The COVIRL-001 Trial: A multicentre, prospective, randomised trial comparing standard of care (SOC) alone, SOC plus hydroxychloroquine monotherapy or SOC plus a combination of hydroxychloroquine and azithromycin in the treatment of non- critical, SARS-CoV | Randomized trial with no results (e.g. trial registry) |
| Feily 2020 | COVID-19: Pentoxifylline as a potential adjuvant treatment | Not a randomized trial |
| Feld 2020 | Peginterferon-lambda for the treatment of COVID-19 in outpatients | Drug treatment |
| Feng 2020 | Differentiation between COVID-19 and bacterial pneumonia using radiomics of chest computed tomography and clinical features | Not a randomized trial |
| Flisiak 2020 | Remdesivir-based therapy improved recovery of patients with COVID-19 in the SARSTer multicentre, real-world study. | Not a randomized trial |
| Folegatti 2020 | Safety and immunogenicity of the ChAdOx1 nCoV-19 vaccine against SARS-CoV-2: a preliminary report of a phase 1/2, single-blind, randomised controlled trial | Vaccine |
| Folegatti 2020 | Erratum: Department of Error: Safety and immunogenicity of the ChAdOx1 nCoV-19 vaccine against SARS-CoV-2: a preliminary report of a phase 1/2, single-blind, randomised controlled trial. | Vaccine |
| Folegatti 2020 | Safety and immunogenicity of the ChAdOx1 nCoV-19 vaccine against SARS-CoV-2: a preliminary report of a phase 1/2, single-blind, randomised controlled trial (vol 396, pg 467, 2020) | Vaccine |
| Folegatti 2020 | Safety and immunogenicity of the ChAdOx1 nCoV-19 vaccine against SARS-CoV-2: a preliminary report of a phase 1/2, single-blind, randomised controlled trial. | Vaccine |
| Fong 2020 | Impact of aerosol box on intubation during COVID-19: a simulation study of normal and difficult airways | Personal protective equipment |
| Fragoso-Saavedra 2020 | A parallel-group, multicenter randomized, double-blinded, placebo-controlled, phase 2/3, clinical trial to test the efficacy of pyridostigmine bromide at low doses to reduce mortality or invasive mechanical ventilation in adults with severe SARS-CoV-2 inf | Randomized trial with no results (e.g. trial registry) |
| Frandell 2020 | The Effects of Electronic Alert Letters in the Time of COVID-19 (preprint) | Not exposed to or diagnosed with (confirmed or suspected) COVID-19 |
| Freedenberg 2021 | Neutralizing Activity to SARS-CoV-2 of Convalescent and Control Plasma Used in a Randomized Controlled Trial | Blood product |
| Fu 2020 | An open-label, randomized trial of the combination of IFN-k plus TFF2 with standard care in the treatment of patients with moderate COVID-19 | Drug treatment |
| Fu 2020 | An open-label, randomized trial of the combination of IFN-k plus TFF2 with standard care in the treatment of patients with moderate COVID-19 | Drug treatment |
| Fu 2020 | Clinical Study on 37 Cases of Novel Coronavirus Pneumonia Treated by Integrated Traditional Chinese and Western Medicine | Traditional Chinese medicine |
| Furtado 2020 | Azithromycin in addition to standard of care versus standard of care alone in the treatment of patients admitted to the hospital with severe COVID-19 in Brazil (COALITION II): a randomised clinical trial | Drug treatment |
| Garcia-Fernandez 2020 | Usefulness and safety of self-electrocardiographic monitoring during treatment with hydroxychloroquine and azithromycin in COVID-19 patients | Not a randomized trial |
| Gautret 2020 | Hydroxychloroquine and azithromycin as a treatment of COVID-19: results of an open-label non-randomized clinical trial | Not a randomized trial |
| Gautret 2020 | Hydroxychloroquine and Azithromycin as a treatment of COVID-19: preliminary results of an open-label non-randomized clinical trial | Not a randomized trial |
| Gautret 2021 | Safety profile of hydroxychloroquine and azithromycin combined treatment in COVID-19 patients | Not a randomized trial |
| Gautret 2021 | Clinical efficacy and safety profile of hydroxychloroquine and azithromycin against COVID-19 | Not a randomized trial |
| Gergi 2020 | Thrombo-inflammation response to Tocilizumab in COVID-19 | Not a randomized trial |
| Ghaderkhani 2020 | Efficacy and Safety of Arbidol in Treatment of Patients with COVID-19 Infection: A Randomized Clinical Trial | Drug treatment |
| Ghandehari 2021 | Progesterone in addition to standard of care versus standard of care alone in the treatment of men admitted to the hospital with moderate to severe COVID-19: a randomised control phase 1 trial | Drug treatment |
| Gharbharan 2020 | Convalescent Plasma for COVID-19. A randomized clinical trial | Blood product |
| Gharbharan 2020 | Convalescent Plasma for COVID-19. A randomized clinical trial | Blood product |
| Gharebaghi 2020 | Intravenous Immunoglobulin Gamma for Severe Cases of Coronavirus Disease of 2019: A Randomized Placebo-Controlled Double-Blinded Clinical Trial | Blood product |
| Gharebaghi 2020 | The use of intravenous immunoglobulin gamma for the treatment of severe coronavirus disease 2019: A randomised placebo-controlled double-blind clinical trial | Blood product |
| Gharebaghi 2020 | The use of intravenous immunoglobulin gamma for the treatment of severe coronavirus disease 2019: A randomised placebo-controlled double-blind clinical trial | Blood product |
| Gharebaghi 2020 | Correction to: The use of intravenous immunoglobulin gamma for the treatment of severe coronavirus disease 2019: A randomised placebo-controlled double-blind clinical trial | Blood product |
| Gharebaghi 2020 | Correction to: The use of intravenous immunoglobulin gamma for the treatment of severe coronavirus disease 2019: A randomised placebo-controlled double-blind clinical trial | Blood product |
| Gokhale 2020 | Tocilizumab improves survival in patients with persistent hypoxia in severe COVID-19 pneumonia | Not a randomized trial |
| Gold 2020 | Efficacy of m-Health for the detection of adverse events following immunization - The stimulated telephone assisted rapid safety surveillance (STARSS) randomised control trial | Others (e.g. mouthwash) |
| Goldman 2020 | Remdesivir for 5 or 10 Days in Patients with Severe Covid-19 | Drug treatment |
| Gonzalez-Ochoa 2020 | Sulodexide in the treatment of patients with early stages of COVID-19: a randomised controlled trial | Drug treatment |
| Gordon 2021 | Interleukin-6 Receptor Antagonists in Critically Ill Patients with Covid-19 - Preliminary report | Drug treatment |
| Govind 2020 | Clozapine Treatment and Risk of COVID-19 | Not a randomized trial |
| Guan 2020 | Erratum to hydrogen/oxygen mixed gas inhalation improves disease severity and dyspnea in patients with Coronavirus disease 2019 in a recent multicenter, open-label clinical trial | Not a randomized trial |
| Guan 2020 | Hydrogen/oxygen mixed gas inhalation improves disease severity and dyspnea in patients with Coronavirus disease 2019 in a recent multicenter, open-label clinical trial | Not a randomized trial |
| Guvenmez 2020 | The comparison of the effectiveness of lincocin® and azitro® in the treatment of covid-19-associated pneumonia: A prospective study | Drug treatment |
| HacibeklroGlu 2020 | Efficacy of convalescent plasma according to blood groups in COVID-19 patients | Not a randomized trial |
| Han 2020 | RUXCOVID: A phase 3, randomized, placebo-controlled study evaluating the efficacy and safety of ruxolitinib in patients with COVID-19-associated cytokine storm | Randomized trial with no results (e.g. trial registry) |
| Hashim 2020 | Controlled randomized clinical trial on using Ivermectin with Doxycycline for treating COVID-19 patients in Baghdad, Iraq | Not a randomized trial |
| Hassan 2020 | Olfactory disturbances as presenting manifestation among Egyptian patients with COVID-19: Possible role of zinc | Drug treatment |
| Havlichek 2020 | A Trial of Lopinavir-Ritonavir in Covid-19 | Not a randomized trial |
| Hermine 2020 | Effect of Tocilizumab vs Usual Care in Adults Hospitalized With COVID-19 and Moderate or Severe Pneumonia A Randomized Clinical Trial | Drug treatment |
| Holubovska 2021 | Enisamium is an inhibitor of the SARS-CoV-2 RNA polymerase and shows 2 improvement of recovery in COVID-19 patients in an interim analysis of a 3 clinical trial | Drug treatment |
| Hong 2020 | Early Hydroxychloroquine Administration for Rapid Severe Acute Respiratory Syndrome Coronavirus 2 Eradication | Not a randomized trial |
| Hong 2020 | Celebrex adjuvant therapy on COVID-19: An experimental study | Not a randomized trial |
| Hong 2020 | Celebrex Adjuvant Therapy on Coronavirus Disease 2019: An Experimental Study | Not a randomized trial |
| Horby 2020 | Lopinavir–ritonavir in patients admitted to hospital with COVID-19 (RECOVERY): a randomised, controlled, open-label, platform trial | Drug treatment |
| Horby 2020 | Lopinavir-ritonavir in Hospitalised Patients with 4 COVID-19 – Preliminary report from a randomised, 5 controlled, open-label, platform trial | Drug treatment |
| Horby 2020 | Effect of Hydroxychloroquine in Hospitalized Patients with Covid-19 | Drug treatment |
| Horby 2020 | Azithromycin in Hospitalised Patients with COVID-19 (RECOVERY): a randomised, controlled, open-label, platform trial | Drug treatment |
| Horby 2020 | Effect of Dexamethasone in Hospitalized Patients with COVID-19 – Preliminary Report | Drug treatment |
| Horby 2020 | Dexamethasone in Hospitalized Patients with Covid-19 — Preliminary Report | Drug treatment |
| Horby 2020 | Effect of Hydroxychloroquine in Hospitalized Patients with COVID-19: Preliminary results from a 5 multi-centre, randomized, controlled trial. | Drug treatment |
| Hu 2020 | A Small-Scale Medication of Leflunomide as a Treatment of COVID-19 in an Open-Label Blank-Controlled Clinical Trial | Drug treatment |
| Hu 2020 | Multi-center clinical observation of honeysuckle oral liquid combined with western medicine in the treatment of common type of COVID-19 | Traditional Chinese medicine |
| Hu 2020 | Chansu Injection Improves the Respiratory Function of Severe COVID-19 Patients | Traditional Chinese medicine |
| Hu 2020 | Efficacy and Safety of Lianhuaqingwen Capsules, a repurposed Chinese Herb, in Patients with Coronavirus disease 2019: A multicenter, prospective, randomized controlled trial | Traditional Chinese medicine |
| Hu 2020 | Multi-center clinical observation of honeysuckle oral liquid combined with western medicine in the treatment of common type of new coronavirus pneumonia | Traditional Chinese medicine |
| Hua 2020 | Short-term skin reactions following use of N95 respirators and medical masks | Personal protective equipment |
| Huang 2020 | Treating COVID-19 with Chloroquine | Drug treatment |
| Huang 2020 | Comparative effectiveness and safety of ribavirin plus interferon-alpha, lopinavir/ritonavir plus interferon-alpha and ribavirin plus lopinavir/ritonavir plus interferon-alpha in patients with mild to moderate novel coronavirus pneumonia: results of a randomized, open-labeled prospective study | Drug treatment |
| Huang 2020 | No Statistically Apparent Difference in Antiviral Effectiveness Observed Among Ribavirin Plus Interferon-Alpha, Lopinavir/Ritonavir Plus Interferon-Alpha, and Ribavirin Plus Lopinavir/Ritonavir Plus Interferon-Alpha in Patients With Mild to Moderate Coronavirus Disease 2019: Results of a Randomized, Open-Labeled Prospective Study | Drug treatment |
| Huimin 2020 | Clinical observation of Lianhua Qingke Granules in the treatment of mild and common new coronavirus pneumonia | Traditional Chinese medicine |
| Hung 2020 | Triple combination of interferon beta-1b, lopinavir–ritonavir, and ribavirin in the treatment of patients admitted to hospital with COVID-19: an open-label, randomised, phase 2 trial | Drug treatment |
| Husain 2020 | Role of doxycycline, oral steroids, and nasal steroid in treatment of anosmia due to COVID-19 with the new insights into the doxycycline activity. | Drug treatment |
| Ibrahim 2020 | Factors Associated with Good Patient Outcomes Following Convalescent Plasma in COVID-19: A Prospective Phase 2 Clinical Trial (preprint) | Removed from preprint server |
| Idelsis 2020 | Effect of combination of interferon alpha-2b and interferon-gamma or interferon alpha- 2b alone for elimination of SARS-CoV-2 viral RNA. Preliminary results of a randomized controlled clinical trial. | Drug treatment |
| Ikonomidis 2020 | Tocilizumab improves oxidative stress and endothelial glycocalyx: a mechanism that may explain the effects of biological treatment on COVID-19. (Special Issue: COVID-19 and treatments: particular emphasis on potential toxic effects.) | Not exposed to or diagnosed with (confirmed or suspected) COVID-19 |
| Ivashchenko 2020 | Interim Results of a Phase II/III Multicenter Randomized Clinical Trial of AVIFAVIR in Hospitalized Patients with COVID-19 | Drug treatment |
| Ivashchenko 2020 | AVIFAVIR for Treatment of Patients with Moderate COVID-19: Interim Results of a Phase II/III Multicenter Randomized Clinical Trial | Drug treatment |
| Ivashchenko 2020 | AVIFAVIR for Treatment of Patients with Moderate COVID-19: Interim Results of a Phase II/III Multicenter Randomized Clinical Trial | Drug treatment |
| Jagannathan | Peginterferon Lambda-1a for treatment of outpatients with uncomplicated COVID-19: a 3 randomized placebo-controlled trial | Drug treatment |
| Jagannathan 2020 | Peginterferon Lambda-1a for treatment of outpatients with uncomplicated COVID-19: a 3 randomized placebo-controlled trial | Drug treatment |
| Janapala 2020 | Novaferon, treatment in COVID-19 patients. | Not a randomized trial |
| Jeronimo 2020 | Methylprednisolone as Adjunctive Therapy for Patients Hospitalized With COVID-19 (Metcovid): A Randomised, Double-Blind, Phase IIb, Placebo-Controlled Trial | Drug treatment |
| Jungreis | Mathematical analysis of Córdoba calcifediol trial suggests strong role for Vitamin D in reducing ICU admissions of hospitalized COVID-19 patients | Drug treatment |
| Kalil 2020 | Baricitinib plus Remdesivir for Hospitalized Adults with Covid-19 | Drug treatment |
| Kamran 2020 | Clearing the fog: Is HCQ effective in reducing COVID-19 progression: A randomized controlled trial (preprint) | Not a randomized trial |
| Karaahmet 2020 | Potential effect of natural and anabolizan steroids in elderly patient with COVID-19 | Not a randomized trial |
| Karan 2020 | Clinical outcomes in patients with COVID-19 infection during phase IV studies of cladribine tablets for treatment of multiple sclerosis | Not a randomized trial |
| Keech 2020 | Phase 1-2 Trial of a SARS-CoV-2 Recombinant Spike Protein Nanoparticle Vaccine | Vaccine |
| Keech 2020 | First-in-Human Trial of a SARS CoV 2 Recombinant Spike Protein Nanoparticle Vaccine (preprint) | Vaccine |
| Khamis 2020 | Randomized Controlled Open Label Trial on the Use of Favipiravir Combined with Inhaled Interferon beta-1b in Hospitalized Patients with Moderate to Severe COVID-19 Pneumonia | Drug treatment |
| Khamis 2020 | Randomized controlled open label trial on the use of favipiravir combined with inhaled interferon beta-1b in hospitalized patients with moderate to severe COVID-19 pneumonia | Drug treatment |
| Khan 2020 | Ivermectin treatment may improve the prognosis of patients with COVID-19 | Not a randomized trial |
| Kimura 2020 | Interim Analysis of an Open-label Randomized Controlled Trial Evaluating Nasal Irrigations in Non-hospitalized Patients with COVID-19 | Drug treatment |
| Kimura 2020 | Interim analysis of an open-label randomized controlled trial evaluating nasal irrigations in non-hospitalized patients with coronavirus disease | Drug treatment |
| Kintscher 2020 | Plasma Angiotensin Peptide Profiling and ACE (Angiotensin-Converting Enzyme)-2 Activity in COVID-19 Patients Treated With Pharmacological Blockers of the Renin-Angiotensin System | Not a randomized trial |
| Kollias 2020 | Anticoagulation therapy in COVID-19: Is there a dose-dependent benefit? | Not a randomized trial |
| Krolewiecki 2020 | Antiviral effect of high-dose ivermectin in adults with COVID-19: a pilot randomised, controlled, open label, multicentre trial. | Drug treatment |
| Kumar 2020 | A two-arm, randomized, controlled, multi-centric, open-label Phase-2 study to evaluate the efficacy and safety of Itolizumab in moderate to severe ARDS patients due to COVID-19 | Drug treatment |
| Kumari 2020 | The Role of Vitamin C as Adjuvant Therapy in COVID-19 | Drug treatment |
| Labib Salem 2020 | The possible beneficial adjuvant effect of Influenza vaccine to minimize the severity of COVID-19 | Not a randomized trial |
| Lanthier 2020 | [In patients hospitalized for COVID-19, does dexamethasone reduce 28-days mortality compared to standard treatment?] | Not a randomized trial |
| Lanzoni 2020 | Umbilical Cord Mesenchymal Stem Cells for COVID-19 ARDS: A Double Blind, Phase 1/2a, Randomized Controlled Trial | Blood product |
| Lanzoni 2021 | Umbilical cord mesenchymal stem cells for COVID-19 acute respiratory distress syndrome: A double-blind, phase 1/2a, randomized controlled trial | Blood product |
| Le Bon 2021 | Efficacy and safety of oral corticosteroids and olfactory training in the management of COVID-19-related loss of smell. | Not a randomized trial |
| Lemos 2020 | Therapeutic versus prophylactic anticoagulation for severe COVID-19: A randomized phase II clinical trial (HESACOVID) | Drug treatment |
| Lenkens 2020 | [Medication and comedication in COVID-19 patients] | Not a randomized trial |
| Lenze 2020 | Fluvoxamine vs Placebo and Clinical Deterioration in Outpatients With Symptomatic COVID-19 A Randomized Clinical Trial | Drug treatment |
| Li 2020 | Effect of Convalescent Plasma Therapy on Time to Clinical Improvement in Patients With Severe and Life-threatening COVID-19 A Randomized Clinical Trial | Blood product |
| Li 2020 | Correction to: Effect of Convalescent Plasma Therapy on Time to Clinical Improvement in Patients With Severe and Life-threatening COVID-19 A Randomized Clinical Trial | Blood product |
| Li 2020 | Duplicate correction to: Effect of Convalescent Plasma Therapy on Time to Clinical Improvement in Patients With Severe and Life-threatening COVID-19 A Randomized Clinical Trial | Blood product |
| Li 2020 | Application of CareDose 4D combined with Karl 3D technology in the low dose computed tomography for the follow-up of COVID-19 | Diagnostic imaging |
| Li 2020 | Evaluation of low-dose CT protocol of novel coronavirus pneumonia based on infection prevention and control | Diagnostic imaging |
| Li 2020 | Engineered interferon alpha effectively improves clinical outcomes of COVID-19 patients | Drug treatment |
| Li 2020 | Efficacy and safety of lopinavir/ritonavir or arbidol in adult patients with mild/moderate COVID-19: an exploratory randomized controlled trial | Drug treatment |
| Li 2020 | An exploratory randomized, controlled study on the efficacy and safety of lopinavir/ritonavir or arbidol treating adult patients hospitalized with mild/moderate COVID-19 (ELACOI) | Drug treatment |
| Li 2020 | Recombinant super-compound interferon (rSIFN-co) versus interferon alfa in the treatment of moderate-to-severe COVID-19: a multicentre, randomised, phase 2 trial | Drug treatment |
| Li 2020 | Bromhexine Hydrochloride Tablets for the Treatment of Moderate COVID-19: An Open-label Randomized Controlled Pilot Study | Drug treatment |
| Li 2020 | Regarding “Ruxolitinib in treatment of severe coronavirus disease 2019 (COVID-19): A multicenter, single-blind, randomized controlled trial” | Not a randomized trial |
| Li 2020 | Observation on the application effect of high-flow humidified oxygen therapy through the nose in the treatment of new coronavirus pneumonia complicated with acute respiratory failure | Oxygen delivery |
| Li 2020 | COVID-19: Repeatedly Video-Watching vs Combined Video-Watching and Live Demonstration as Training to Healthcare Providers for Donning and Doffing Personal Protective Equipment | Psychological and educational |
| Li 2020 | Convalescent Plasma Treatment in Severe and Life-Threatening COVID-19: A Randomized Controlled Clinical Trial | Randomized trial with no results (e.g. trial registry) |
| Libster 2020 | Prevention of severe COVID-19 in the elderly by early high- titer plasma | Blood product |
| Libster 2021 | Early High-Titer Plasma Therapy to Prevent Severe Covid-19 in Older Adults | Blood product |
| Liesenborghs 2020 | ITRACONAZOLE FOR COVID-19: PRECLINICAL STUDIES AND A PROOF-OF-CONCEPT PILOT CLINICAL STUDY | Drug treatment |
| Liu 2020 | Effects of progressive muscle relaxation on anxiety and sleep quality in patients with COVID-19 | Exercise/rehabilitation |
| Liu 2020 | Significance and operation mode of moxibustion intervention for the group under quarantine after close contact with COVID-19 | Exercise/rehabilitation |
| Liu 2020 | Respiratory rehabilitation in elderly patients with COVID-19: A randomized controlled study | Not a randomized trial |
| Liu 2020 | The dual role of anti-viral therapy in the treatment of Coronavirus disease 2019. | Not a randomized trial |
| Liu 2020 | Efficacy of chloroquine versus lopinavir/ritonavir in mild/general COVID-19 infection: a prospective, open-label, multicenter, randomized controlled clinical study | Randomized trial with no results (e.g. trial registry) |
| Liu 2020 | Clinical effect of dulaglutide injection in the treatment of novel coronavirus pneumonia with type 2 diabetes | Randomized trial with no results (e.g. trial registry) |
| Liu 2020 | Research on the Significance and Operation Mode of Moxibustion Intervention for New Coronavirus Pneumonia Close Contact and Isolated Persons | Traditional Chinese medicine |
| Liu 2020 | Effect of moxibustion on clinical symptoms, peripheral blood inflammatory indexes and T lymphocyte subsets in patients with new coronavirus pneumonia | Traditional Chinese medicine |
| Liu 2020 | [Effect of moxibustion on clinical symptoms, peripheral inflammatory indexes and T lymphocyte subsets in COVID-19 patients] | Traditional Chinese medicine |
| Lopes 2020 | Beneficial effects of colchicine for moderate to severe COVID-19: an interim analysis of a randomized, double-blinded, placebo controlled clinical trial | Drug treatment |
| López-Zúñiga 2020 | High-Dose Corticosteroid Pulse Therapy¬†Increases the Survival Rate in COVID-19 Patients at Risk of Cytokine Storm (preprint) | Not a randomized trial |
| Lou 2020 | Clinical Outcomes and Plasma Concentrations of Baloxavir Marboxil and Favipiravir in COVID-19 Patients: An Exploratory Randomized, Controlled Trial | Drug treatment |
| Lou 2020 | Clinical Outcomes and Plasma Concentrations of Baloxavir Marboxil and Favipiravir in COVID-19 Patients: An Exploratory Randomized, Controlled Trial | Drug treatment |
| Lou 2020 | Clinical Outcomes and Plasma Concentrations of Baloxavir Marboxil and Favipiravir in COVID-19 Patients: An Exploratory Randomized, Controlled Trial | Drug treatment |
| Luis Callejas Rubio 2020 | Of Corticoid Pulses In Patients With Cytokine Release Syndrome Induced By Sars-Cov-2 Infection | Not a randomized trial |
| Lundgren 2020 | A Neutralizing Monoclonal Antibody for Hospitalized Patients with Covid-19 | Blood product |
| Luo 2020 | Application of chest low-dose CT screening of Corona Virus Disease 2019 with a third-generation dual-source scanner | Diagnostic imaging |
| Lyngbakken 2020 | A pragmatic randomized controlled trial reports the efficacy of hydroxychloroquine on coronavirus disease 2019 viral kinetics | Drug treatment |
| Lyngbakken 2020 | A pragmatic randomized controlled trial reports lack of efficacy of hydroxychloroquine on coronavirus disease 2019 viral kinetics | Drug treatment |
| Madabhushi 2020 | Time to adapt in the pandemic era: a prospective randomized non -inferiority study comparing time to intubate with and without the barrier box | Personal protective equipment |
| Mahevas 2020 | No evidence of clinical efficacy of hydroxychloroquine in patients hospitalized for COVID-19 infection with oxygen requirement: results of a study using routinely collected data to emulate a target trial | Not a randomized trial |
| Mahmud 2020 | Clinical Trial of Ivermectin Plus Doxycycline for the Treatment of Confirmed Covid-19 Infection | Drug treatment |
| Maldonado 2020 | Pentoxifylline decreases serum LDH levels and increases lymphocyte count in COVID-19 patients: Results from an external pilot study | Drug treatment |
| Mansour 2020 | Pharmacological inhibition of the kinin-kallikrein system in severe COVID-19 – A proof-of-concept study | Drug treatment |
| Marshall 2020 | A phase II trial to promote recovery from COVID-19 with endocrine therapy | Randomized trial with no results (meeting abstract) |
| Masiá 2020 | Lack of detrimental effect of corticosteroids on antibody responses to SARS-CoV-2 and viral clearance in patients hospitalized with COVID-19 | Not a randomized trial |
| Mehboob 2020 | Aprepitant as a combinant with Dexamethasone reduces the inflammation via Neurokinin 1 Receptor Antagonism in severe to critical Covid-19 patients and potentiates respiratory recovery: A novel therapeutic approach | Drug treatment |
| Meng 2020 | An experimental trial of recombinant human interferon alpha nasal drops to prevent coronavirus disease 2019 in medical staff in an epidemic area | Not a randomized trial |
| Miller 2020 | Auxora versus standard of care for the treatment of severe or critical COVID-19 pneumonia: results from a randomized controlled trial | Drug treatment |
| MingZhong 2020 | A Randomized, Single-blind, Group sequential, Active-controlled Study to evaluate the clinical efficacy and safety of α-Lipoic acid for critically ill patients with coronavirus disease 2019（COVID-19） | Drug treatment |
| Mitja 2020 | Hydroxychloroquine alone or in combination with Cobicistat-boosted Darunavir for treatment of mild Covid-19: a cluster-randomized clinical trial | Drug treatment |
| Mitjà 2020 | Hydroxychloroquine for Early Treatment of Adults with Mild Covid-19: A Randomized-Controlled Trial | Drug treatment |
| Mohamed 2020 | EARLY VIRAL CLEARANCE AMONG COVID-19 PATIENTS WHEN GARGLING WITH POVIDONE-IODINE AND ESSENTIAL OILS: A PILOT CLINICAL TRIAL (preprint) | Others (e.g. mouthwash) |
| Mohammadzadeh 2020 | Effectiveness of electronic health care and drug monitoring program to prevent COVID-19 and adherence to therapeutic regimen in patients with ischemic heart disease - A pilot study | Others (e.g. mouthwash) |
| Moisset 2021 | Nasopharyngeal swab-induced pain for SARS-CoV-2 screening: a randomised controlled trial of conventional and self-swabbing. | Not exposed to or diagnosed with (confirmed or suspected) COVID-19 |
| Moniuszko-Malinowska 2021 | Convalescent Plasma Transfusion for the Treatment of COVID-19‚ÄîExperience from Poland: A Multicenter Study | Not a randomized trial |
| Monk 2020 | Safety and efficacy of inhaled nebulised interferon beta-1a (SNG001) for treatment of SARS-CoV-2 infection: a randomised, double-blind, placebo-controlled, phase 2 trial | Drug treatment |
| Mugheddu 2020 | Safety of secukinumab treatment in COVID-19 affected psoriatic patients. | Not a randomized trial |
| Mulligan 2020 | Phase 1/2 study of COVID-19 RNA vaccine BNT162b1 in adults | Vaccine |
| Murai 2020 | Effect of Vitamin D3 Supplementation vs Placebo on Hospital Length of Stay in 2 Patients with Severe COVID-19: A Multicenter, Double-blind, Randomized 3 Controlled Trial | Drug treatment |
| Myasnikov 2020 | Efficacy of Interferon Gamma in the Prevention of SARS-CoV-2 Infection (COVID-19): Results of a Prospective Controlled Trial | Not a randomized trial |
| Namazi 2020 | Ellagic acid can possibly be an adjuvant treatment for COVID-19 | Not a randomized trial |
| Niaee 2020 | Ivermectin as an adjunct treatment for hospitalized adult COVID-19 patients: A randomized multi-center clinical trial | Drug treatment |
| Nojomi 2020 | Effect of Arbidol on COVID-19: A Randomized Controlled Trial | Drug treatment |
| Nojomi 2020 | Effect of Arbidol (Umifenovir) on COVID-19: a randomized controlled trial | Drug treatment |
| NoorAzhar 2020 | COVID-19 aerosol box as protection from droplet and aerosol contaminations in healthcare workers performing airway intubation: a randomised cross-over simulation study | Personal protective equipment |
| NR 2020 | Ruijin Hospital Affiliated to Shanghai Jiaotong University School of Medicine released the results of a multi-center clinical study of hydroxychloroquine in the treatment of novel coronavirus pneumonia %J Journal of Shanghai Jiaotong University (Medical Edition) | Not a randomized trial |
| Omrani 2020 | Randomized double-blinded placebo-controlled trial of hydroxychloroquine with or without azithromycin for virologic cure of non-severe Covid-19 | Drug treatment |
| Padmanabhan 2020 | Phase II Clinical trial for Evaluation of BCG as potential therapy for COVID-19 | Drug treatment |
| Pan 2020 | Repurposed antiviral drugs for COVID-19 –interim WHO SOLIDARITY trial results | Drug treatment |
| Pan 2020 | Repurposed Antiviral Drugs for Covid-19 — Interim WHO Solidarity Trial Results | Drug treatment |
| Pang 2020 | Efficacy and tolerability of bevacizumab in patients with severe Covid -19 | Not a randomized trial |
| Panigada 2020 | Effect of heparin on viscoelastic parameters of COVID-19 critically ill patients. A Viscoelastic Coagulation Monitor (VCM) analysis | Not a randomized trial |
| Papamanoli 2020 | 76. Effect of Early Administration of Systemic Corticosteroids on Outcomes in Patients with COVID-19 Pneumonia | Not a randomized trial |
| Petrak 2020 | Early Tocilizumab Dosing is Associated with Improved Survival In Critically Ill Patients Infected With Sars-CoV-2 | Not a randomized trial |
| Piralla 2020 | Residual SARS-CoV-2 RNA in nasal swabs of convalescent COVID-19 patients: Is prolonged quarantine always justified? | Not a randomized trial |
| Plaze 2020 | [Repurposing chlorpromazine to treat COVID-19: the reCoVery study] | Randomized trial with no results (e.g. trial registry) |
| Plaze 2020 | Repurposing of chlorpromazine in COVID-19 treatment: the reCoVery study | Randomized trial with no results (e.g. trial registry) |
| Polack 2020 | Safety and Efficacy of the BNT162b2 mRNA Covid-19 Vaccine | Vaccine |
| Pompeii 2020 | Training and Fit Testing of Health Care Personnel for Reusable Elastomeric Half-Mask Respirators Compared With Disposable N95 Respirators | Personal protective equipment |
| Potere 2020 | Low-dose subcutaneous tocilizumab to prevent disease progression in patients with moderate COVID-19 pneumonia and hyperinflammation. (Special Issue: Coronavirus (COVID-19) collection.) | Not a randomized trial |
| Poulakou 2021 | Beneficial Effects of Intermediate Dosage of Anticoagulation Treatment on the Prognosis of Hospitalized COVID-19 Patients: The ETHRA Study. | Not a randomized trial |
| Pu 2020 | An in-depth investigation of the safety and immunogenicity of an inactivated SARS-CoV-2 vaccine (preprint) | Vaccine |
| Qamar 2020 | Protective Effects of CVD and DM Medications in SARS-CoV-2 Infection | Not a randomized trial |
| Qiu 2020 | Observation on the Efficacy of Maxing Xuanfei Jiedu Decoction in the Treatment of Common New Type Coronavirus Pneumonia | Traditional Chinese medicine |
| RachesElla 2020 | Safety and immunogenicity clinical trial of an inactivated SARS-CoV-2 vaccine, BBV152 (a phase 2, double-blind, randomised controlled trial) and the persistence of immune responses from a phase 1 follow-up report | Vaccine |
| Rahmani 2020 | Interferon β-1b in treatment of severe COVID-19: a randomized clinical trial | Drug treatment |
| Rahmani 2020 | Interferon beta-1b in treatment of severe COVID-19: A randomized clinical trial | Drug treatment |
| Raj 2020 | Effect of a novel ayurvedic preparation, Raj Nirvan Bati (rnb), on symptomatic patients of covid-19 | Traditional Chinese medicine |
| Ramasamy 2020 | Safety and immunogenicity of ChAdOx1 nCoV-19 vaccine administered in a prime-boost regimen in young and old adults (COV002): a single-blind, randomised, controlled, phase 2/3 trial | Vaccine |
| Rao 2020 | AYUSH medicine as add-on therapy for mild category COVID-19; an open label randomised, controlled clinical trial. | Traditional Chinese medicine |
| Rasmussen 2020 | Pulmonary administration of remdesivir in the treatment of COVID-19 | Not a randomized trial |
| Rastogi 2020 | Short term, high-dose vitamin D supplementation for COVID-19 disease: a randomised, placebo-controlled, study (SHADE study) | Drug treatment |
| Ravikirti 2021 | Ivermectin as a potential treatment for mild to moderate COVID-19 – A double blind randomized placebo-controlled trial | Drug treatment |
| Ray 2020 | Clinical and immunological benefits of convalescent plasma therapy in severe COVID-19: insights from a single center open label randomised control trial | Blood product |
| Reig 2020 | 560. Repurposing Eravacycline for the Treatment of SARS-CoV-2 Infections | Not a randomized trial |
| Reis 2021 | Hydroxychloroquine or Lopinavir/ Ritonavir for Early Treatment of COVID-19 (The TOGETHER Trial): An Adaptive Platform Trial in High-Risk Populations with Symptomatic Disease | Drug treatment |
| Ren 2020 | A Randomized, Open-label, Controlled Clinical Trial of Azvudine Tablets in the Treatment of Mild and Common COVID-19, A Pilot Study | Drug treatment |
| Rocco 2020 | Early use of nitazoxanide in mild Covid-19 disease: randomized, placebo- controlled trial | Drug treatment |
| Rocco 2020 | Early use of nitazoxanide in mild Covid-19 disease: randomized, placebo- controlled trial | Drug treatment |
| Roozbeh 2020 | Sofosbuvir and daclatasvir for the treatment of COVID-19 outpatients: a double-blind, randomized controlled trial | Drug treatment |
| Rosas 2020 | Tocilizumab in Hospitalized Patients With COVID-19 Pneumonia | Drug treatment |
| Ruzhentsova 2020 | Phase 3 Trial of Coronavir (Favipiravir) in patients with mild to moderate COVID-19 | Drug treatment |
| Sadeghi 2020 | Sofosbuvir and daclatasvir compared with standard of care in the treatment of patients admitted to hospital with moderate or severe coronavirus infection (COVID-19): a randomized controlled trial | Drug treatment |
| Sadoff 2020 | Safety and immunogenicity of the Ad26.COV2.S COVID-19 vaccine candidate: interim results of a phase 1/2a, double-blind, randomized, placebo-controlled trial (preprint) | Vaccine |
| Sadoff 2021 | Interim Results of a Phase 1-2a Trial of Ad26.COV2.S Covid-19 Vaccine. | Vaccine |
| Sakoulas 2020 | Intravenous Immunoglobulin (IVIG) Significantly Reduces Respiratory Morbidity in COVID-19 Pneumonia: A Prospective Randomized Trial | Blood product |
| Sakoulas 2020 | Intravenous Immunoglobulin Plus Methylprednisolone Mitigate Respiratory Morbidity in Coronavirus Disease 2019 | Blood product |
| Salama 2020 | Tocilizumab in nonventilated patients hospitalized with Covid-19 pneumonia | Drug treatment |
| Salama 2020 | Tocilizumab in patients hospitalized with Covid-19 pneumonia | Drug treatment |
| Salazar 2020 | Treatment of Coronavirus Disease 2019 (COVID-19) Patients with Convalescent Plasma | Not a randomized trial |
| Salehzadeh 2020 | The Impact of Colchicine on The COVID-19 Patients; A Clinical Trial Study | Drug treatment |
| Salman 2020 | Efficacy and safety of transfusing plasma from COVID-19 survivors to COVID-19 victims with severe illness. A double-blinded controlled preliminary study | Blood product |
| Salvarani 2020 | Effect of Tocilizumab vs Standard Care on Clinical Worsening in Patients Hospitalized With COVID-19 Pneumonia A Randomized Clinical Trial | Drug treatment |
| Sardari 2021 | Therapeutic effect of thyme (Thymus vulgaris) essential oil on patients with covid19: A randomized clinical trial | Traditional Chinese medicine |
| Schumacher 2020 | The impact of respiratory protective equipment on difficult airway management: a randomised, crossover, simulation study | Not exposed to or diagnosed with (confirmed or suspected) COVID-19 |
| Sekhavati 2020 | NSafety and Effectiveness of Azithromycin in Patients with COVID-19: an open-label randomized trial | Drug treatment |
| Sekhavati 2020 | Safety and effectovemess pf azithromycin in patients with COVID-19: An open-label randomised trial | Drug treatment |
| Sekhavati 2020 | Safety and effectovemess pf azithromycin in patients with COVID-19: An open-label randomised trial | Drug treatment |
| Self | Effect of Hydroxychloroquine on Clinical Status at 14 Days in Hospitalized Patients With COVID-19 A Randomized Clinical Trial | Drug treatment |
| Self 2020 | Effect of Hydroxychloroquine on Clinical Status at 14 Days in Hospitalized Patients With COVID-19 A Randomized Clinical Trial | Drug treatment |
| Selman 2020 | Results of a Chilean Nation-Wide Network of Blood Banks for Convalescent Plasma Collection for Pandemic SARS-CoV-2 Treatment | Not a randomized trial |
| Seneviratne 2020 | Efficacy of commercial mouth-rinses on SARS-CoV-2 viral load in saliva: Randomized Control Trial in Singapore (preprint) | Others (e.g. mouthwash) |
| Shah 2020 | Safety and efficacy of ozone therapy in mild to moderate COVID-19 patients: A Phase 1/11 Randomized control Trial (SEOT study) | Oxygen delivery |
| Shaw 2020 | Wearing of Cloth or Disposable Surgical Face Masks has no Effect on Vigorous Exercise Performance in Healthy Individuals | Personal protective equipment |
| Sheng 2020 | Canakinumab to reduce deterioration of cardiac and respiratory function in SARS-CoV-2 associated myocardial injury with heightened inflammation (canakinumab in Covid-19 cardiac injury: The three C study) | Randomized trial with no results (e.g. trial registry) |
| Shi 2020 | Treatment with human umbilical cord-derived mesenchymal stem cells 2 for COVID-19 patients with lung damage: a randomised, double-blind, 3 placebo-controlled phase 2 trial | Blood product |
| Shi 2020 | Treatment with human umbilical cord-derived mesenchymal stem cells 2 for COVID-19 patients with lung damage: a randomised, double-blind, 3 placebo-controlled phase 2 trial | Blood product |
| Shi 2020 | Clinical Observation of the Rehabilitation Formula for Banking up Earth to Generate Metal in Treating COVID-19 Patients with Deficiency of Lung and Spleen Syndrome in the Recovery Stage | Traditional Chinese medicine |
| Shih 2020 | Remdesivir is Effective for Moderately Severe Patients: A Re-Analysis of the First Double-Blind, Placebo-Controlled, Randomized Trial on Remdesivir for Treatment of Severe COVID-19 Patients Conducted in Wuhan City | Drug treatment |
| ShilongYang 2020 | Safety and immunogenicity of a recombinant tandem-repeat dimeric RBD protein vaccine against COVID-19 in adults: pooled analysis of two randomized, double-blind, placebo-controlled, phase 1 and 2 trials | Vaccine |
| Shogenova 2020 | Effect of thermal helium-oxygen mixture on viral load in COVID-19 | Oxygen delivery |
| Shu 2020 | Treatment of severe COVID-19 with human umbilical cord mesenchymal stem cells | Blood product |
| Sigamani 2020 | Galectin antagonist use in mild cases of SARS-CoV-2; pilot feasibility 2 randomised, open label, controlled trial. | Drug treatment |
| Simonovich 2020 | A Randomized Trial of Convalescent Plasma in Covid-19 Severe Pneumonia | Blood product |
| Singh 2020 | In adults exposed to COVID-19, hydroxychloroquine did not reduce confirmed or probable COVID-19;trial stopped for futility | Not a randomized trial |
| Sinha 2020 | Interleukin-6 Receptor Inhibitor Therapy Is Associated with Improved Outcomes in Patients with Severe COVID-19 Disease (preprint) | Not a randomized trial |
| Sivapalan 2020 | Proactive prophylaxis with azithromycin and hydroxychloroquine in hospitalized patients with COVID-19 (ProPAC-COVID): a statistical analysis plan | Randomized trial with no results (e.g. trial registry) |
| Skipper 2020 | Hydroxychloroquine in nonhospotalized adults with early COVID-19 | Drug treatment |
| Spinner 2020 | Effect of Remdesivir vs Standard Care on Clinical Status at 11 Days in Patients With Moderate COVID-19 A Randomized Clinical Trial | Drug treatment |
| Spoorthi 2020 | Utility of Ivermectin and Doxycycline combination for the treatment of SARS-CoV-2 | Not a randomized trial |
| Stone 2020 | Efficacy of Tocilizumab in Patients Hospitalized with Covid-19 | Drug treatment |
| Strohbehn 2020 | COVIDOSE: A phase 2 clinical trial of low-dose tocilizumab in the treatment of non-critical COVID-19 pneumonia. | Not a randomized trial |
| Strohbehn 2020 | COVIDOSE: Low-dose tocilizumab in the treatment of COVID-19 pneumonitis | Randomized trial with no results (e.g. trial registry) |
| Sun 2020 | Clinical value of prophylactic liver protecting drugs in coronavirus disease 2019 (COVID -19) | Drug treatment |
| Sun 2020 | Study on the clinical efficacy of Lianhua Qingke Granules in the treatment of mild and common new coronavirus pneumonia | Traditional Chinese medicine |
| Suofang 2020 | Treatment of 30 Cases of Qi and Yin Deficiency Syndrome in the Recovery Period of New Coronavirus Pneumonia by Comprehensive Therapy of Traditional Chinese Medicine | Traditional Chinese medicine |
| Suppan 2020 | Effect of an E-Learning Module on Personal Protective Equipment Proficiency by Prehospital Personnel: Web-Based, Randomized Controlled Trial | Psychological and educational |
| Tabarsi 2020 | Evaluating the effects of Intravenous Immunoglobulin (IVIg) on the management of severe COVID-19 cases: A randomized controlled trial | Blood product |
| Tahmasebi 2020 | Immunomodulatory effects of nanocurcumin on Th17 cell responses in mild and severe COVID‐19 patients | Drug treatment |
| Takayama 2020 | A multi-center, randomized controlled trial by the Integrative Management in Japan for Epidemic Disease (IMJEDI study-RCT) on the use of Kampo medicine, kakkonto with shosaikotokakikyosekko, in mild-to-moderate COVID-19 patients for symptomatic relief and | Randomized trial with no results (e.g. trial registry) |
| Tan 2020 | Effect of Intravenous Lidocaine on Central Venous Puncture Analgesia in Cases of Suspected or Confirmed COVID-19 | Others (e.g. mouthwash) |
| Tang 2020 | Hydroxychloroquine in patients with COVID-19: an open-label, randomized, controlled trial | Drug treatment |
| Tang 2020 | Hydroxychloroquine in patients with mainly mild to moderate coronavirus disease 2019: open label, randomised controlled trial | Drug treatment |
| Tomazini 2020 | Effect of Dexamethasone on Days Alive and Ventilator-Free in Patients With Moderate or Severe Acute Respiratory Distress Syndrome and COVID-19 The CoDEX Randomized Clinical Trial | Drug treatment |
| Toniati 2020 | Tocilizumab for the treatment of severe COVID-19 pneumonia with hyperinflammatory syndrome and acute respiratory failure: A single center study of 100 patients in Brescia, Italy | Not a randomized trial |
| Torres Zambrano 2020 | Features and outcomes of secondary sepsis and urinary tract infections in COVID-19 patients treated with stem cell nebulization | Blood product |
| Udwadia 2020 | Efficacy and Safety of Favipiravir, an Oral RNA-Dependent RNA Polymerase Inhibitor, in Mild-to-Moderate COVID-19: A Randomized, Comparative, Open-Label, Multicenter, Phase 3 Clinical Trial | Drug treatment |
| Udwadia 2020 | Efficacy and Safety of Favipiravir, an Oral RNA-Dependent RNA Polymerase Inhibitor, in Mild-to-Moderate COVID-19: A Randomized, Comparative, Open-Label, Multicenter, Phase 3 Clinical Trial | Drug treatment |
| Ulrich 2020 | TREATING COVID-19 WITH HYDROXYCHLOROQUINE (TEACH): A MULTICENTER, DOUBLE-BLIND, RANDOMIZED CONTROLLED TRIAL IN HOSPITALIZED PATIENTS | Drug treatment |
| Vaira 2020 | Efficacy of corticosteroid therapy in the treatment of long- lasting olfactory disorders in COVID-19 patients* | Drug treatment |
| Valizadeh 2020 | Nano-curcumin therapy, a promising method in modulating inflammatory cytokines in COVID-19 patients | Drug treatment |
| Villa 2020 | Blood purification therapy with a hemodiafilter featuring enhanced adsorptive properties for cytokine removal in patients presenting COVID-19: a pilot study | Not a randomized trial |
| Villar 2020 | Dexamethasone treatment for the acute respiratory distress syndrome: a multicentre, randomised controlled trial | Not exposed to or diagnosed with (confirmed or suspected) COVID-19 |
| Vlaar 2020 | Anti-C5a antibody (IFX-1) treatment of severe Covid-19: an exploratory phase 2 randomized controlled trial | Drug treatment |
| Vlaar 2020 | Anti-C5a antibody (IFX-1) treatment of severe Covid-19: an exploratory phase 2 randomized controlled trial | Drug treatment |
| Voysey 2020 | Safety and efficacy of the ChAdOx1 nCoV-19 vaccine (AZD1222) against SARS-CoV-2: an interim analysis of four randomised controlled trials in Brazil, South Africa, and the UK | Vaccine |
| Walsh 2020 | Safety and Immunogenicity of Two RNA-Based Covid-19 Vaccine Candidates. | Vaccine |
| Walsh 2020 | RNA-Based COVID-19 Vaccine BNT162b2 Selected for a Pivotal Efficacy Study (preprint) | Vaccine |
| Wan 2020 | Influence of Baduanjin on the health effects of patients with cold-dampness-stagnation type of new coronavirus pneumonia | Exercise/rehabilitation |
| Wang 2020 | Preliminary clinical effect analysis of the treatment of novel coronavirus pneumonia by internal administration of traditional Chinese medicine plus fumigation and absorption combined with super dose of vitamin C in treating NOVID-19 | Drug treatment |
| Wang 2020 | Remdesivir in adults with severe COVID-19: a randomised, double-blind, placebo-controlled, multicentre trial | Drug treatment |
| Wang 2020 | Remdesivir in adults with severe COVID-19: a randomised, double-blind, placebo-controlled, multicentre trial | Drug treatment |
| Wang 2020 | Preliminary clinical effect analysis of the treatment of novel coronavirus pneumonia by internal administration of traditional Chinese medicine plus fumigation and absorption combined with super dose of vitamin C in treating NOVID-19 | Drug treatment |
| Wang 2020 | Lianhua Qingwen capsule and interferon-α combined with lopinavir /ritonavir for the treatment of 30 COVID-19 patients | Drug treatment |
| Wang 2020 | Tocilizumab ameliorates the hypoxia in COVID-19 moderate patients with bilateral pulmonary lesions: a randomized, controlled, open-label, multicenter trial | Drug treatment |
| Wang 2020 | Treatment of COVID-19 Patients with Prolonged Post-Symptomatic Viral Shedding with Leflunomide -- a Single-Center, Randomized, Controlled Clinical Trial | Drug treatment |
| Wang 2020 | Preliminary clinical effect analysis of the treatment of novel coronavirus pneumonia by internal administration of traditional Chinese medicine plus fumigation and absorption combined with super dose of vitamin C in treating NOVID-19 | Drug treatment |
| Wang 2020 | Tocilizumab Ameliorates the Hypoxia in COVID-19 Moderate Patients with Bilateral Pulmonary Lesions: A Randomized, Controlled, Open-Label, Multicenter Trial | Drug treatment |
| Wang 2020 | Efficacy and Safety of Leflunomide for Refractory COVID-19: An Open-label Controlled Study | Not a randomized trial |
| Wang 2020 | Exploring an Integrative Therapy for Treating COVID-19: A Randomized Controlled Trial | Traditional Chinese medicine |
| Wang 2020 | Preliminary clinical efficacy analysis of oral Chinese medicine plus fumigation combined with super high dose vitamin C in the treatment of new coronavirus pneumonia | Traditional Chinese medicine |
| Wang 2020 | Clinical study on the treatment of patients with novel coronavirus pneumonia and asymptomatic infection with integrated traditional Chinese and western medicine | Traditional Chinese medicine |
| Wanjarkhedkar 2020 | A prospective clinical study of an Ayurveda regimen in COVID 19 patients | Traditional Chinese medicine |
| Ward 2020 | Phase 1 trial of a Candidate Recombinant Virus-Like Particle Vaccine for Covid-19 Disease Produced in Plants | Vaccine |
| Wei 2020 | A multi-center, prospective study of early abidol + lopinavir/ritonavir + recombinant interferon α-2b combined antiviral therapy in patients with novel coronavirus pneumonia in Zhejiang Province | Not a randomized trial |
| Weijin 2020 | Clinical Observation of Tanshinone ⅡA Sulfonate in the Treatment of New Coronary Pneumonia | Drug treatment |
| Weinreich 2020 | REGN-COV2, a Neutralizing Antibody Cocktail, in Outpatients with Covid-19 | Blood product |
| Wen 2020 | [Effect of Xuebijing injection on inflammatory markers and disease outcome of coronavirus disease 2019] | Traditional Chinese medicine |
| WHO REACT Working Group 2020 | Association Between Administration of Systemic Corticosteroids and Mortality Among Critically Ill Patients With COVID-19 A Meta-analysis | Drug treatment |
| Wu 2020 | Efficacy and safety of triazavirin therapy for coronavirus disease 2019: A pi‐ lot randomized controlled trial | Drug treatment |
| Wu 2020 | Identification and validation of a novel clinical signature to predict the prognosis in confirmed COVID-19 patients | Not a randomized trial |
| Wu 2020 | Phase 1 trial for treatment of COVID-19 patients with pulmonary fibrosis using hESC-IMRCs | Not a randomized trial |
| Wu 2020 | Effect of emotional care based on traditional Chinese medicine combining respiratory muscle training on depression and anxiety among patients with COVID-19 | Traditional Chinese medicine |
| Xia 2020 | Effect of an Inactivated Vaccine Against SARS-CoV-2 on Safety and Immunogenicity Outcomes: Interim Analysis of 2 Randomized Clinical Trials | Vaccine |
| Xia 2020 | Safety and immunogenicity of an inactivated SARS-CoV-2 vaccine, BBIBP-CorV: a randomised, double-blind, placebo-controlled, phase 1/2 trial | Vaccine |
| Xiao 2020 | Analysis of the value of traditional Chinese medicine Shufeng Jiedu Capsule combined with Arbidol in the treatment of mild new coronavirus pneumonia | Not a randomized trial |
| Xiao 2020 | Efficacy of Huoxiang Zhengqi dropping pills and Lianhua Qingwen granules in treatment of COVID-19: A randomized controlled trial | Traditional Chinese medicine |
| Xiasai 2020 | Comparison of clinical effects of conventional nasal catheter oxygen inhalation and high-flow oxygen inhalation in the treatment of new coronavirus pneumonia | Oxygen delivery |
| Xie 2020 | Effect of regular intravenous immunoglobulin therapy on prognosis of severe pneumonia in patients with COVID-19 | Not a randomized trial |
| Xiong 2020 | Efficacy of herbal medicine (Xuanfei Baidu decoction) combined with conventional drug in treating COVID-19:A pilot randomized clinical trial | Not a randomized trial |
| Yadegarinia 2020 | Evaluation of the efficacy of arbidol in comparison with the standard treatment regimen of hospitalized patients with COVID-19: A randomized clinical trial | Drug treatment |
| Yakoot 2020 | Efficacy and Safety of Sofosbuvir/Daclatasvir in the Treatment of COVID19 A randomized, controlled study | Drug treatment |
| Yali 2020 | Preliminary clinical efficacy analysis of traditional Chinese medicine taken orally plus smoked inhalation combined with super-high dose vitamin C in the treatment of new coronavirus pneumonia | Others (e.g. mouthwash) |
| Yan 2020 | Large-scale prospective clinical study on prophylactic intervention of COVID-19 in community population using Huoxiang Zhengqi Oral Liquid and Jinhao Jiere Granules. [Chinese] | Traditional Chinese medicine |
| Yan 2020 | Large-sample prospective clinical study on the preventive intervention of COVID-19 in the community by the combined use of Huoxiangzhengqi oral liquid and Jinhaojiere granules | Traditional Chinese medicine |
| Yan 2020 | Effects and Safety of Herbal Medicines among Community-dwelling Residents during COVID-19 Pandemic: A Large Prospective, Randomized Controlled Trial (RCT) | Traditional Chinese medicine |
| Yang 2020 | Clinical Characteristics and Outcomes of COVID-19 Patients Receiving Compassionate Use Leronlimab | Not a randomized trial |
| Ye 2020 | Clinical efficacy of lopinavir/ritonavir in the treatment of Coronavirus disease 2019 | Not a randomized trial |
| Ye 2020 | Guideline-based Chinese herbal medicine treatment plus standard care for severe coronavirus disease 2019 (G-CHAMPS): evidence from China | Traditional Chinese medicine |
| Yethindra 2020 | Efficacy of umifenovir in the treatment of mild and moderate COVID-19 patients | Drug treatment |
| Younie 2020 | Improving young children&#039;s handwashing behaviour and understanding of germs: The impact of A Germ&#039;s Journey educational resources in schools and public spaces. | Psychological and educational |
| Yu 2020 | Effects of Lianhua Qingwen Granules Plus Arbidol on Treatment of Mild Corona Virus Disease-19. [Chinese] | Traditional Chinese medicine |
| Yu 2020 | Efficacy of Lianhua Qingwen Granules combined with Arbidol in the treatment of mild novel coronavirus pneumonia | Traditional Chinese medicine |
| Yuan 2020 | Pulmonary radiological change of COVID-19 patients with 99mTc-MDP treatment | Drug treatment |
| Zeng 2020 | Comparative effectiveness and safety of ribavirin plus interferon-alpha, lopinavir/ritonavir plus interferon-alpha and ribavirin plus lopinavir/ritonavir plus interferon-alphain in patients with mild to moderate novel coronavirus pneumonia | Randomized trial with no results (e.g. trial registry) |
| Zha 2020 | Corticosteroid treatment of patients with coronavirus disease 2019 (COVID-19) | Not a randomized trial |
| Zhang 2020 | High-dose vitamin C infusion for the treatment of critically ill COVID-19 | Drug treatment |
| Zhang 2020 | Pilot Trial of High-dose vitamin C in critically ill COVID-19 patients | Drug treatment |
| Zhang 2020 | Berberine reduces circulating inflammatory mediators in patients with severe COVID-19 | Drug treatment |
| Zhang 2020 | A comparative study on the time to achieve negative nucleic acid testing and hospital stays between Danoprevir and Lopinavir/Ritonavir in the treatment of patients with COVID-19 | Not a randomized trial |
| Zhang 2020 | Clinical study on the treatment of new type of coronavirus pneumonia (COVID-19) from the perspective of "epidemic virus" | Traditional Chinese medicine |
| Zhang 2020 | Analysis of Clinical Efficacy of Honeysuckle Oral Liquid in Treating 80 Cases of New Coronavirus Pneumonia | Traditional Chinese medicine |
| Zhang 2020 | Immunogenicity and Safety of a SARS-CoV-2 Inactivated Vaccine in Healthy Adults Aged 18-59 years: Report of the Randomized, Double-blind, and Placebo-controlled Phase 2 Clinical Trial (preprint) | Vaccine |
| Zhang 2020 | Safety, tolerability, and immunogenicity of an inactivated SARS-CoV-2 vaccine in healthy adults aged 18-59 years: a randomised, double-blind, placebo-controlled, phase 1/2 clinical trial. | Vaccine |
| Zhang 2021 | High-dose vitamin C infusion for the treatment of critically ill COVID-19 | Drug treatment |
| Zhao 2020 | Tocilizumab combined with favipiravir in the treatment of COVID-19: A multicenter trial in a small sample size | Drug treatment |
| Zhao 2020 | Tocilizumab combined with favipiravir in the treatment of COVID-19: A multicenter trial in a small sample size | Drug treatment |
| Zhao 2020 | Yidu-toxicity blocking lung decoction ameliorates inflammation in severe pneumonia of SARS-COV-2 patients with Yidu-toxicity blocking lung syndrome by eliminating IL-6 and TNF-a | Traditional Chinese medicine |
| Zhdanov 2020 | Clinical efficacy and safety of nebulized prostacyclin in patients with sARs-CoV-2 (prospective comparative study). [Russian] | Not a randomized trial |
| Zheng 2020 | A Novel Protein Drug, Novaferon, as the Potential Antiviral Drug for COVID-19 | Drug treatment |
| Zheng 2020 | SARS-CoV-2 Clearance in COVID-19 Patients with Novaferon Treatment: A Randomized, Open-label, Parallel Group Trial | Drug treatment |
| Zhou 2020 | Diamine glycyrrhizinate in common COVID-19 patients. Clinical value in treatment | Drug treatment |
| Zhou 2020 | Diamine glycyrrhizinate in common COVID-19 patients.  Clinical value in treatment | Drug treatment |
| Zhou 2020 | Interferon-a2b treatment for COVID-19 | Not a randomized trial |
| Zhou 2020 | Low-dose corticosteroid combined with immunoglobulin reverses deterioration in severe cases with COVID-19. | Not a randomized trial |
| Zhu 2020 | Arbidol Monotherapy is Superior to Lopinavir/ritonavir in Treating COVID-19 | Not a randomized trial |
| Zhu 2020 | Study on the application value of psychological nursing in pregnant women suspected of new coronavirus pneumonia | Psychological and educational |

**Additional study characteristics and outcome data for each study.**

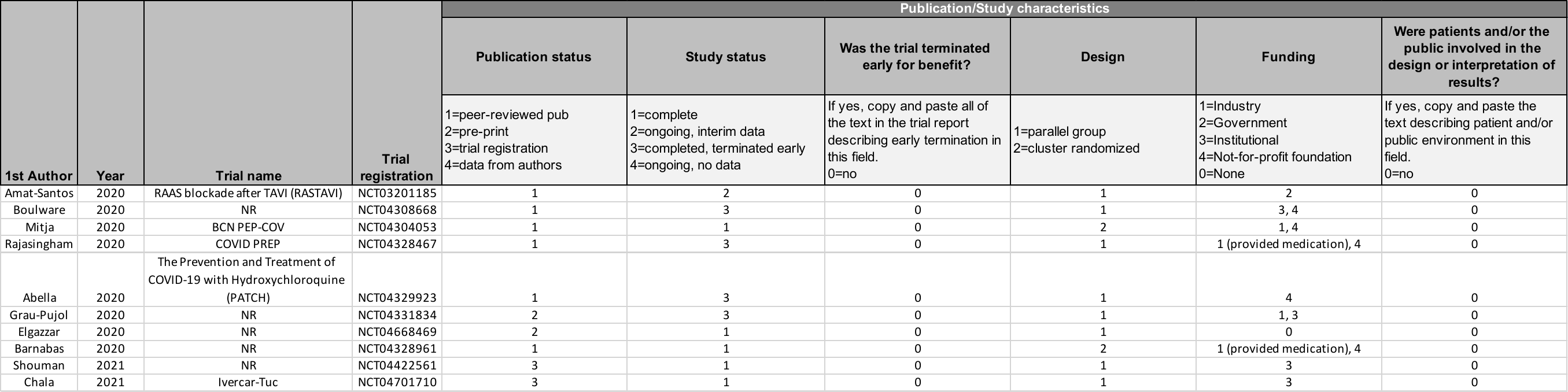

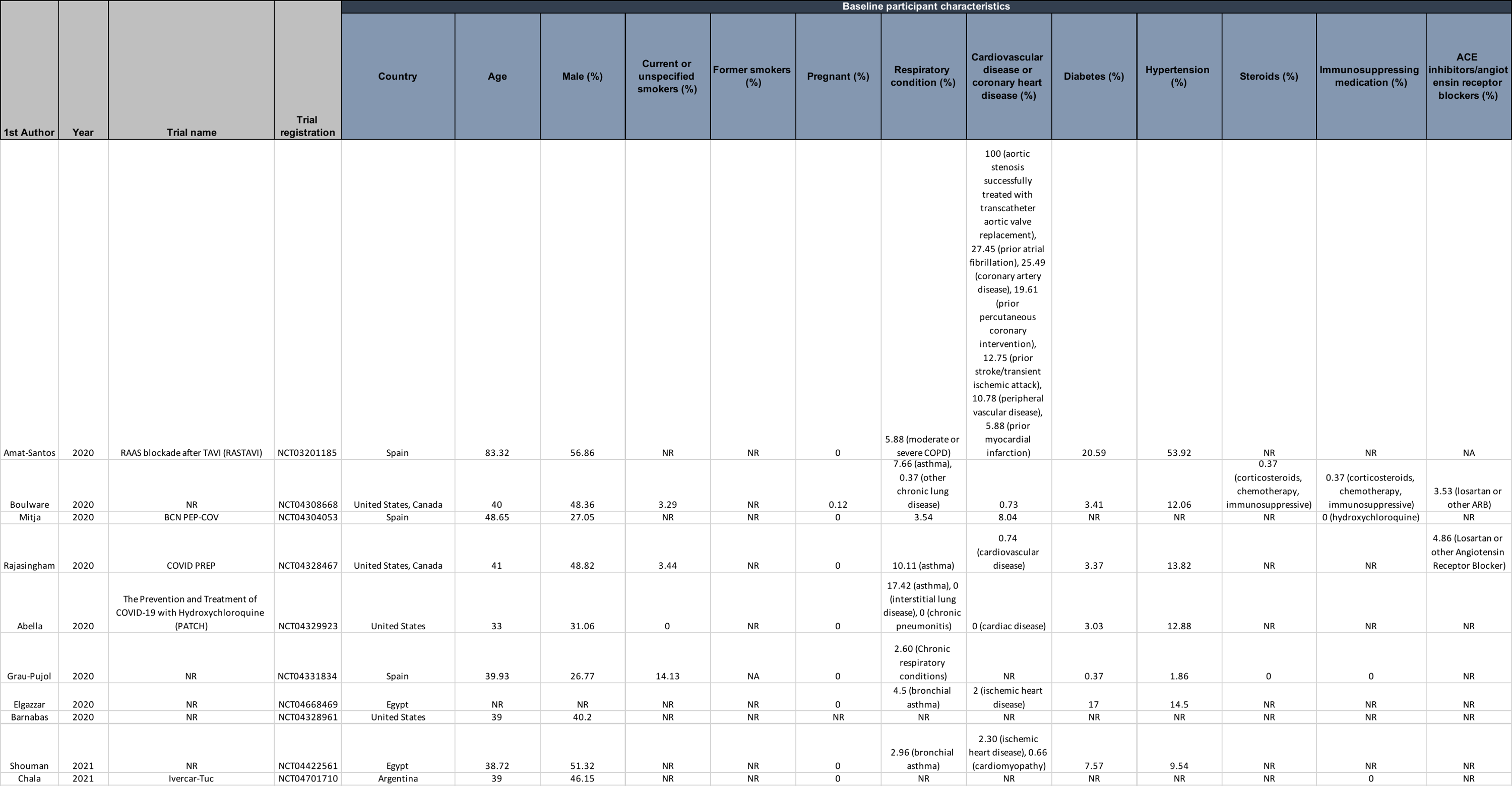

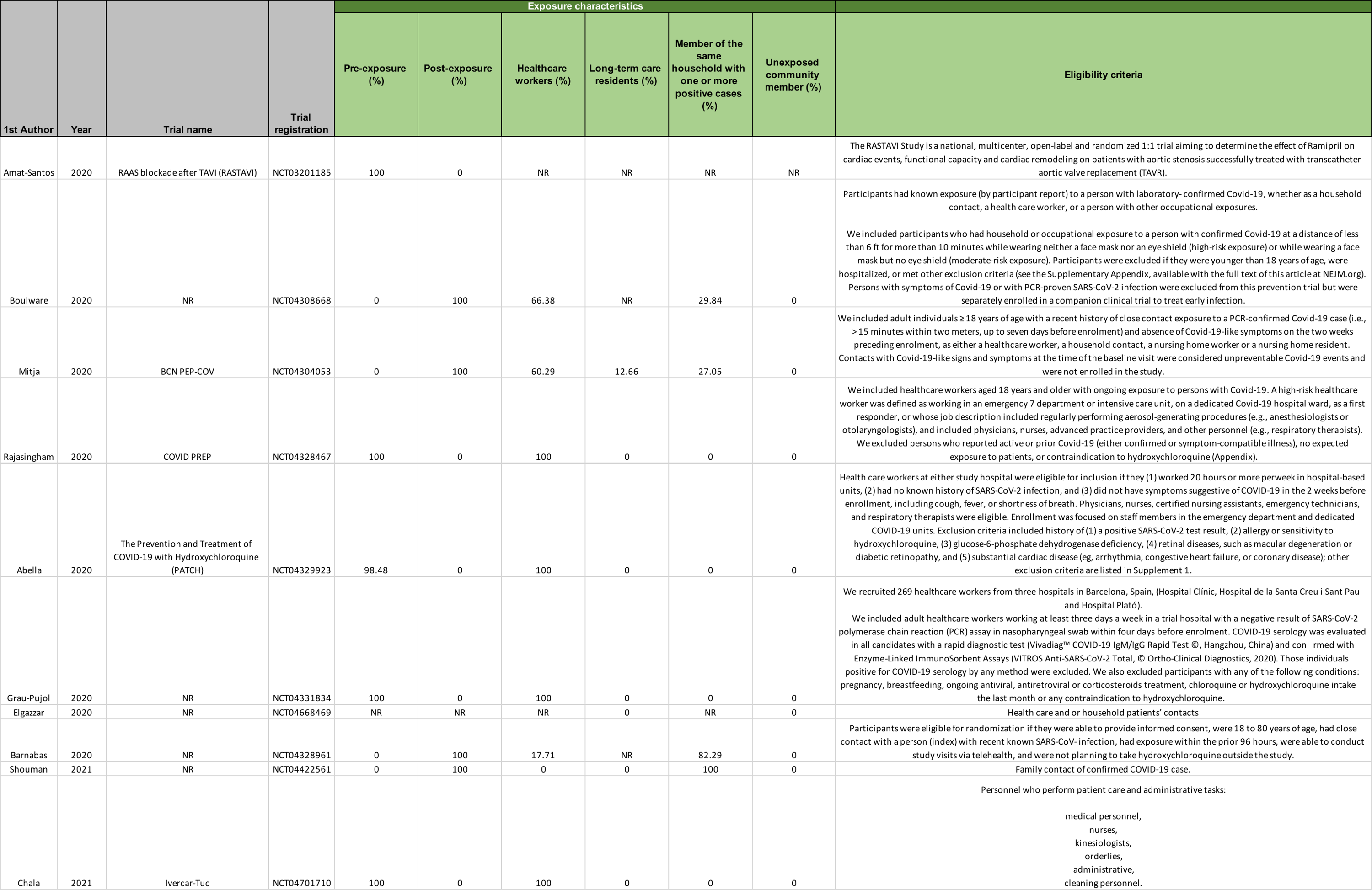

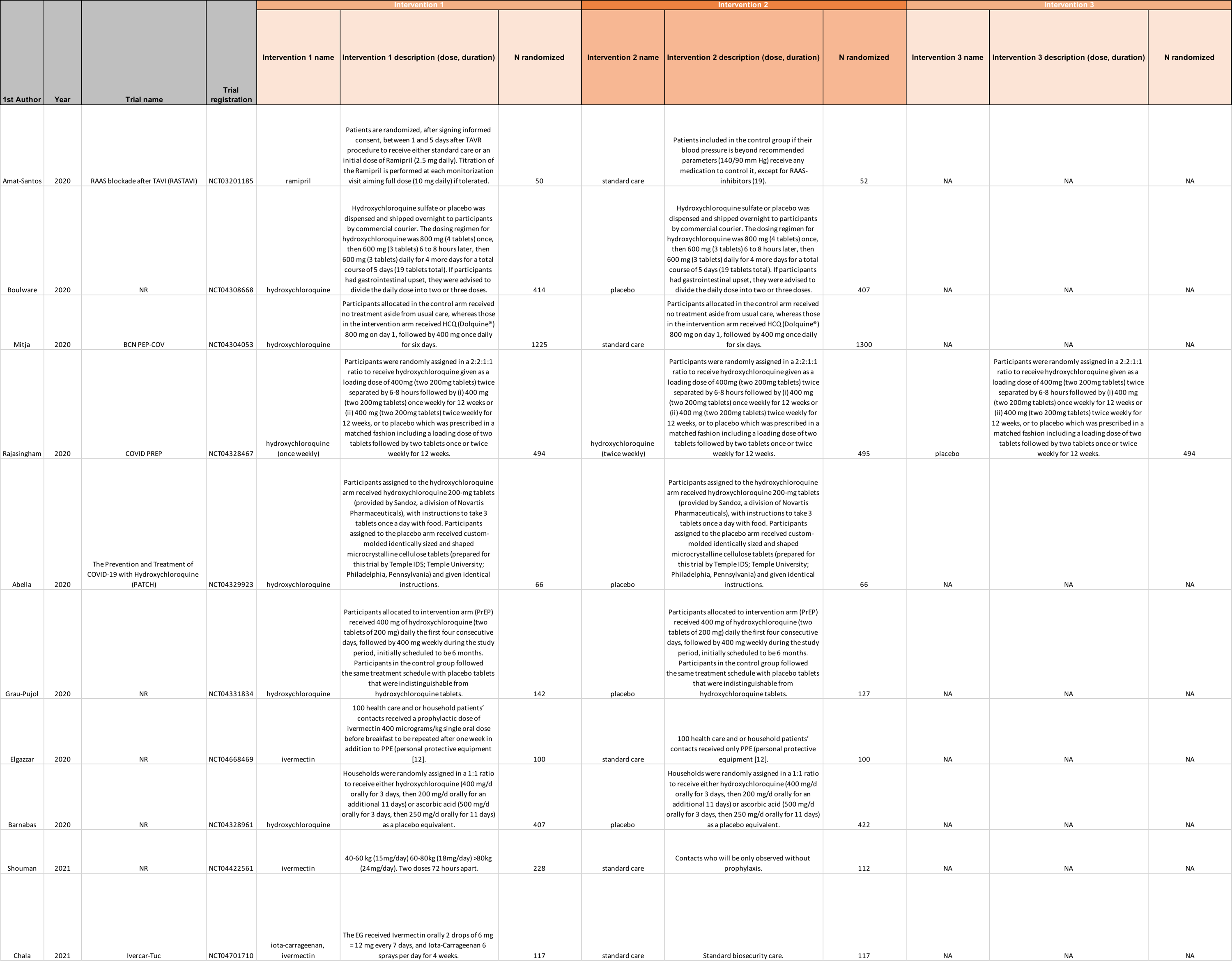

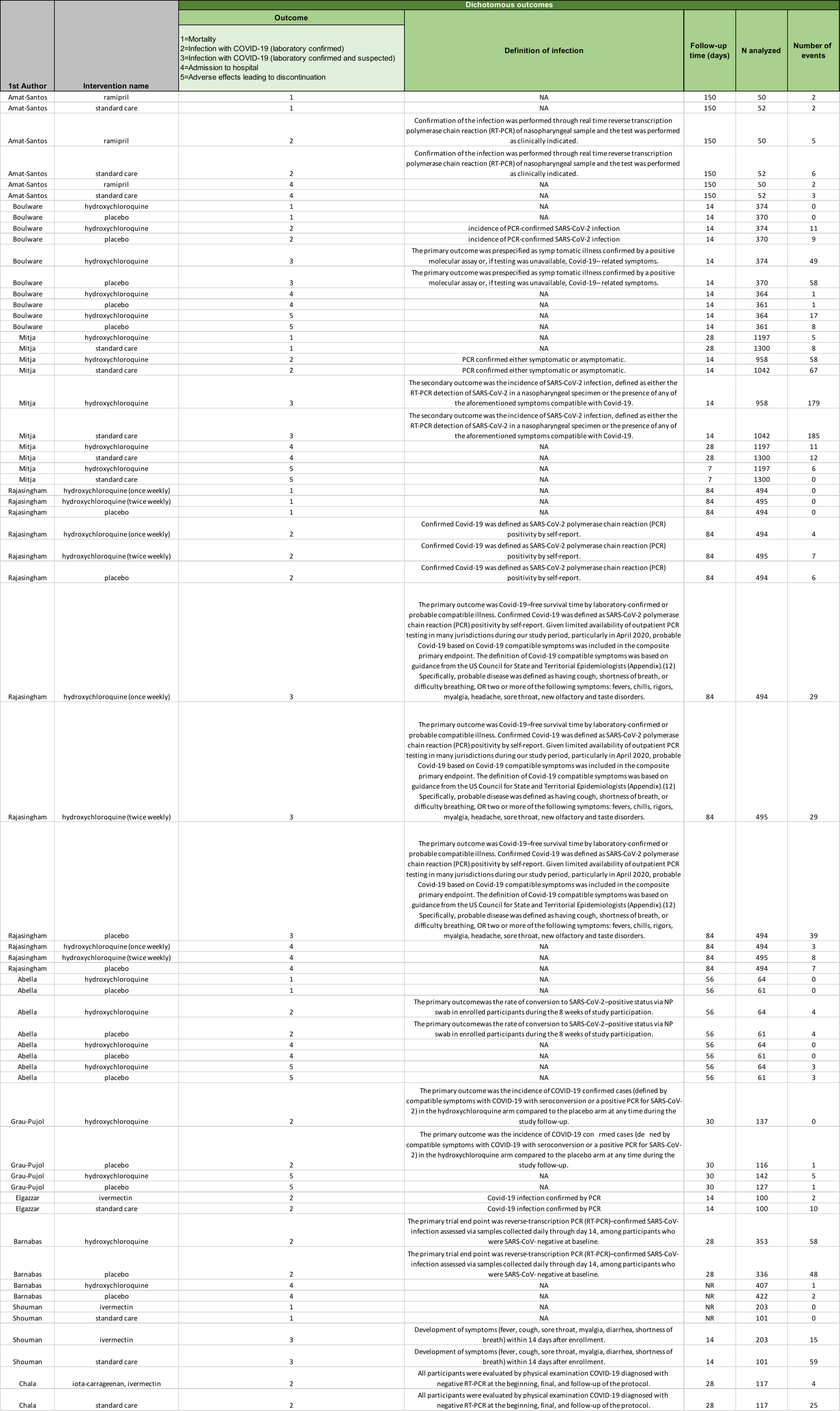

**Table of reporting differences between versions of study preprints**

| **Study** | **Differences between study preprint versions and/or peer-reviewed publications** |
| --- | --- |
| Elgazzar | The latest version of the preprint reports a trial registration whereas the first version of the preprint does not.  *Baseline characteristics* The latest version of the preprint reports mean age and proportion with respiratory conditions, cardiovascular conditions, diabetes, and hypertension. The first version of the preprint does not report these baseline characteristics.  *Risk of bias* The latest version of the preprint reports that the study is double blind whereas the first version of the preprint has no statement on blinding. We kept our risk of bias rating for bias due to deviations from the intended intervention at probably high risk of bias since the study does not mention that it is placebo-controlled. Therefore, blinding of patients and health care providers was likely compromised. The latest version of the preprint reports a method of randomization whereas the first version of the preprint does not. We kept our risk of bias rating for bias arising from the randomization process at probably high risk of bias since the study does not mention allocation concealment. |
| Mitja; BCN PEP-COV | *Baseline characteristics* Number of reported household contacts differ between the preprint and the peer-reviewed publications. Proportion of patients with cardiovascular disease differ between the preprint and the peer-reviewed publications. Proportion of patients with respiratory conditions differ between the preprint and the peer-reviewed publications.  *Results* Number of patients that discontinued due to adverse events differ between the preprint and the peer-reviewed publications.  *Risk of bias* The peer-reviewed publication includes a protocol in the supplementary appendix. We initially rated two outcomes at probably high risk of bias for selective reporting due to differences in the reported and pre-specified time points in the trial registration. After reviewing the protocol in the supplementary appendix, we revised these risk of bias ratings to definitely low risk of bias since the timepoints between the peer-reviewed publications and the protocol are consistent. |
| Rajasingham; COVID PREP | *Baseline characteristics* The preprint includes supplementary material reporting cardiovascular disease, diabetes, and use of ACE inhibitors/angiotensin receptor blockers whereas the supplementary material is not included in the peer-reviewed publication.  *Risk of bias* The peer-reviewed publication includes the protocol as a supplementary file whereas the preprint does not. The protocol prespecified confirmed and suspected infection whereas the trial registration available in the preprint does not. The risk of bias rating for this outcome changed from probably low risk of bias to definitely low risk of bias for selective reporting. |

**Risk of bias assessments for each study and outcome**

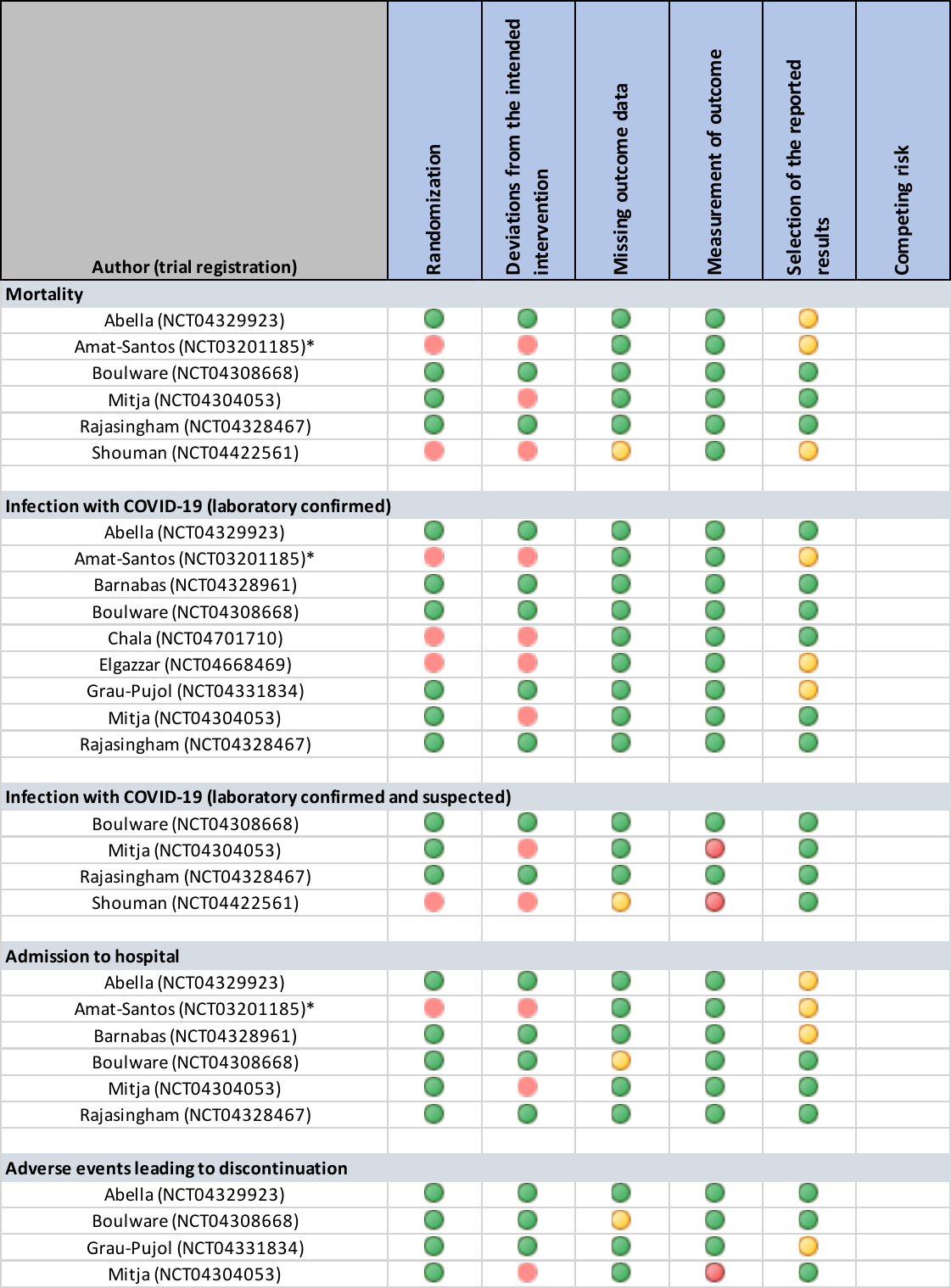

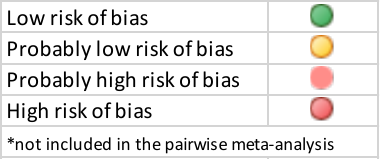

**Network plots**

For all of the following network plots, the width of the lines is proportional to the inverse of the variance of the effect estimate for that comparison (i.e., the wider the line, the narrower the credible interval).

*Laboratory-confirmed SARS-CoV-2 infection*

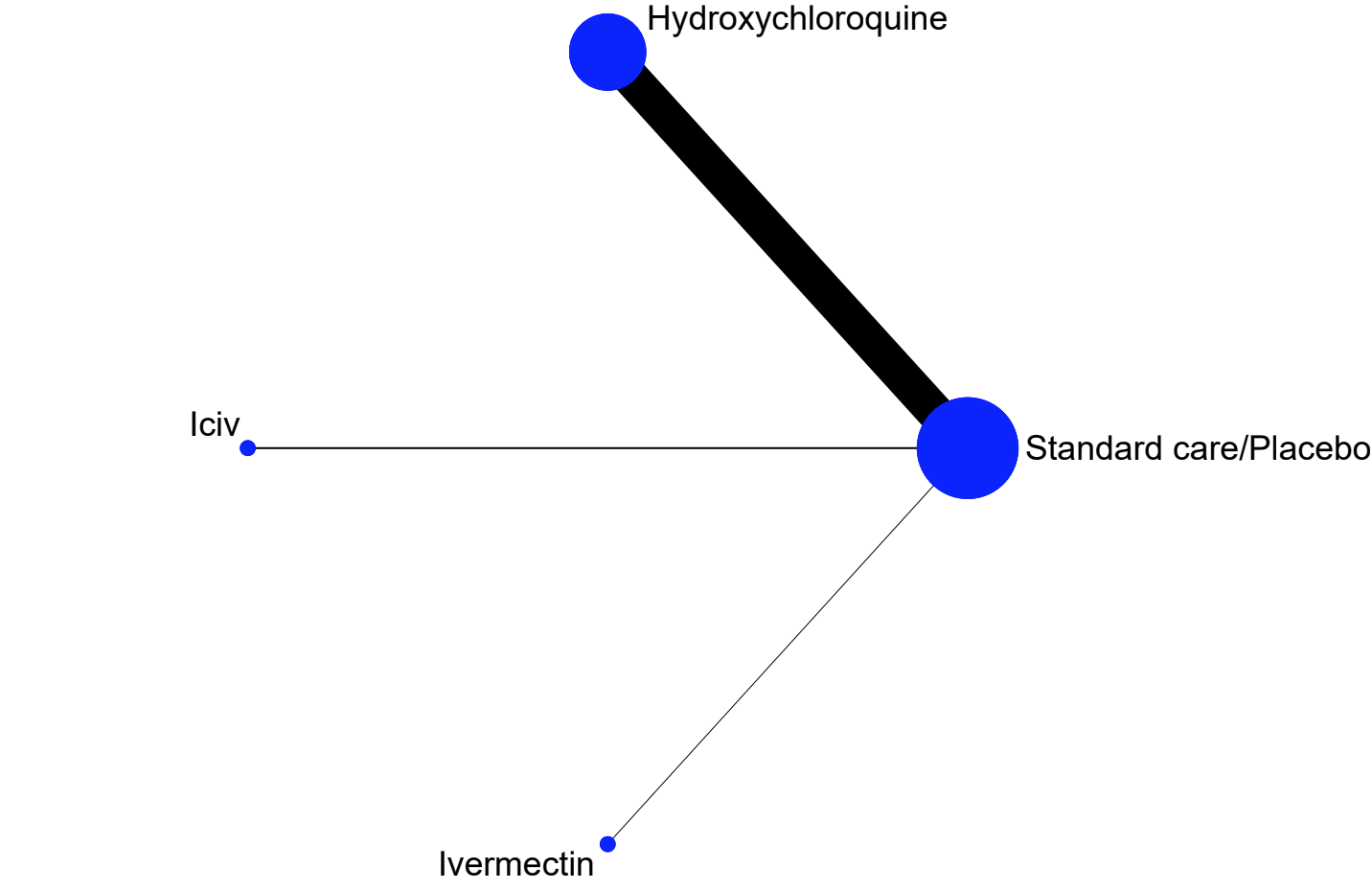

*Suspected, probable or laboratory-confirmed SARS-CoV-2 infection*

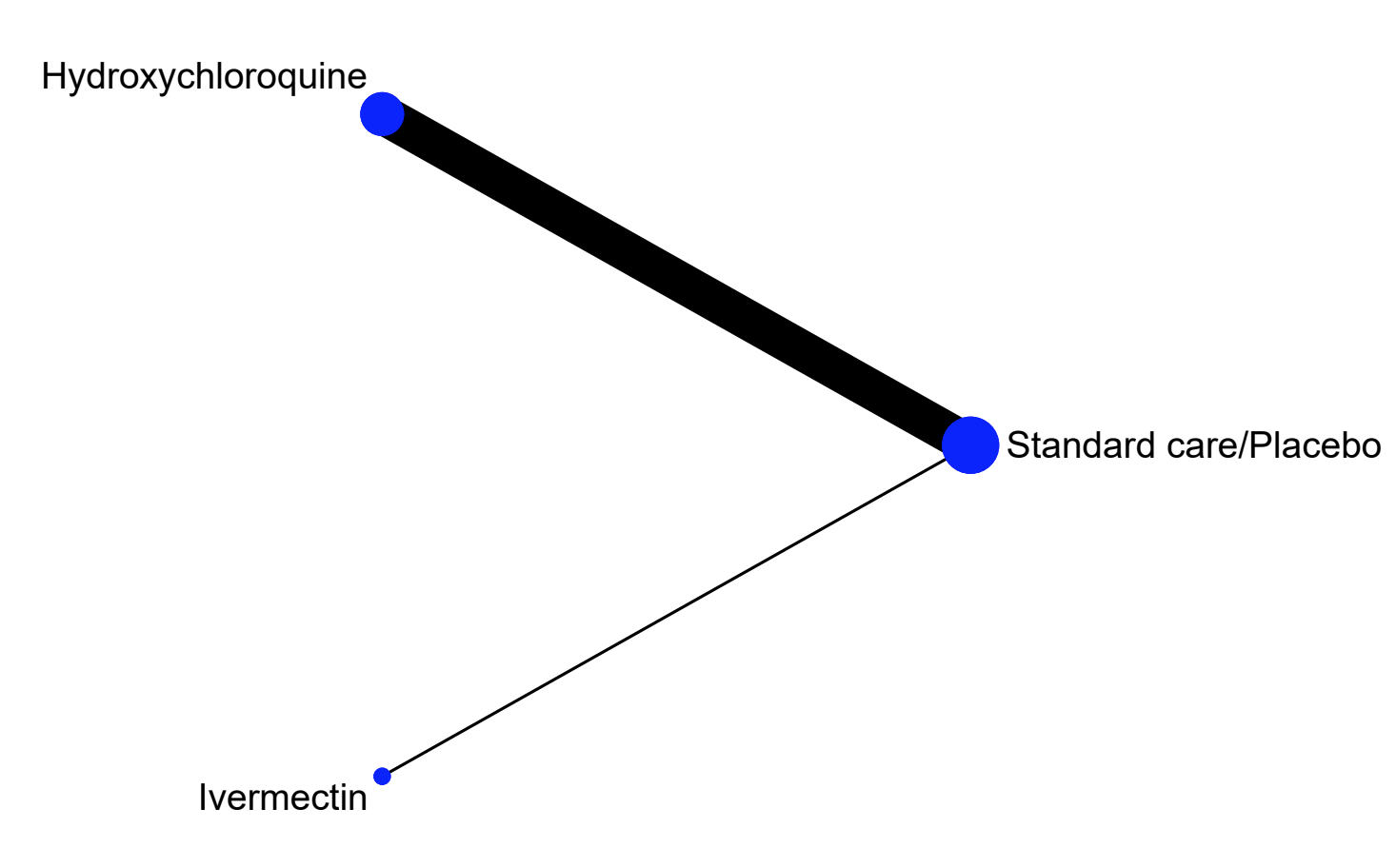

**Forest plots by outcome**

*Laboratory-confirmed SARS-CoV-2 infection*

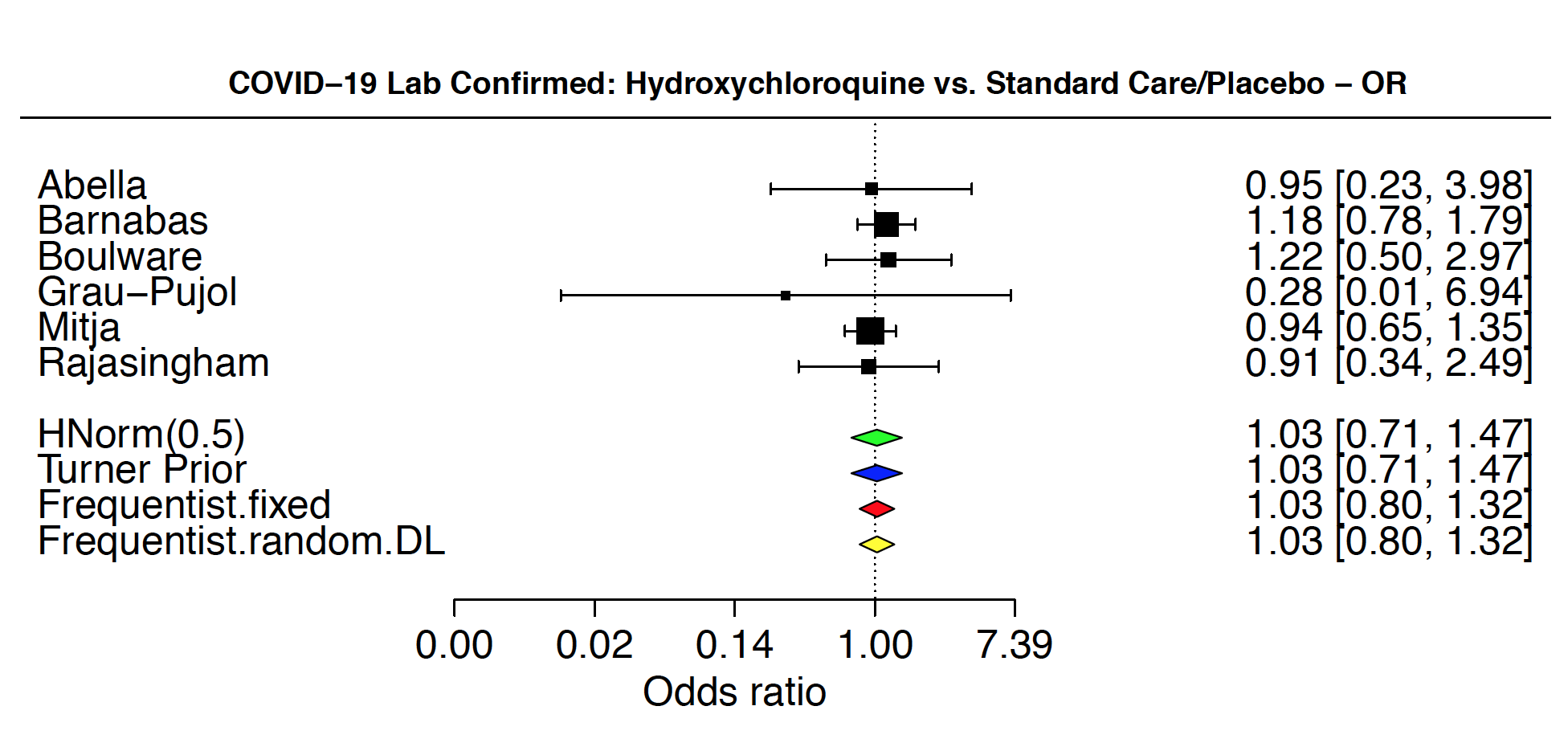

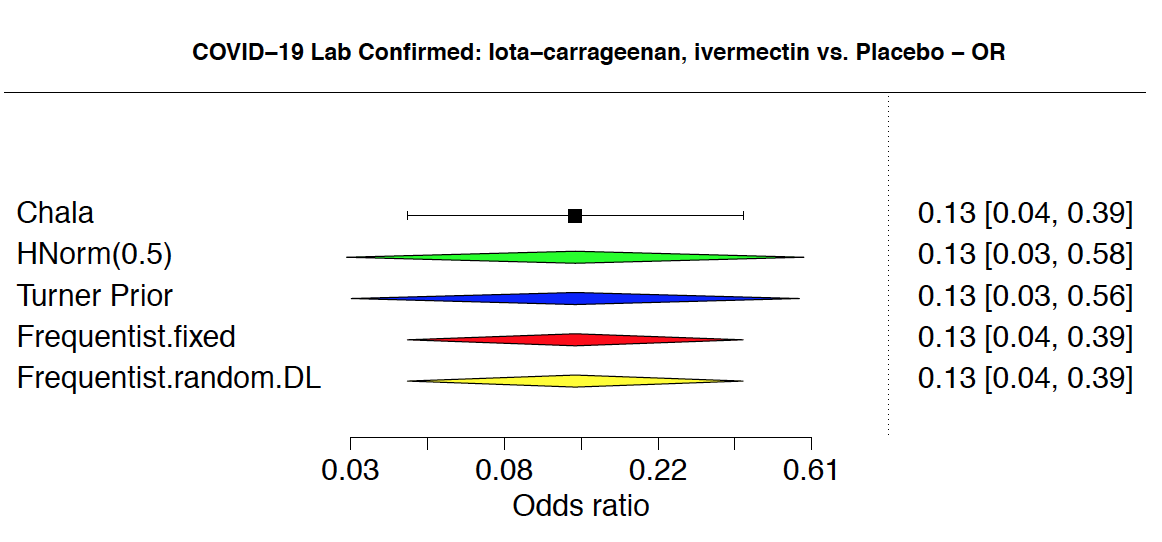

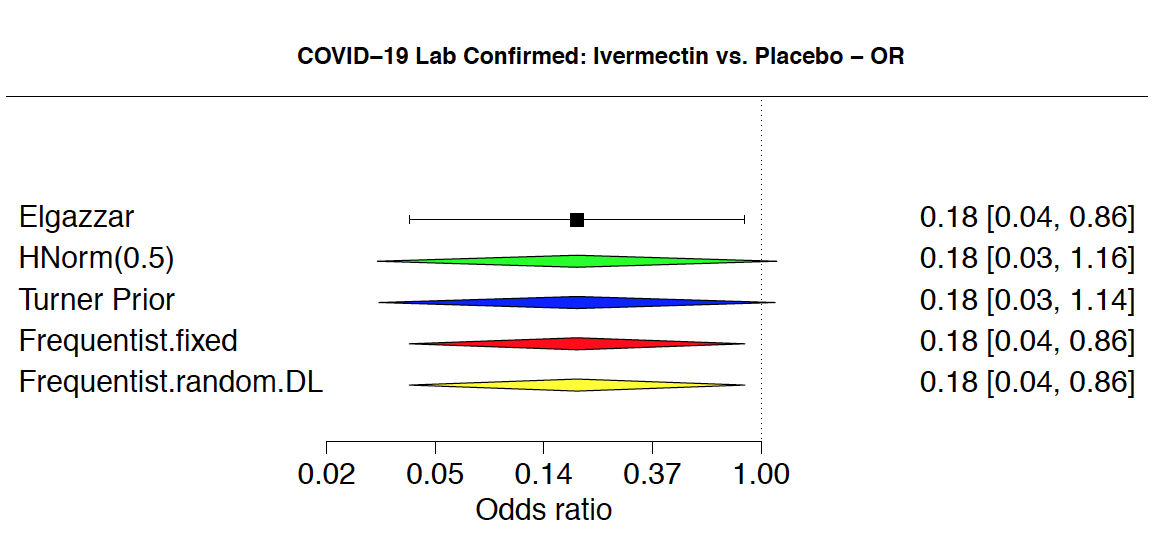

*Suspected, probable or laboratory-confirmed SARS-CoV-2 infection*

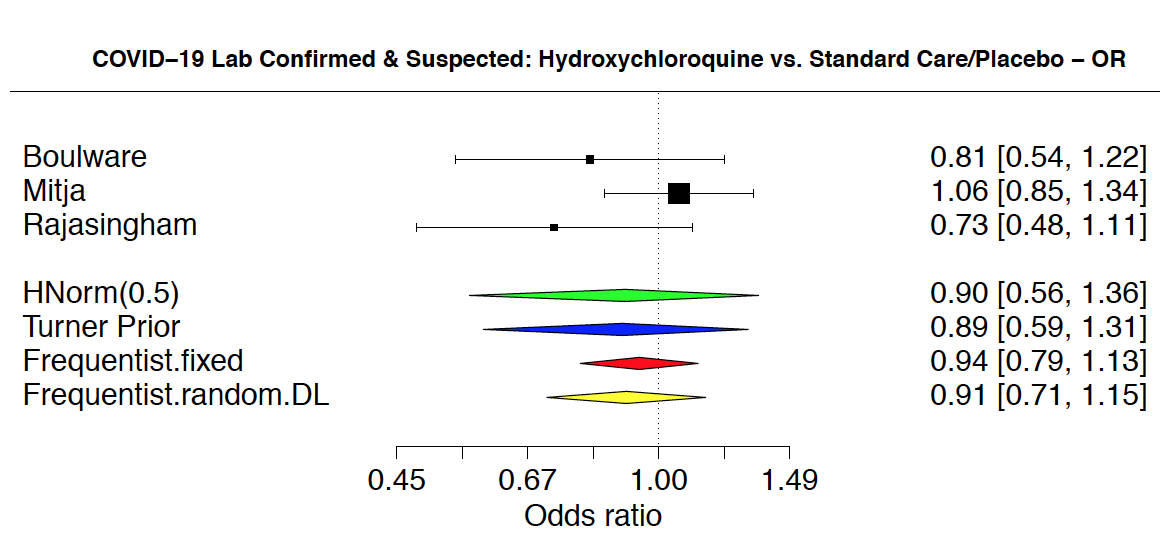

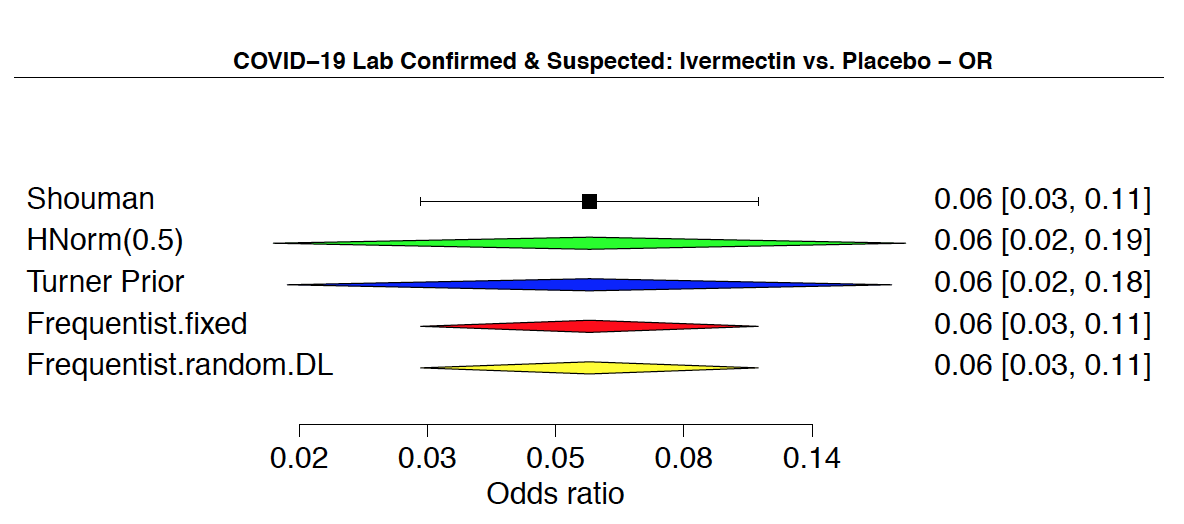

*Admission to hospital*

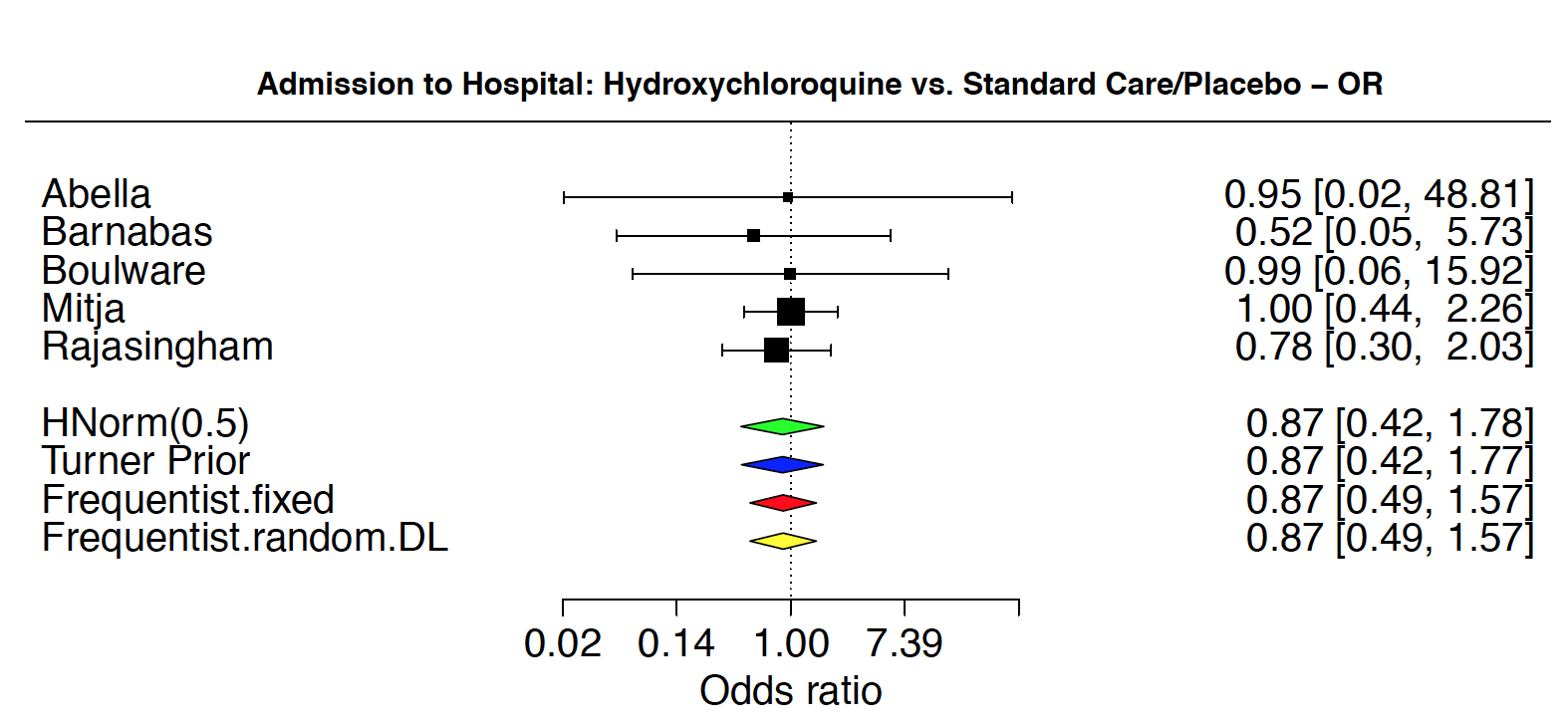

*Mortality*

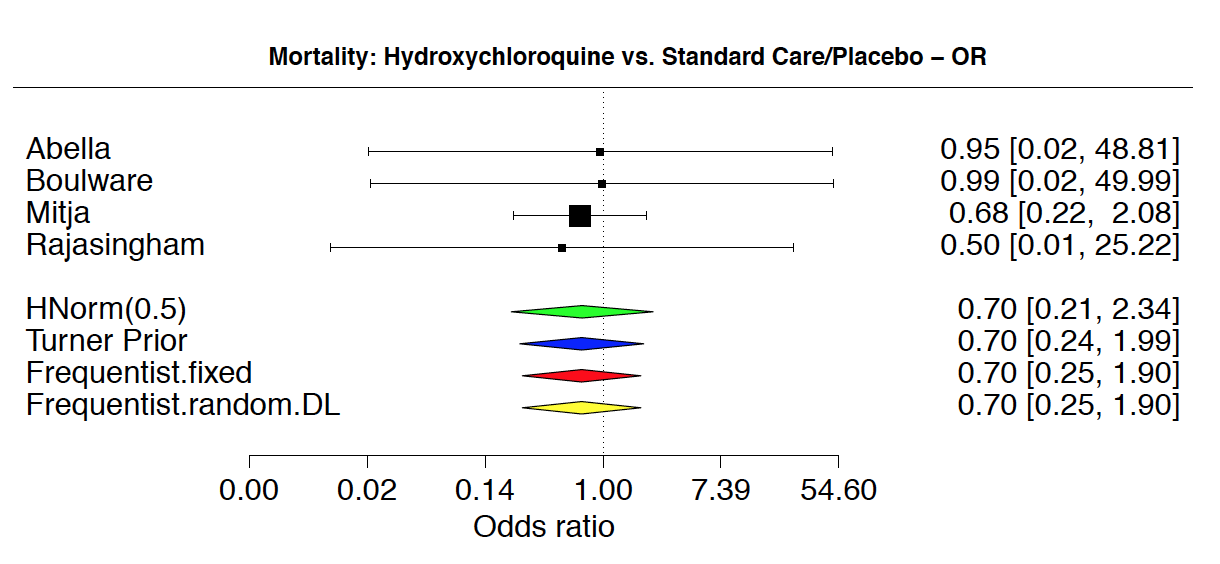

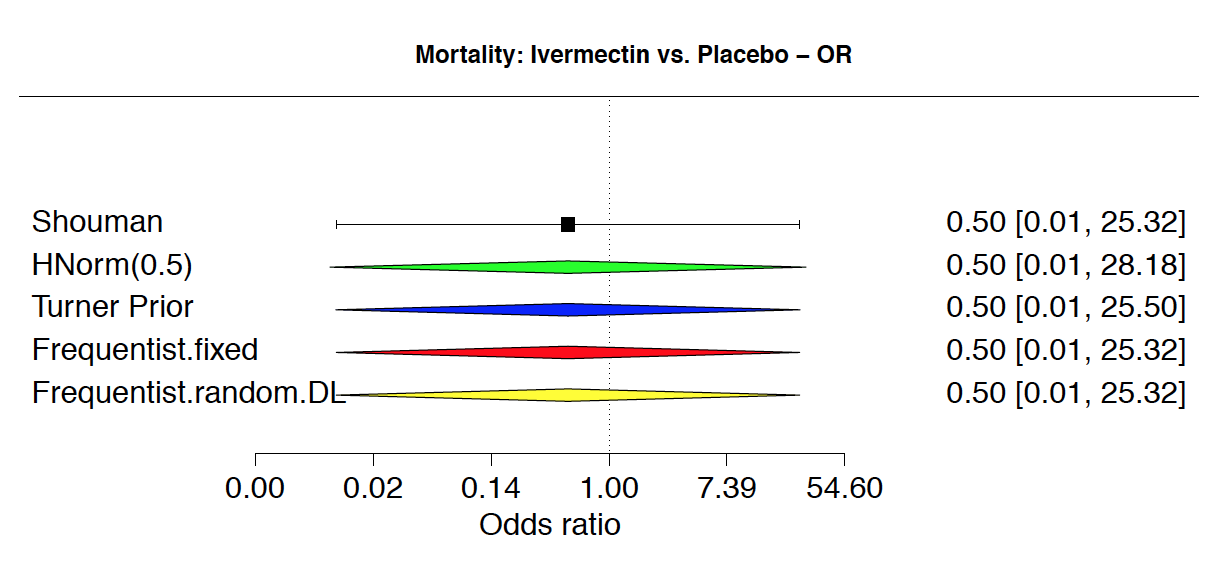

*Adverse effects leading to drug discontinuation*

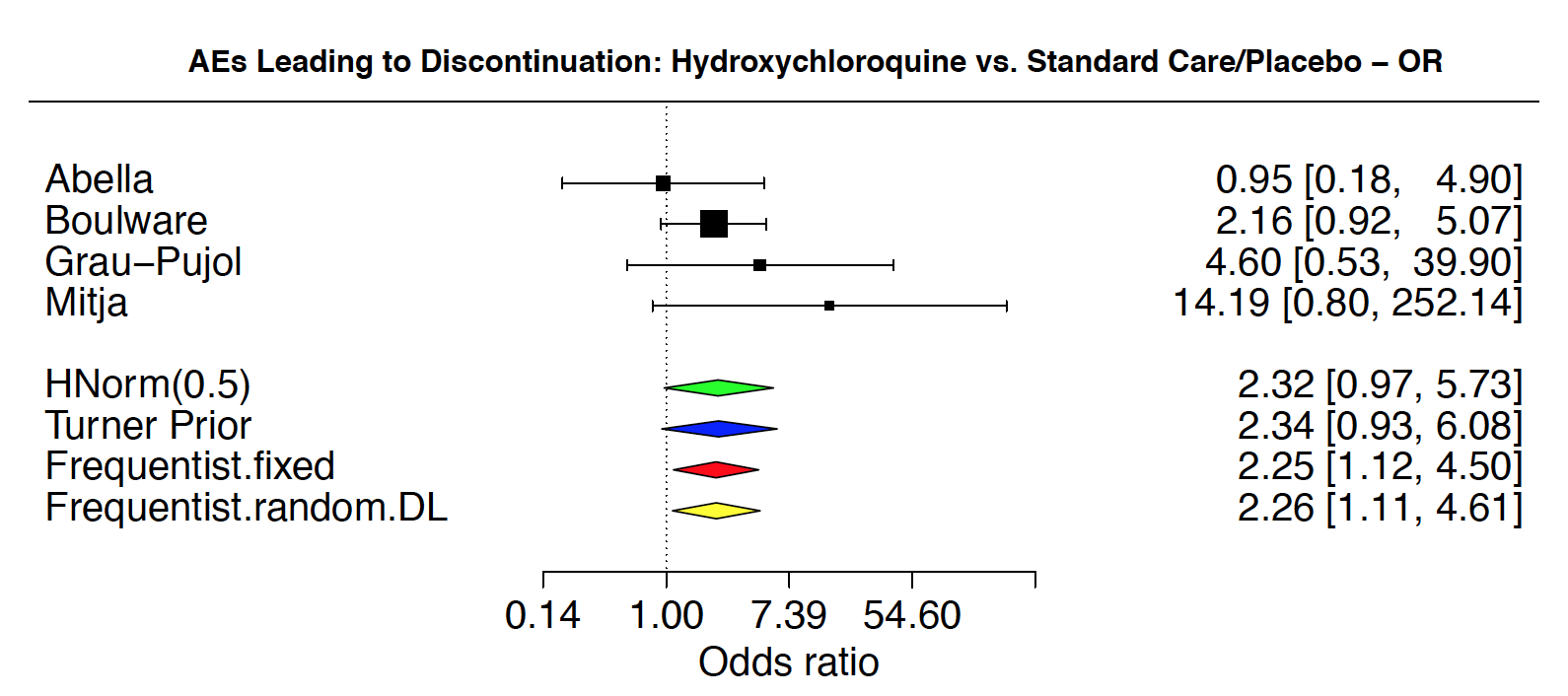

*Adverse effects: cardiac toxicity*

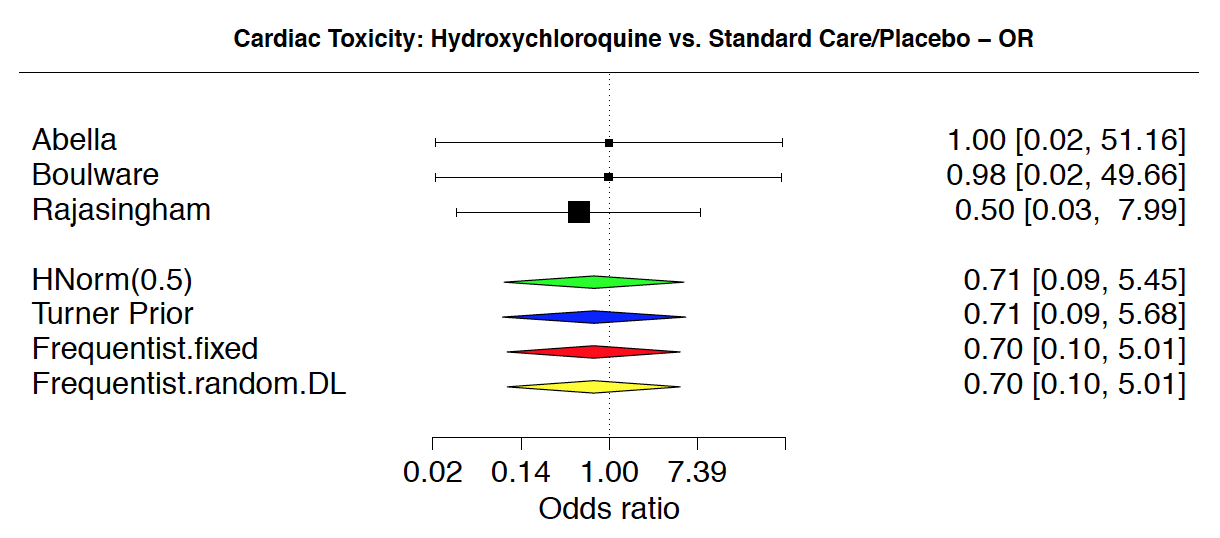

*Adverse effects: non-serious gastrointestinal adverse effects*

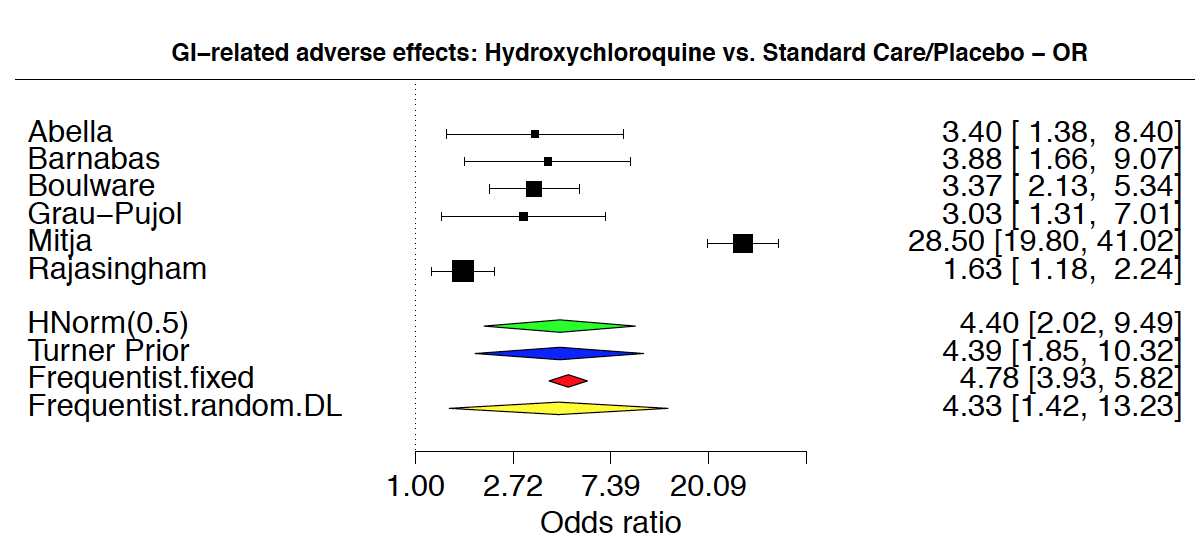

**Completed network meta-analysis results and GRADE**

| **Comparison** | | **Direct estimate** | | | | | | **Indirect estimate** | | | | | | **Network estimate** | | | | | | **Rating** | |
| --- | --- | --- | --- | --- | --- | --- | --- | --- | --- | --- | --- | --- | --- | --- | --- | --- | --- | --- | --- | --- | --- |
| Treatment 1 | Treatment 2 | **Relative effects** | | | **Absolute effect per 1,000** | | | **Relative effects** | | | **Absolute effect per 1,000** | | | **Relative effects** | | | **Absolute effect per 1,000** | | | Final rating | Reasons |
|  |  | Point estimate | CI lower limit | CI upper limit | Point estimate | CI lower limit | CI upper limit | Point estimate | CI lower limit | CI upper limit | Point estimate | CI lower limit | CI upper limit | Point estimate | CI lower limit | CI upper limit | Point estimate | CI lower limit | CI upper limit |  |  |
| **LABORATORY-CONFIRMED SARS-COV-2 INFECTION** | | | | | | | | | | | | | | | | | | | | | |
| hydroxychloroquine | standard care/placebo | 1.03 | 0.71 | 1.47 | 2.00 | -18.00 | 28.00 | 1.03 | 0.71 | 1.47 | 1.52 | -16.90 | 25.90 | 1.03 | 0.71 | 1.47E+00 | 1.52 | -16.90 | 25.90 | MODERATE | risk of bias |
| ivermectin | standard care/placebo | 0.18 | 0.03 | 1.14 | -53.00 | -63.00 | 8.00 | 0.16 | 0.02 | 0.73 | -50.10 | -58.80 | -15.70 | 0.16 | 0.02 | 0.73 | -50.10 | -58.80 | -15.70 | VERY LOW | risk of bias, imprecision 2x |
| ivermectin | hydroxychloroquine | NA | NA | NA | NA | NA | NA | 0.15 | 0.02 | 7.40E-01 | -50.20 | -77.30 | -13.50 | 0.15 | 0.02 | 0.74 | -50.20 | -77.30 | -13.50 | VERY LOW | risk of bias, imprecision 2x |
| iota-carrageenan + ivermectin | standard care/placebo | 0.13 | 0.03 | 0.56 | -56.00 | -63.00 | -28.00 | 0.12 | 0.03 | 0.38 | -52.40 | -58.10 | -36.60 | 0.12 | 0.03 | 0.38 | -52.40 | -58.10 | -36.60 | VERY LOW | risk of bias, imprecision 2x |
| iota-carrageenan + ivermectin | hydroxychloroquine | NA | NA | NA | NA | NA | NA | 0.12 | 0.03 | 0.39 | -53.10 | -78.60 | -29.90 | 0.12 | 0.03 | 0.39 | -53.10 | -78.60 | -29.90 | VERY LOW | risk of bias, imprecision 2x |
| iota-carrageenan + ivermectin | ivermectin | NA | NA | NA | NA | NA | NA | 0.76 | 0.10 | 8.13 | -2.10 | 15.60 | -36.50 | 0.76 | 0.10 | 8.13 | -2.10 | 15.60 | -36.50 | VERY LOW | risk of bias, imprecision 2x |
| **SUSPECTED, PROBABLE OR LABORATORY-CONFIRMED SARS-COV-2 INFECTION** | | | | | | | | | | | | | | | | | | | | | |
| hydroxychloroquine | standard care/placebo | 0.89 | 0.59 | 1.31 | -16.00 | -61.00 | 41.00 | 0.90 | 0.58 | 1.31 | -15.00 | -63.60 | 41.30 | 0.90 | 0.58 | 1.31E+00 | -15.00 | -63.60 | 41.30 | LOW | risk of bias, imprecision |
| ivermectin | standard care/placebo | 0.06 | 0.02 | 0.18 | -155.00 | -163.00 | -132.00 | 0.06 | 0.02 | 0.13 | -159.00 | -165.00 | -144.00 | 0.06 | 0.02 | 0.13 | -159.00 | -165.00 | -144.00 | VERY LOW | risk of bias, imprecision 2x |
| ivermectin | hydroxychloroquine | NA | NA | NA | NA | NA | NA | 0.06 | 0.02 | 1.70E-01 | -143.00 | -199.00 | -91.70 | 0.06 | 0.02 | 0.17 | -143.00 | -199.00 | -91.70 | LOW | risk of bias, imprecision |
| **HOSPITAL ADMISSION** | | | | | | | | | | | | | | | | | | | | | |
| hydroxychloroquine | standard care/placebo | 0.87 | 0.42 | 1.77 | -1.00 | -3.00 | 4.00 | NA | NA | NA | NA | NA | NA | NA | NA | NA | NA | NA | NA | HIGH | NA |
| **MORTALITY** | | | | | | | | | | | | | | | | | | | | | |
| hydroxychloroquine | standard care/placebo | 0.70 | 0.24 | 1.99 | -1.00 | -2.00 | 3.00 | NA | NA | NA | NA | NA | NA | NA | NA | NA | NA | NA | NA | HIGH | NA |
| ivermectin | standard care/placebo | 0.50 | 0.01 | 25.50 | -1.00 | -3.00 | 68.00 | NA | NA | NA | NA | NA | NA | NA | NA | NA | NA | NA | NA | VERY LOW | risk of bias, imprecision 2X |
| ivermectin | hydroxychloroquine | NA | NA | NA | NA | NA | NA | NA | NA | NA | NA | NA | NA | NA | NA | NA | NA | NA | NA | NA | NA |
| **ADVERSE EFFECTS LEADING TO DRUG DISCONTINUATION** | | | | | | | | | | | | | | | | | | | | | |
| hydroxychloroquine | standard care/placebo | 2.34 | 0.93 | 6.08 | 19.00 | -1.00 | 70.00 | NA | NA | NA | NA | NA | NA | NA | NA | NA | NA | NA | NA | MODERATE | imprecision |
| **TIME TO SYMPTOM RESOLUTION OR CLINICAL IMPROVEMENT** | | | | | | | | | | | | | | | | | | | | | |
| NA | NA | NA | NA | NA | NA | NA | NA | NA | NA | NA | NA | NA | NA | NA | NA | NA | NA | NA | NA | NA | NA |

**Subgroup Analyses**

**Hydroxychloroquine for Prophylaxis**

Estimation of subgroup effects based on exposure status and

meta-regression based on dosing regimen

METHODS

We tested for subgroup effects based on whether trials randomised participants to prophylaxis before or after exposure to covid-19 using bayesian hierarchical regression models, with study as a random effect. As suggested by Turner et al. (2015) were used, we used a plausible prior for the variance parameter and a uniform prior for the effect parameter.

To investigate the impact of dosing regimen on efficacy and safety outcomes, we performed bayesian hierarchical meta-regression models, with study as a random effect. Priors for heterogeneity were set as mentioned above**. Table S1** provides the exposure status and dosing regimen in included studies.

**Table S2** provides the mean and variance of heterogeneity priors included in all hierarchical models.

To investigate the effects of hydroxychloroquine dose on efficacy and safety outcomes, we created separate models for pre- and post-exposure studies. For pre-exposure studies, we determined the continuing dose and entered this as a variable in our meta-regression, expressed in units of 100 mg/week doses. For post-exposure studies, we determined the cumulative dose and entered this as a variable in our meta-regression, expressed in units of 1000mg.

Treatment duration refers to the period of time (in days) for which hydroxychloroquine was taken.

Because of inconsistent reporting observed across trials, we analysed non-serious GI adverse events as a composite of diarrhea, nausea, abdominal discomfort, and vomiting. If a study reported any of these adverse events individually, we analyzed the outcome with the highest number of events to avoid double counting of participants.

RESULTS

Hierarchical estimation of effects of COVID-19 exposure status and hydroxychloroquine dosing on efficacy outcomes

| Variable | n trials | n events/N Total | | β treatment^[1]^ | | β treatment*variable^[2]^ | | |
| --- | --- | --- | --- | --- | --- | --- | --- | --- |
|  |  | HCQ | SC | OR | 95% CrI | OR | 95% CrI | Pr(OR >1) |
| SARS CoV-2 - Laboratory confirmed infection | | | | | | | | |
| Exposure status^[3]^ | 6 | 142/2875 | 135/2419 | 1.02 | 0.72, 1.45 | 0.827 | 0.35, 1.97 | 0.33 |
| Continuing dose (per 100mg/week)^[4]^ | 3 | 15/1190 | 11/671 | 0.76 | 0.25, 2.27 | 0.664 | 0.23, 1.70 | 0.26 |
| Cumulative dose (per 1000mg)^[5]^ | 3 | 127/1685 | 124/1748 | 0.89 | 0.15, 5.53 | 1.02 | 0.25, 4.18 | 0.52 |
| SARS CoV-2 – Suspected, probable or laboratory confirmed or suspected infection | | | | | | | | |
| Exposure status | 3 | 286/2321 | 282/1906 | 0.90 | 0.59, 1.30 | 0.79 | 0.37, 1.80 | 0.25 |
| Continuing dose (per 100mg/week) | 1 | 58/989 | 39/494 | -- | -- | -- | -- | -- |
| Cumulative dose (per 1000mg) | 2 | 228/1332 | 243/1412 | 1.25 | 0.23, 6.89 | 0.96 | 0.24, 3.86 | 0.48 |
| Hospital admission | | | | | | | | |
| Exposure status | 5 | 24/3021 | 22/2638 | 0.86 | 0.43, 1.70 | 0.905 | 0.28, 2.89 | 0.43 |
| Continuing dose (per 100mg/week) | 2 | 11/1053 | 7/555 | 0.62 | 0.15, 2.56 | 1.041 | 0.26, 4.06 | 0.53 |
| Cumulative dose (per 1000mg) | 3 | 13/1968 | 15/2083 | 1.01 | 0.16, 6.36 | 0.99 | 0.24, 4.10 | 0.49 |

Hierarchical estimation of hydroxychloroquine dosing, and duration of hydroxychloroquine prophylaxis on safety outcomes

| Variable | n trials | n events/N Total | | β treatment^[6]^ | | β treatment*variable^[7]^ | | |
| --- | --- | --- | --- | --- | --- | --- | --- | --- |
|  |  | HCQ | SC | OR | 95% CrI | OR | 95% CrI | Pr(OR >1) |
| Adverse effects leading to discontinuation | | | | | | | | |
| Duration of therapy (per day) | 4 | 31/1767 | 12/1849 | 1.93 | 0.70, 5.47 | 0.99 | 0.95, 1.04 | 0.39 |
| Continuing dose (per 100mg/week)^[8]^ | 2 | 8/206 | 4/188 | 2.03 | 0.46, 9.12 | 1.45 | 0.72, 3.06 | 0.85 |
| Cumulative dose (per 1000mg)^[9]^ | 2 | 23/1561 | 8/1661 | 1.28 | 0.19, 8.08 | 1.15 | 0.28, 4.66 | 0.58 |
| Cardiac toxicity | | | | | | | | |
| Duration of therapy (per day) | 3 | 1/1468 | 1/966 | 0.97 | 0.15, 6.62 | 1.62 | 0.86, 4.62 | 0.88 |
| Continuing dose (per 100mg/week) | 2 | 1/1054 | 1/559 | 0.90 | 0.15, 5.53 | 0.94 | 0.23, 3.86 | 0.47 |
| Cumulative dose (per 1000mg) | 1 | 0/414 | 0/407 | -- | -- | -- | -- | -- |
| GI-related adverse effects | | | | | | | | |
| Duration of therapy (per day) | 6 | 833/3214 | 140/2815 | 4.22 | 1.54, 9.78 | 0.99 | 0.95, 1.04 | 0.36 |
| Continuing dose (per 100mg/week) | 3 | 218/1196 | 73/686 | 1.75 | 0.81, 3.78 | 1.221 | 0.83, 1.82 | 0.87 |
| Cumulative dose (per 1000mg) | 3 | 615/2018 | 67/2129 | 1.39 | 0.23, 9.12 | 1.284 | 0.31, 5.53 | 0.63 |

**Table S1.** Lognormal heterogeneity priors used in Bayesian hierarchical models based on Turner *et al.*

| Outcome | Outcome category | Tau (mu, sigma) |
| --- | --- | --- |
| Adverse effects leading to discontinuation | adverse events | -0.935, 0.76 |
| Cardiac toxicity | adverse events | -0.935, 0.76 |
| Hospital admission | resource use / hospital stay / process | -1.17, 0.87 |
| Laboratory confirmed SARS CoV-2 infection | infection / onset of new disease | -1.245, 0.76 |
| Non-serious gastrointestinal adverse effects | adverse events | -0.935, 0.76 |
| Suspected, probable, or confirmed SARS CoV-2 infection | infection / onset of new disease | -1.245, 0.76 |

R.M. Turner, D. Jackson, Y. Wei, S.G. Thompson, J.P.T. Higgins. Predictive distributions for between-study heterogeneity and simple methods for their application in Bayesian meta-analysis. Statistics in Medicine, **34**(6):984-998, 2015.

**Table S2.** COVID-19 exposure status and dosing regimen of included studies.

| Author | Interventions | Exposure Status | Description of dosing regimen in trial report | Cumulative dose | | Continuing dose | | HCQ therapy duration (in days) |
| --- | --- | --- | --- | --- | --- | --- | --- | --- |
|  |  |  |  | mg | per 1,000mg | mg | per 100mg/wk |  |
| Boulware | Hydroxychloroquine; placebo | post-exposure | The dosing regimen for hydroxychloroquine was 800 mg (4 tablets) once, then 600 mg (3 tablets) 6 to 8 hours later, then 600 mg (3 tablets) daily for 4 more days for a total course of 5 days (19 tablets total). If participants had gastrointestinal upset, they were advised to divide the daily dose into two or three doses. | 3800 | 3.8 | NA | NA | 5 |
| Mitja | Hydroxychloroquine; standard care | post-exposure | Participants allocated in the control arm received no treatment aside from usual care, whereas those in the intervention arm received HCQ (Dolquine®) 800 mg on day 1, followed by 400 mg once daily for six days. | 3200 | 3.2 | NA | NA | 7 |
| Rajasingham | hydroxychloroquine (once weekly); placebo | pre-exposure | Participants were randomly assigned in a 2:2:1:1 ratio to receive hydroxychloroquine given as a loading dose of 400mg (two 200mg tablets) twice separated by 6-8 hours followed by (i) 400 mg (two 200mg tablets) once weekly for 12 weeks or (ii) 400 mg (two 200mg tablets) twice weekly for 12 weeks, or to placebo which was prescribed in a matched fashion including a loading dose of two tablets followed by two tablets once or twice weekly for 12 weeks. | NA | NA | 400/week | 4 | 84 |
| Rajasingham | hydroxychloroquine (twice weekly); placebo | pre-exposure | Participants were randomly assigned in a 2:2:1:1 ratio to receive hydroxychloroquine given as a loading dose of 400mg (two 200mg tablets) twice separated by 6-8 hours followed by (i) 400 mg (two 200mg tablets) once weekly for 12 weeks or (ii) 400 mg (two 200mg tablets) twice weekly for 12 weeks, or to placebo which was prescribed in a matched fashion including a loading dose of two tablets followed by two tablets once or twice weekly for 12 weeks. | NA | NA | 800/week | 8 | 84 |
| Abella | Hydroxychloroquine; placebo | pre-exposure | Participants assigned to the hydroxychloroquine arm received hydroxychloroquine 200-mg tablets (provided by Sandoz, a division of Novartis Pharmaceuticals), with instructions to take 3 tablets once a day with food. Participants assigned to the placebo arm received custom-molded identically sized and shaped microcrystalline cellulose tablets (prepared for this trial by Temple IDS; Temple University; Philadelphia, Pennsylvania) and given identical instructions. | NA | NA | 4200/week | 42 | 56 |
| Grau-Pujol | Hydroxychloroquine; placebo | pre-exposure | Participants allocated to intervention arm (PrEP) received 400 mg of hydroxychloroquine (two tablets of 200 mg) daily the first four consecutive days, followed by 400 mg weekly during the study period, initially scheduled to be 6 months. Participants in the control group followed the same treatment schedule with placebo tablets that were indistinguishable from hydroxychloroquine tablets. | NA | NA | 400/week | 4 | 30 |
| Barnabas | Hydroxychloroquine; placebo | post-exposure | Households were randomly assigned in a 1:1 ratio to receive either hydroxychloroquine (400 mg/d orally for 3 days, then 200 mg/d orally for an additional 11 days) or ascorbic acid (500 mg/d orally for 3 days, then 250 mg/d orally for 11 days) as a placebo equivalent. | 3400 | 3.4 | NA | NA | 14 |

1. Estimate of the effect of HCQ on outcome, adjusting for variable of interest. For example, if row corresponds to “duration of therapy” for adverse effects leading to discontinuation, this OR represents the effect of HCQ on adverse effects, adjusting for duration of HCQ therapy (in days). An OR of 1.93 means that for patients taking HCQ as prophylaxis, the odds of adverse effects increases two-fold, compared to those who are not taking HCQ.
2. Estimate of the magnitude of interaction between variable of interest and treatment on outcome. For example, if the row corresponds to “exposure status” for laboratory confirmed infection, this OR represents the magnitude of interaction between the HCQ therapy and exposure status. If the OR is 0.83 and HCQ is effective as a prophylactic drug, HCQ is more effective in patients who take HCQ before exposure to COVID-19.
3. Pre-exposure prophylaxis coded as “1” and post-exposure prophylaxis coded as “0.” Therefore, OR of interaction corresponds to the difference in the odds of achieving the outcome if HCQ taken for prophylaxis before exposure to COVID-19.
4. Analysis limited to pre-exposure prophylaxis trials only.
5. Analysis limited to post-exposure prophylaxis trials only.
6. Estimate of the effect of HCQ on outcome, adjusting for variable of interest. For example, if row corresponds to “duration of therapy” for adverse effects leading to discontinuation, this OR represents the effect of HCQ on adverse effects, adjusting for duration of HCQ therapy (in days). An OR of 1.93 means that for patients taking HCQ as prophylaxis, the odds of adverse effects increases two-fold, compared to those who are not taking HCQ.
7. Estimate of the magnitude of interaction between variable of interest and treatment on outcome. For example, if row corresponds to “continuing dose” for cardiac toxicity, this OR represents the magnitude of interaction between HCQ therapy and per 100mg weekly dose increases in HCQ. For example, if the OR of interaction is 1.62 and the intervention causes cardiac toxicity, higher weekly HCQ doses are associated with increased events of cardiac toxicity.
8. Analysis limited to pre-exposure prophylaxis trials only.
9. Analysis limited to post-exposure prophylaxis trials only.
